# Heightened resistance to type 1 interferons characterizes HIV-1 at transmission and following analytical treatment interruption

**DOI:** 10.1101/2020.08.24.20181149

**Authors:** Marcos V. P. Gondim, Scott Sherrill-Mix, Frederic Bibollet-Ruche, Ronnie M. Russell, Stephanie Trimboli, Andrew G. Smith, Yingying Li, Weimin Liu, Alexa N. Avitto, Julia DeVoto, Jesse Connell, Angharad E. Fenton-May, Pierre Pellegrino, Ian Williams, Emmanouil Papasavvas, Julio C. C. Lorenzi, D. Brenda Salantes, Felicity Mampe, M. Alexandra Monroy, Yehuda Z. Cohen, Sonya Heath, Michael S. Saag, Luis J. Montaner, Ronald G. Collman, Janet M. Siliciano, Robert F. Siliciano, Lindsey Plenderleith, Paul M. Sharp, Marina Caskey, Michel C. Nussenzweig, George M. Shaw, Persephone Borrow, Katharine J. Bar, Beatrice H. Hahn

**Affiliations:** Department of Medicine, University of Pennsylvania, Philadelphia, PA 19104, USA; Department of Microbiology, University of Pennsylvania, Philadelphia, PA 19104, USA; Nuffield Department of Clinical Medicine, University of Oxford, Oxford OX3 7FZ, UK; Centre for Sexual Health and HIV Research, University College London, London WC1E 6JB, United Kingdom; Vaccine and Immunotherapy Center, The Wistar Institute, Philadelphia, PA 19104, USA; Laboratory of Molecular Immunology, The Rockefeller University, New York, NY 10065, USA; Department of Medicine, University of Alabama at Birmingham, Birmingham, AL 35294, USA; Department of Medicine, Johns Hopkins University, Baltimore, MD 21205, USA; Howard Hughes Medical Institute, Johns Hopkins University, Baltimore, MD 21205, USA; Institute of Evolutionary Biology, University of Edinburgh, Edinburgh EH9 3FL, United Kingdom; Centre for Immunity, Infection and Evolution, University of Edinburgh, Edinburgh EH9 3FL, United Kingdom; Howard Hughes Medical Institute, The Rockefeller University, New York, NY 10065, USA

## Abstract

Type 1 interferons (IFN-I) are potent innate antiviral effectors that constrain HIV-1 transmission. However, harnessing these cytokines for HIV-1 cure strategies has been hampered by an incomplete understanding of their anti-viral activities at later stages of infection. Here, we characterized the IFN-I sensitivity of 500 clonally-derived HIV-1 isolates from plasma and CD4+ T cells of 26 individuals sampled longitudinally following transmission and/or after antiretroviral therapy (ART) and analytical treatment interruption (ATI). Determining the concentration of IFNα2 and IFNβ that reduced HIV-1 replication by 50% (IC_50_), we found remarkably consistent changes in the sensitivity of viruses to IFN-I inhibition, both across individuals and over time. IFN-I resistance was uniformly high during acute infection, decreased in all subjects in the first year post-infection, was reacquired concomitant with CD4+ T cell loss, and remained elevated in subjects with accelerated disease. Isolates obtained by viral outgrowth during suppressive ART were relatively IFN-I sensitive, resembling viruses circulating just prior to ART initiation. However, viruses that rebounded following treatment interruption displayed the highest levels of IFNα2 and IFNβ resistance observed at any time during the infection course. These findings indicate a dynamic interplay between host innate immune responses and the evolving HIV-1 quasispecies, with the relative contribution of IFN-I to HIV-1 control impacted by both ART and ATI. Although elevated at transmission, IFN-mediated pressures are the highest during viral rebound, limiting the viruses that successfully reactivate from latency.

**One Sentence Summary:** HIV-1 resistance to IFN-I is highest during acute infection and following analytic treatment interruption, indicating a dynamic interplay between host innate immunity and virus biology.

## Introduction

Type 1 interferon (IFN-I) comprises a family of pro-inflammatory and immunomodulatory cytokines with potent antiviral activity (*1, 2*). In humans, this family includes 13 IFNα subtypes as well as IFNβ, IFNε, IFNκ, and IFNω, of which IFNα and IFNβ are the best characterized (*3*). IFN-I is rapidly up-regulated in response to pathogen exposure and infection (*4, 5*), and mediates its activity by binding to the heterodimeric IFNAR receptor expressed on all nucleated cells (*6*). This binding results in the activation of signal transduction cascades that trigger the expression of hundreds of interferon-stimulated genes (ISGs), which have direct and indirect antiviral activity, and regulate the activation state, function, proliferation and survival of host immune cells (*7, 8*). Although all IFN-I subtypes bind to the same receptor, they differ in their affinity for the two IFNAR subunits and elicit different patterns of ISG expression, suggesting that they vary in their *in vivo* activity and potency (*9-12*).

Up-regulation of IFN-I expression is one of the earliest innate responses to infection with both human (HIV-1) and simian (SIV) immunodeficiency viruses (*13-17*). In acute HIV-1 infection, maximal IFN-I levels are detected in the plasma prior to peak viremia (*18*). In acute SIVmac infection of macaques, plasmacytoid dendritic cells (pDCs) are actively recruited to mucosal sites, where they produce high levels of both IFNα and IFNβ (*19, 20*). IFN-I is known to control HIV/SIV replication in CD4+ T cells and macrophages by targeting multiple steps in the viral lifecycle (*8*). Consistent with this, pre-treatment of rhesus macaques with exogenous IFNα2 increased the number of intrarectal challenges required to achieve systemic SIVmac infection (*21*) and vaginal administration of IFNβ protected rhesus macaques from recombinant simian/human immunodeficiency virus (SHIV) infection (*22*). Likewise, blocking IFN-I during acute SIVmac infection led to higher viral titers, more rapid decline of CD4+ T cells, and faster disease progression (*23*). Thus, numerous studies in both humans and primates have shown that IFN-I play a key role in controlling HIV/SIV replication during the earliest stages of infection.

In contrast to the beneficial effects of IFN-I upregulation during acute HIV-1 infection, sustained IFN-I signaling during chronic infection appears to have an overall negative effect (*17, 24-28*). In untreated subjects, plasma IFNα levels are positively correlated with HIV-1 viral load and inversely correlated with CD4+ T cell counts, and individuals with higher viral loads and faster disease progression frequently exhibit higher ISG expression (*24, 27*) and heightened IFNβ production in the gut (*29*). Continuous IFN-I signaling increases the number of susceptible CD4+ target cells, induces CD4 T cell apoptosis, limits antigen specific CD4+ and CD8+ T cell responses, and contribute to immune exhaustion (*15, 30, 31*). In SIVmac infected macaques, prolonged IFNα2 treatment resulted in decreased antiviral ISG expression, an increased SIVmac reservoir size, and the loss of CD4+ T cells (21). Indeed, sustained IFN-I signaling and ISG up-regulation differentiates pathogenic from non-pathogenic SIV infections *(32-34)* and drives systemic immune activation in HIV-1 infection, which is associated with poor CD4+ T-cell recovery and a predictor of disease progression (*35*).

Given the highly pleiotropic and context-dependent effects of IFN-I, it is not surprising that clinical studies evaluating its therapeutic benefits have yielded mixed results *(14, 16, 36)*. IFNα2 administration can suppress HIV-1 replication, blunt rebound viremia, and extend viral control following analytic treatment interruptions (ATI) (*37-40*). However, various combinations of anti-retroviral and IFN-I treatments have not led to long-term improvements of CD4 T cell reconstitution or clinical outcome (*14*). Moreover, blocking rather than stimulating IFN-I signaling, or selectively counteracting only certain ISGs, had beneficial effects in HIV-1 infected humanized mice (*41, 42*). Thus, it seems clear that effective manipulation of the IFN-I system to prevent, treat or cure HIV-1 infection will require a much more detailed understanding of the kinetics, potency and quality of the endogenous IFN-I response.

One approach to gain insight into the *in vivo* IFN-I response is to test the IFN sensitivity of plasma viruses, which are reliable indicators of selection pressures that act on virions and virus-infected cells (*43-48*). Such studies have shown that transmitted founder (TF) viruses are highly IFN-I resistant (*49*) and that this resistance constitutes a major determinant of HIV-1 transmission fitness (*50*). In contrast, viruses isolated during chronic infection are generally IFN-I sensitive (*45, 46, 47*), although this sensitivity seems to revert as subjects progress toward AIDS (*51, 52*). Here, we performed a systematic analysis of the IFN-I sensitivity of plasma and CD4+ T cell derived viral isolates from prospectively sampled subjects before and after antiretroviral therapy (ART), and following analytical treatment interruption (ATI). We found that the *in vivo* IFN-I response is much more dynamic than previously assumed and places varying pressures on the viral population at different stages of the infection. In particular, the high IFN-I resistance of rebound viruses indicates intensified IFN-I activity during viral recrudescence.

## Results

### Generation of plasma isolates from longitudinally sampled individuals

To characterize the kinetics of IFN-I resistance over the course of HIV-1 infection, we generated plasma isolates from 10 prospectively sampled individuals, who were followed from shortly after transmission until 4.1 - 12.4 years post-infection (table S1). All participants were men-who-have-sex-with-men (MSM) who presented with symptomatic primary subtype B infection. Peripheral blood was collected at regular intervals immediately following onset of symptoms and continued for 74 to 318 weeks (1.6 to 6.2 years) in the absence of antiretroviral treatment. At each time point, patients underwent clinical evaluation, including viral load and CD4+ T cell count determinations (table S1). Six individuals experienced a gradual loss of their CD4+ T cells over the study period (typical progressors; Fig. 1A), two individuals maintained high CD4+ T cell counts and low viral loads throughout their 3.6- and 5.6-year clinical follow-up (non-progressors; Fig. 1B), and two individuals developed AIDS (CD4 T cells < 300 cell/μl) within 0.7 and 1.5 years of infection (rapid progressors; Fig. 1C). ART was initiated in eight participants between 85 and 323 weeks post onset of symptoms based on the standard-of-care at the time (shaded in Fig. 1).

**Fig. 1.**
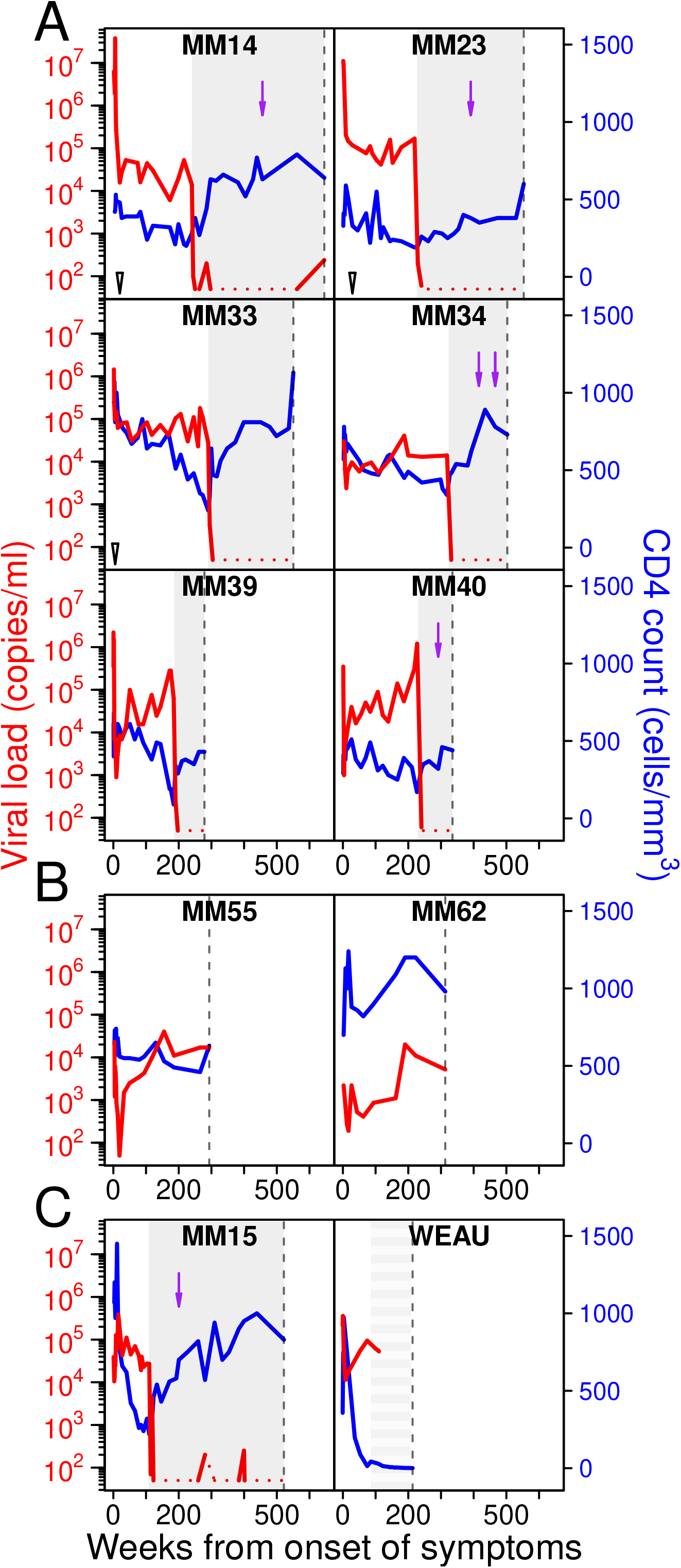
HIV-1 viral loads and CD4+ T cell counts in prospectively studied individuals. Viral loads (RNA copies/ml; red) and CD4+ T cell counts (cells/μl; blue) are shown for 10 acutely infected individuals (y-axis), who were followed from the onset of symptoms until 213 - 645 weeks (4.1 - 12.4 years) post-infection (x-axis). Participants are grouped based on disease progression, with (A) including typical progressors, (B) non-progressors, and (C) rapid progressors. Gray shading indicates suppressive antiretroviral therapy (alternating shading in WEAU indicates non-suppressive zidovudine monotherapy). Purple arrows denote the time points of PBMC samples that yielded QVOA isolates, open triangles denote superinfection, and dashed vertical lines indicate termination of the study or loss to follow up.

We also characterized the evolving HIV-1 quasispecies in these subjects. This was done by amplifying viral RNA directly from the plasma using single genome amplification (SGA), which retains genetic linkage across viral genes and generates sequences devoid of PCR-induced artifacts (*53, 54*). For each individual, 3’ half genomes or *env* gene sequences were amplified from sequential plasma samples, with sequences from the earliest time point used for TF enumeration (fig. S1). This analysis showed that seven participants acquired a single TF virus, while the remaining three became infected with two or more TF viruses (table S1). Sequences from subsequent time points exhibited the expected patterns of viral diversification (*55-57*), except for MM14, MM23 and MM33, who became superinfected with additional subtype B strains 5 to 29 weeks following the initial infection (Fig. 1 and fig. S2).

To determine the IFN sensitivity of individual quasispecies members at different stages of HIV-1 infection, we generated 277 limiting dilution plasma isolates from five of the 10 subjects (table S1). Plasma samples were end-point diluted, co-cultured with healthy donor CD4+ T-cells, and the resulting isolates sequenced to ensure that they were indeed single virion-derived (50). In phylogenetic trees of *env* gene sequences, the limiting dilution isolates were completely interspersed with sequences amplified directly from the plasma, thus confirming their authenticity (fig. S2). To characterize their sensitivity to IFN-I inhibition, we determined the IFNα2 and IFNβ concentrations that reduced virus replication *in vitro* by 50% (50). Briefly, donor CD4+ T cells were treated with increasing quantities of IFNα2 and IFNβ, and then infected with equal amounts of virus (50). Cells were cultured for 7 days, with the IFN-containing media replenished every second day. Virus replication was measured for each IFN-I concentration by quantifying p24 antigen in supernatants and used to calculate the half-maximal inhibitory concentrations (IC_50_) (fig. S3).

Since the IFNα2 and IFNβ IC_50_ values for limiting dilution isolates from the same plasma sample were very similar (Fig. 2), we reasoned that conventional (bulk) virus isolation was likely sufficient to capture the IFN-I sensitivity of the circulating virus pool. To test this, we generated 18 bulk isolates from 11 of these plasma samples (table S1) and determined their IFN-I resistance (table S2). All bulk isolates yielded IC_50_ values that fell within 5-fold of the mean of the corresponding limiting dilution isolates (Fig. 2; note that the IC_50_ range spanned over 1,000-fold for IFNα2 and over 40,000-fold for IFNβ). Moreover, the mean interferon resistance of the bulk isolates was highly correlated with that of the corresponding limiting dilution isolates (IFNα2 r=0.95 and IFNβ r=0.96). We thus used the less labor-intensive conventional approach to generate 79 plasma isolates for the remaining five subjects.

**Fig. 2.**
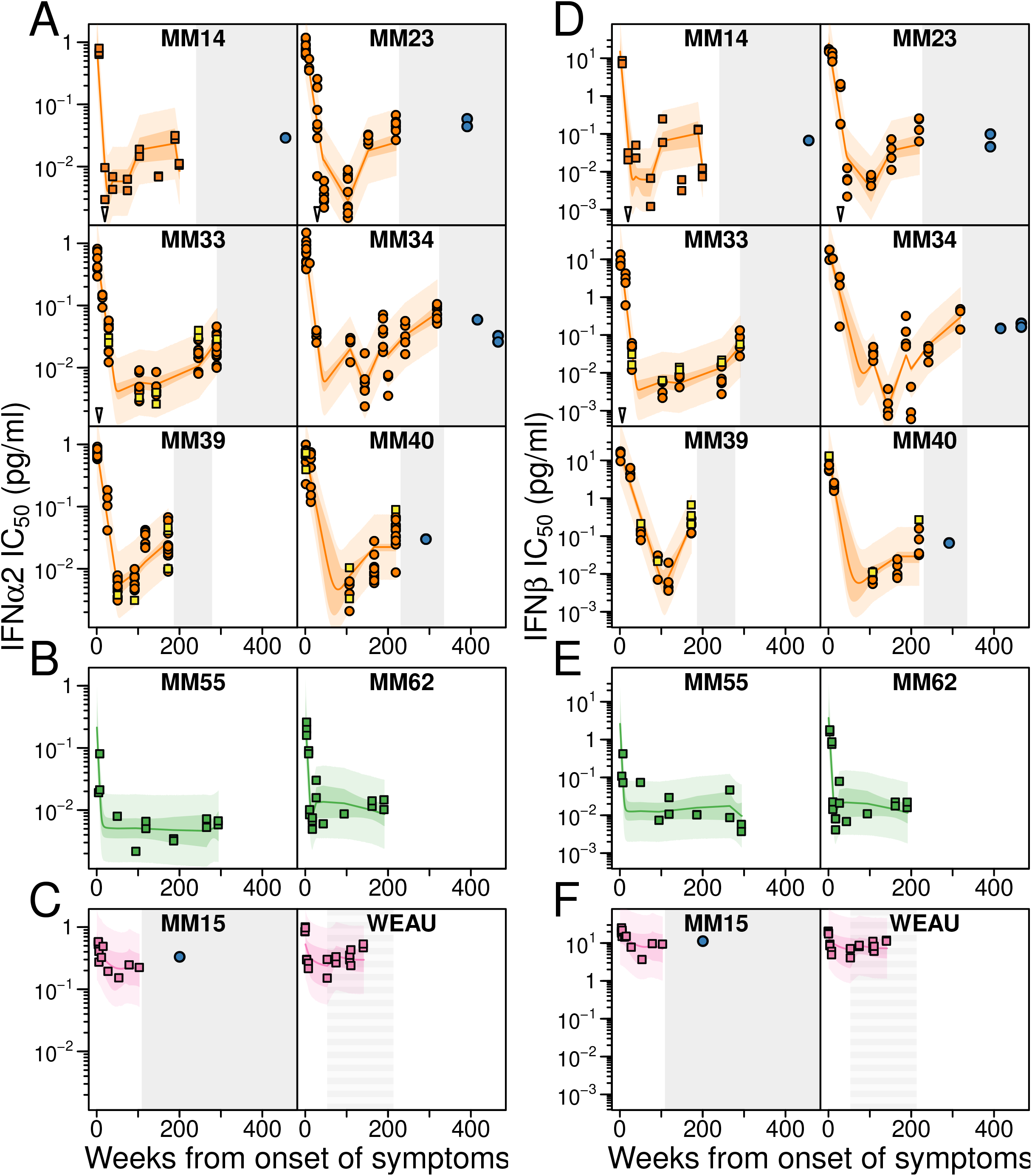
Kinetics of IFN-I resistance over the course of HIV-1 infection. IFNα2 (A-C) and IFNβ (D-F) IC_50_ values (pg/ml) are shown for limiting dilution (circles) and bulk culture-derived (squares) plasma isolates from the onset of symptoms until 140 - 466 weeks (2.7 – 8.9 years) post-infection (yellow squares indicate bulk isolates from plasma samples that also yielded limiting dilution isolates). Colored lines denote the average IC_50_ values as estimated by a Bayesian model, with darker shading indicating the 95% credible and lighter shading the 95% prediction intervals, respectively. Gray shading indicates suppressive antiretroviral therapy (alternating shading in WEAU indicates non-suppressive zidovudine monotherapy). Blue circles indicate isolates obtained by viral outgrowth from CD4+ T cells after years of suppressive ART. Open triangles denote superinfection (fig. S2). Participants are grouped as in Fig. 1, with panels A, B and C showing typical progressors (orange), non-progressors (green), and rapid progressors (pink), respectively.

### Kinetics of IFN-I resistance in untreated HIV-1 infection

Analysis of 374 plasma isolates from 10 prospectively sampled subjects revealed marked differences in their sensitivity to IFNα2 and IFNβ inhibition over the course of the infection (Fig. 2). To quantify these dynamic changes, we developed a Bayesian hierarchical change point model *(50, 58)*, which allowed us to infer the temporal patterns of IFNα2 and IFNβ IC_50_ values and their associations with CD4+ T cell counts across different individuals while accounting for differences in sampling times and frequencies (Supplementary Methods, fig. S4).

Consistent with previous results *(49, 50, 59)*, isolates obtained during acute HIV-1 infection were uniformly IFN-I resistant, yielding mean IC_50_ values for IFNα2 of 0.90 pg/ml and for IFNβ of 19 pg/ml (Fig. 2). This resistance decreased in all subjects, with IFNα2 and IFNβ IC_50_ values falling in typical progressors on average 180-fold (95% Credible Interval [CrI]: 100 - 340-fold) and 3,500-fold (95% CrI: 1500 - 8900-fold) within 340 days (95% CrI: 250 - 490 days) and 460 days (95% CrI: 310 - 700 days), respectively. Isolates from non-progressing individuals fell to similar levels, but reached a nadir earlier at an estimated 100 days (95% CrI: 50 - 200 days) for IFNα2 and 110 days (95% CrI: 50 - 240 days) for IFNβ. The time to nadir for fast progressing individuals was difficult to determine because IFNα2 and IFNβ IC_50_ values decreased only 2.8-fold (95% CrI: 0.78 - 10.5-fold) and 2.7-fold (95% CrI: 0.52 - 16-fold), respectively, resulting in 39-fold and 1,000-fold higher levels than in subjects with more typical disease progression (Fig. 2C and fig. S4, A and B).

Plasma virus from typical progressors exhibited a consistent rise in IFNα2 and IFNβ resistance during the later stages of infection (Fig. 2). This rise and other fluctuations in resistance during chronic infection (e.g., MM34 in Fig. 2), appeared to associate with changes in CD4+ T cell counts. Each decrease of 100 CD4+ T cells/μl was associated with a 2.3-fold (95% CrI: 1.3 - 4.4-fold) increase in mean IFNα2 IC_50_ and a 4.0-fold (95% CrI: 1.9 - 8.7-fold) increase in mean IFNβ IC_50_ values (Fig. 2A and fig. S4, C and D). The two non-progressing individuals maintained their CD4+ T cell counts at relatively high levels and the two fast progressing individuals were largely depleted of CD4+ T cells early in infection. Thus, the model predicted little association between IFN-I resistance and CD4+ T cell counts in these participants (Fig. 2, B and C and fig. S4, C and D).

Overall, the temporal changes in IFN-I resistance were remarkably similar among the different individuals. Without exception, viral IFN-I resistance was high during acute infection, decreased during early chronic infection, and increased again during later stages concomitant with CD4+ T cell loss and disease progression. Remarkably, neither the number of transmitted founder viruses at transmission nor subsequent superinfection events appeared to influence these kinetics (Fig. 2). In all analyses, IFNα2 and IFNβ IC_50_ values were highly correlated (fig. S5), although IFNβ IC_50_ values spanned nearly four orders of magnitude compared to two orders of magnitude for IFNα2 (Fig. 2). These findings thus indicate a highly dynamic interplay between host innate immune responses and the evolving HIV-1 quasispecies, with the relative contribution of IFN-I to HIV-1 control varying markedly, but predictably, over the course of HIV-1 infection.

### IFN-I sensitivity of ex vivo reactivated latent viruses

Having characterized the IFN-I response of plasma virus during untreated infection, we next used a quantitative viral outgrowth assay (QVOA) to generate reactivated latent viruses from some of the same individuals after they were placed on therapy. For five of the 10 longitudinally followed subjects, cryopreserved peripheral blood mononuclear cells (PBMCs) collected 1 to 4 years after ART initiation were available (Fig. 1). These were stimulated, co-cultured with primary CD4+ T cells from healthy donors, and tested for viral replication by monitoring p24 levels in culture supernatants (*60*). Due to limited numbers of cryopreserved cells, only eight QVOA isolates were recovered (Fig. 2 and table S1). All isolates were sequenced (fig. S2) and their IFN-I resistance determined (table S2). Although participant WEAU received zidovudine (AZT) beginning 1.6 years post infection (Figs. 1 and 2), this monotherapy was not sufficient to maintain suppression of plasma viral loads (table S1). Thus, post-treatment samples from this individual were not suitable for QVOA analyses.

As shown in Fig. 2, seven of the eight QVOA isolates were moderately IFN-I resistant, exhibiting IC_50_ values very similar to plasma isolates obtained from the same individuals immediately prior to ART initiation. This was true for individuals with single as well as multiple QVOA isolates, including viruses collected several months apart (MM34). A single QVOA isolate from a rapidly progressing subject (MM15) was more IFN-I resistant, but again plasma virus obtained just prior to treatment initiation displayed the same elevated IFN-I resistance. Phylogenetic analyses confirmed these findings, showing that the eight outgrowth isolates did not cluster with viruses recovered during acute or early infection, but were most closely related to plasma viruses replicating during late infection (Fig. 3 and fig. S2). Thus, both IFN-I and phylogenetic analyses indicated that the QVOA derived viruses had entered the latent reservoir near the time of therapy initiation.

**Fig. 3.**
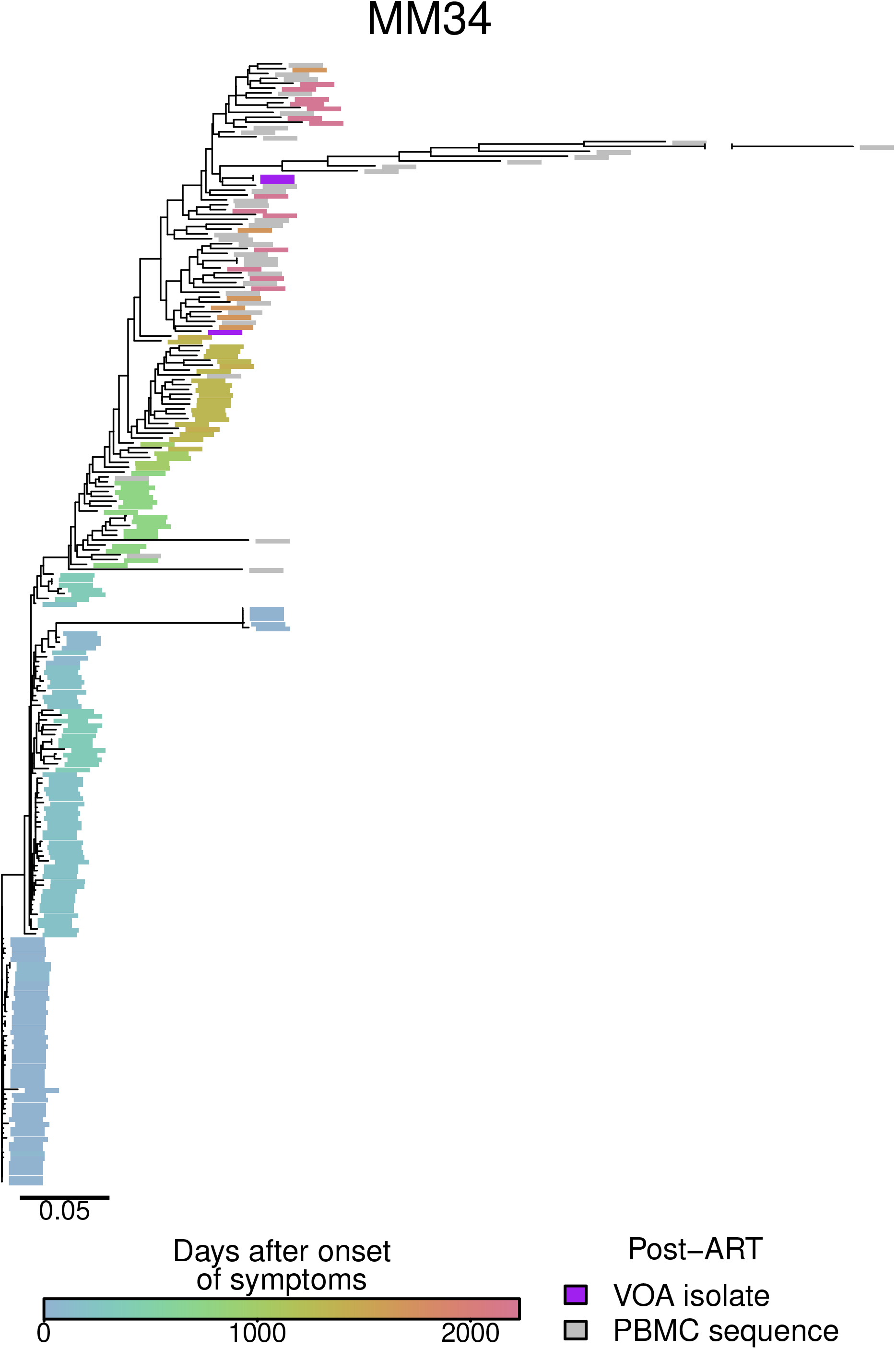
Position of QVOA isolates within the evolving HIV-1 quasispecies. The evolutionary relationships of *env* nucleotide sequences generated by SGA either directly from plasma viral RNA or plasma viral isolates of participant MM34 are shown for a 6-year time period. Samples are colored by time point, with blue sequences derived early and red sequences derived late in infection. Purple leaves indicate the position of QVOA isolates obtained 2 and 3 years following ART initiation, while grey leaves indicate proviral sequences amplified from the corresponding PBMC sample (more detailed versions of this and phylogenetic trees from other participants are shown in fig. S2). One hypermutated PBMC-derived sequence is shown with a gap in the branch. The scale bar indicates 0.05 substitutions per site.

To increase the number of outgrowth viruses for IFN-I phenotypic analyses, we obtained cryopreserved PBMCs or previously generated QVOA isolates from 9 additional ART suppressed individuals who participated in different HIV-1 treatment interruption and/or latency studies (table S3). Two individuals were leukapheresed after prolonged ART suppression for a qualitative and quantitative analysis of their replication competent HIV-1 reservoir (*61*); four individuals were leukapheresed before (−2 weeks) and during (12 weeks) ATI, but prior to viral rebound while receiving the two human broadly neutralizing antibodies 3BNC117 and 10-1074 (*62*); and three individuals were leukapheresed before and six months after ATI and infusion of the broadly neutralizing antibody VRC01 (*63, 64*). From the available material, we were able to generate or expand 52 QVOA isolates and test their IFN-I sensitivity (table S4). The results showed a range of IFNα2 and IFNβ IC_50_ values that was very similar to that of the eight QVOA isolates from the longitudinally sampled cohort, with most isolates exhibiting low or moderate IFN-I resistance (Fig. 4, A and C). There were only two outgrowth viruses that displayed an unusually high IFN-I resistance, especially for IFNα2, both of which were isolated from the post-ATI leukapheresis sample of a single individual (A08).

**Fig. 4.**
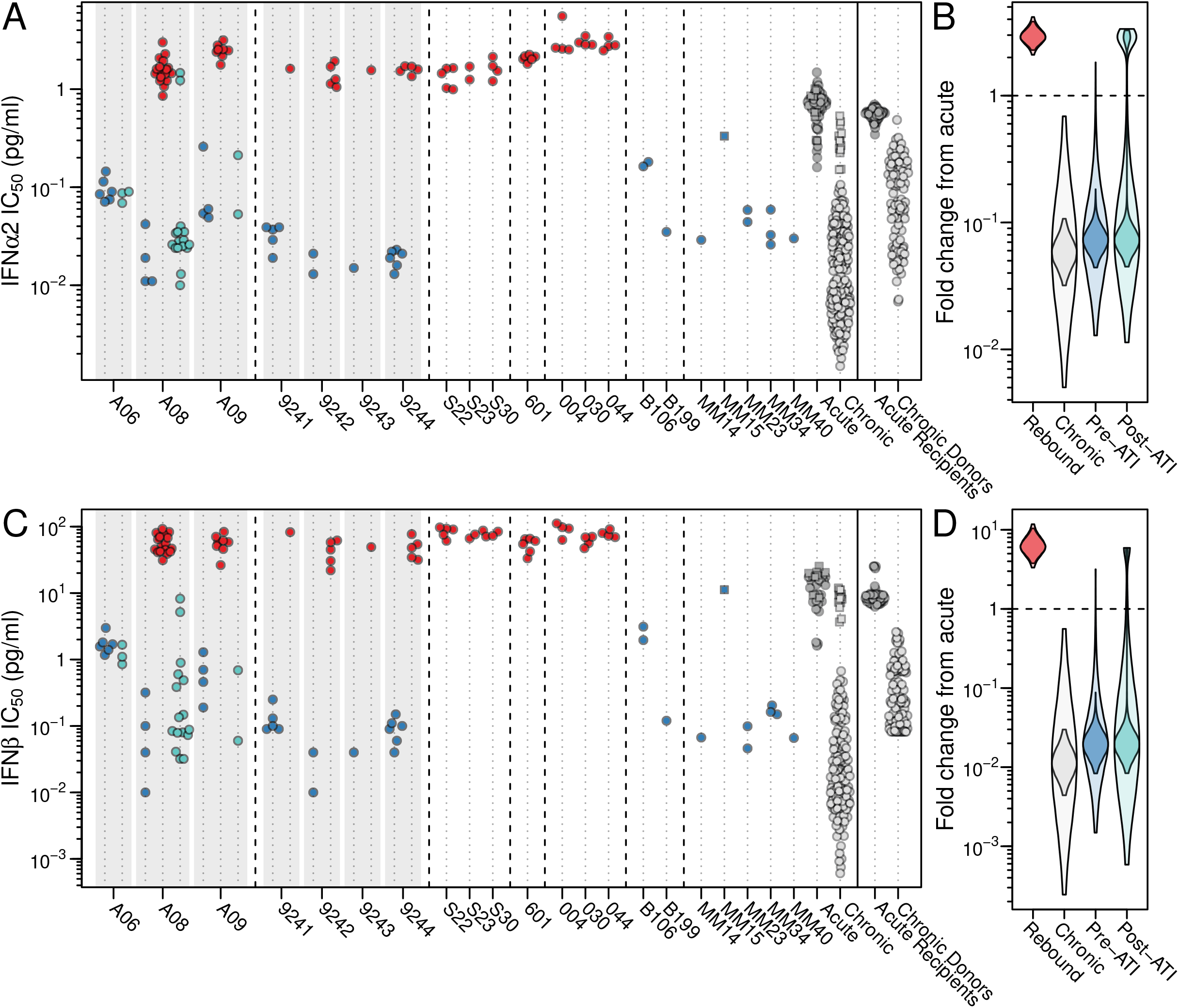
IFN-I resistance of QVOA versus rebound viruses. (A, C) IFNα2 (A) and IFNβ (C) IC_50_ values are shown for plasma isolates of individuals experiencing rebound viremia after ART cessation (red) and QVOA isolates generated from the PBMCs of ART suppressed individuals before (blue) or after (turquoise) ATI. Isolates are grouped by individuals (shaded boxes), with pre-ATI, rebound, and post-ATI isolates depicted in temporal order when available (see table S4 for details). Also shown are the IFNα2 (A) and IFNβ (C) IC_50_ values for outgrowth isolates (blue) and plasma viruses (grey) from the longitudinal cohort (Fig. 2) as well as previously reported donor-recipient transmission pairs *(50)*. For the latter, IC_50_ values were adjusted to account for potency differences among commercial IFN-I batches. Squares indicate isolates from fast progressing individuals. (B,D) The fold-change in IFNα2 and IFNβ IC_50_ values from acute infection isolates is shown for rebound (red), chronic (grey), and pre-ATI (blue) and post-ATI (turquoise) QVOA isolates (see Supplementary Methods for a description of the Bayesian model). Violin plots indicate the estimated posterior probability of the mean fold change between isolate types, with the darker shading indicating the 95% credible interval and the lighter the 95% prediction interval. Dashed horizontal line indicates a fold change of 1 (which indicates no change).

### IFN-I sensitivity of rebound viruses following treatment interruption

Some of the individuals for whom QVOA isolates were available also underwent treatment interruptions, which allowed the comparison of the IFN-I sensitivity of *in vitro* and *in vivo* reactivated viruses. Using plasma samples collected shortly after detection of recrudescent viremia, we generated 37 limiting dilution isolates from six such individuals (table S3). In addition, we obtained plasma samples from seven individuals who participated in other treatment interruption trials (table S3). Three of these underwent successive ATI cycles of increasing duration, but without further intervention (*65*); one individual was infused with the broadly neutralizing human monoclonal antibody 3BNC117 prior to and during ATI (*66*); and three individuals received a 20 week course of pegylated IFNα2 (1μg/kg) prior to and during ART interruption (*67*). Again, using samples collected shortly after detectable viremia, we were able to generate 29 additional limiting dilution isolates (table S3). All rebound isolates formed individual-specific clusters in a phylogenetic tree (fig. S6) and grew efficiently in CD4+ T cells (table S4). However, analysis of their IFNα2 and IFNβ IC_50_ values yielded unexpected results: All 66 rebound isolates were highly IFN-I resistant, exceeding even highly resistant isolates from acute infection (Fig. 4).

To quantify differences in IFN-I sensitivity between viral groups, we developed a hierarchical Bayesian model (Supplementary Methods), which combined the data from the QVOA and rebound isolates with results from the longitudinal cohort and previously published transmission pairs (50). Specifically, we compared IFN-I IC_50_ values of acute infection isolates (<30 days after onset of symptoms) with those from rebound, chronic (>300 days after onset of symptoms) and outgrowth viruses, dividing the latter into pre- and post-ATI isolates (Fig. 4, B and D). Two apparent outliers isolated from the CD4+ T cells of participant A08 six months after re-initiation of therapy suggested that viruses phenotypically resembling rebound viruses might be present in post-ATI samples (Fig. 4, A and C). To account for this possibility, we included a mixture term into the model that allowed for some proportion of post-ATI viruses to derive from the rebound population. This mixture term was also included for pre-ATI samples to allow comparison between pre- and post-ATI viruses.

Comparing all available IFN-I IC_50_ data, we determined the fold-change in IFNα2 and IFNβ IC_50_ values of acute, rebound, chronic as well as pre-ATI and post-ATI isolates (Fig. 4, B and D). Acute infection isolates from untreated individuals were on average 18-fold (95% Crl: 9.3 – 31-fold) more IFNα2 and 99-fold (33 – 230-fold) more IFNβ resistant than chronic isolates. This was also the case for pre-ATI QVOA isolates, which exhibited 13-fold (7.5 – 21-fold) lower IFNα2 and 50-fold (19 – 107-fold) lower IFNβ IC_50_ values than acute infection isolates. Remarkably, rebound viruses were the most IFN-I resistant, exhibiting on average 3.0-fold (2.3 - 3.8-fold) and 6.4-fold (3.8 - 10-fold) higher IFNα2 and IFNβ IC_50_ values than acute infection isolates, respectively. These differences were highly significant and observed regardless of whether ART interruption was combined with additional interventions, such as the administration of broad neutralizing antibodies. Interestingly, the IFNα2 IC_50_ values of rebound isolates from individuals who were treated with pegylated IFNα2 (1μg/kg) before and during ART interruption (subjects 004, 030, 044) were on average 1.8-fold (1.2 - 2.6-fold) higher than those of rebound viruses from IFNα2 untreated individuals, while no significant differences were observed for the corresponding IFNβ IC_50_ values. The fact that IFNα2 administration had a measurable effect suggests that the *in vivo* concentrations of IFNα2 were not saturating. However, the increase in viral IFNα2 resistance resulting from exogenous administration was modest compared to that driven by endogenous IFNα2 and lacked a concomitant IFNβ response. Taken together, it seems clear that treatment interruption and viral recrudescence trigger a vigorous IFN-I response that places near-maximal pressures on the rebounding virus.

### IFN-I sensitivity of post-ATI outgrowth viruses

Given the heightened IFN-I resistance of rebound viruses, we examined post-ATI isolates for an enrichment of such viruses. Post-ATI outgrowth viruses had mean IFN-I resistance values that were very similar to those of pre-ATI viruses, but appeared to comprise two populations (Fig. 4, B and D), primarily because of two post-ATI outgrowth viruses from one individual (A08) that displayed an unusually high IFN-I resistance. The model estimated the proportion of “rebound-like” viruses in post-ATI isolates to be 13% (2.9 - 29%) for IFNα2 and 4% (0.1% - 15%) for IFNβ. While no “rebound-like” highly resistant viruses were detected in pre-ATI samples, this group had large credible intervals that overlapped those estimated for post-ATI samples. Thus, existing data are insufficient to determine whether “rebound-like” isolates are exclusively observed in post-ATI samples.

To further characterize the two post-ATI QVOA isolates with elevated IC_50_ values (Fig. 4), we compared their sequences to those of other QVOA and rebound viruses from the same individual (Fig. 5). As previously reported (*63, 64*), participant A08 initiated ART during chronic infection, underwent treatment interruption and VRC01 monoclonal antibody treatment, and was restarted on ART after detection of rebound viremia. Phylogenetic analysis of viral sequences obtained before, during and after treatment interruption identified multiple rebound lineages, some of which were closely related to both pre-ATI and post-ATI QVOA isolates (Fig. 5). When 20 of these QVOA isolates were phenotypically tested, 18 exhibited very low IFNα2 and IFNβ IC_50_ values, indicating that they were highly IFN-I sensitive (Fig. 4). The exception were the two post-ATI QVOA isolates (3D8 and 6F6), which were not only more IFN-I resistant, but also shared identical *env* sequences with three rebound viruses (highlighted in Fig. 5). Further sequencing of the entire genome showed that the two QVOA isolates differed from the closest rebound virus (2F4) by only three nucleotides, which conferred single amino acid changes in Pol and Vpr, and a nucleotide substitution in the LTR (table S5). These genetic and phenotypic relationships suggest that treatment interruption in participant A08 may have re-seeded the reservoir with IFN-I resistant viruses.

**Fig. 5.**
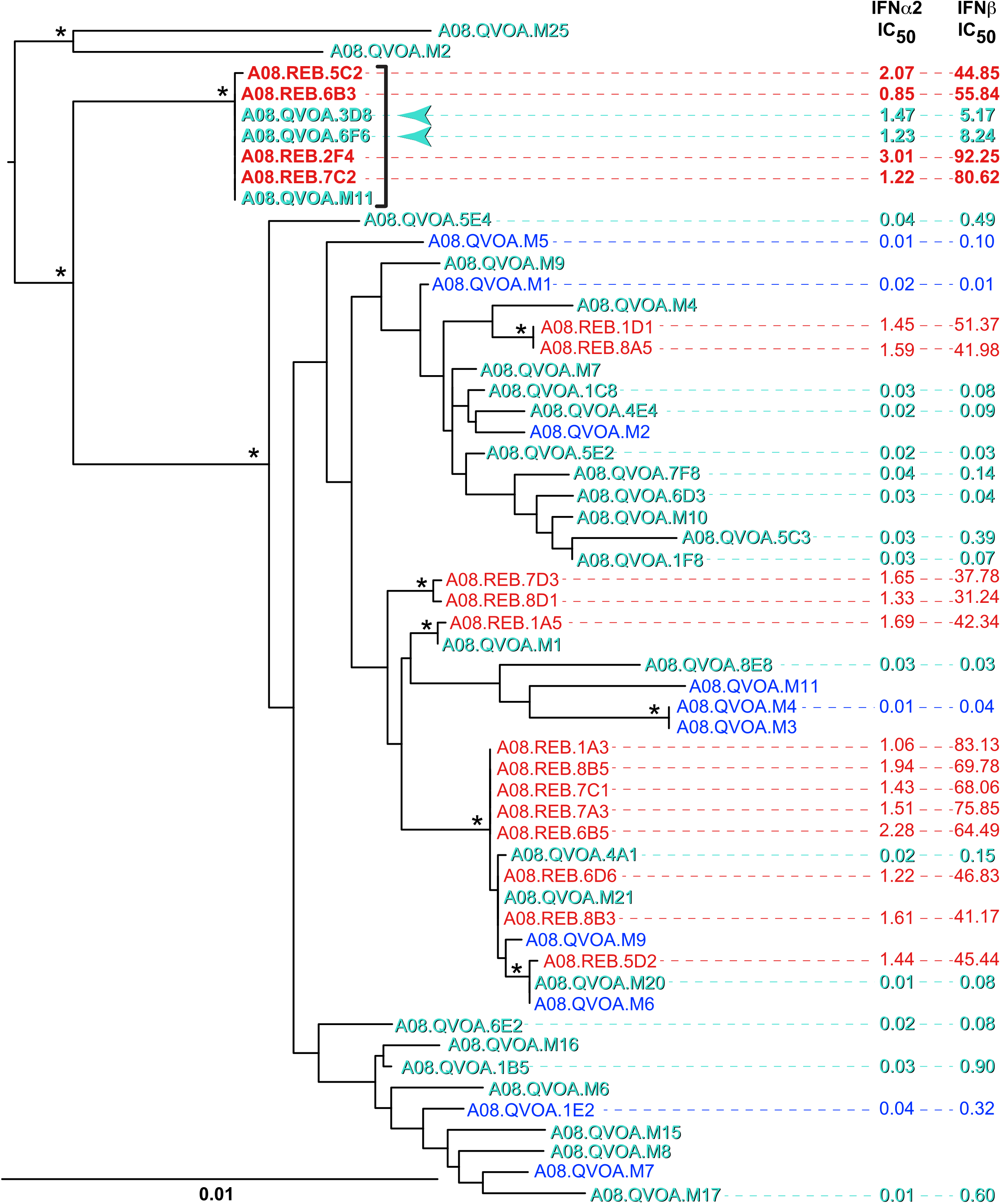
Genotype and IFN-I phenotype of rebound and outgrowth viruses before and after treatment interruption in a single individual. The phylogenetic relationships of *env* gene sequences from pre-ATI QVOA (blue), plasma rebound (red) and post-ATI QVOA (turquoise) isolates are shown for participant A08, along with available IFNα2 and IFNβ IC_50_ values (pg/ml). Asterisks indicate bootstrap values >90%; the scale bar indicates 0.01 substitution per site. A clade of near identical rebound and QVOA isolates is highlighted by a bracket, with the two post-ATI QVOA isolates with elevated IFN-I resistance denoted by arrows.

## Discussion

In untreated HIV-1 infected individuals, plasma virus has a half-life of less than one hour and the cells that are producing most of this virus have a half-life of less than one day *(43, 47, 48, 68)*. Thus, virus circulating in the plasma of infected individuals is a sensitive real time indicator of the *in vivo* selection pressures that act on virus and virally infected cells, such as neutralizing antibodies (Nab), cytotoxic T lymphocytes (CTL), and antiviral drugs (*43-46*). Here, we characterized the resistance of plasma viruses to IFNα2 and IFNβ in prospectively sampled individuals and show that the relative contribution of these cytokines to HIV-1 control varies markedly throughout the infection course (Fig. 2). IFNα2 and IFNβ IC_50_ values were uniformly high during acute infection, consistent with the rapid induction of a potent innate antiviral state that selects for founder viruses that are highly IFN-I resistant *(18, 50)*. IFNα2 and IFNβ IC_50_ values declined in all individuals in the first year post infection (Fig. 2), at least in part because of viral escape from adaptive immune responses *(45, 46, 69, 70)*. Recent studies showed that HIV-1 resistance to interferon-induced transmembrane proteins (IFITMs), and IFN-I itself, decreased over the first 6 months of infection as a direct result of acquiring neutralizing antibody (Nab) escape mutations (*69*). Although a similar causal relationship has not yet been shown for escape from cellular immune pressures, CTL responses are known to place strong pressure on the viral quasispecies during early HIV-1 infection and rapidly select for escape variants *(45, 46)*. Thus, some of the same mutations that allow HIV-1 to successfully evade potent Nab, CTL and other adaptive immune pressures likely result in a loss of IFN-I resistance, even if IFN-I signaling is maintained at a higher than normal level (26, *32, 71, 72)*. IFN-I resistance is re-acquired during later stages of infection due to a decline in cellular immunity and increasing IFN-I production, fueled in part by microbial translocation in the gut (29, 73). Thus, there is a dynamic interplay between innate and adaptive immune pressures, with the circulating virus reflecting the predominant selective forces at any given time.

Although IFN-I IC_50_ values varied by orders of magnitude, the observed temporal changes were surprisingly similar across all individuals (Fig. 2). Plasma viruses from all typical progressors displayed the same initial decline of IFNα2 and IFNβ IC_50_ values irrespective of the multiplicity of infection (number of TF viruses), followed by the gradual reacquisition of IFN-I resistance during later stages of infection (Fig. 2A). In most individuals, a nadir was reached for both IFNα2 and IFNβ within two years of symptom onset, with a large overlap in credible intervals suggesting that the respective times were not significantly different. Surprisingly, superinfection did not change this trajectory, although minor differences in IC_50_ values could have been missed due to infrequent sampling. A similar course was also observed in the two rapid progressors, although the fall and rise of IFN-I IC_50_ values was much less pronounced (Fig. 2C). Plasma viruses from the two non-progressors displayed the same initial IC_50_ decline, but then maintained low resistance values (Fig. 2B). These varying kinetics are most likely explained by differences in the quality, magnitude and duration of the adaptive immune response. In the two rapid progressors, the initial reduction in IFN-I resistance was likely curtailed by rapidly declining adaptive responses *(43, 45)*. In the non-progressors, IFN-I resistance remained low after the initial decline because adaptive responses maintained effective anti-viral control (*74*). Thus, despite differences in disease progression, similar dynamics in IFN-I resistance suggest common patterns of virus-host interactions.

The balance between innate and adaptive immune pressures is also reflected in the IFN-I phenotype of outgrowth and rebound viruses. The latent reservoir is largely comprised of proviruses that are transcriptionally and translationally silent and are therefore not subject to ongoing innate or adaptive immune pressures. Moreover, adaptive immune responses are reduced under suppressive ART due to reduced viremia and may require time for reactivation after treatment interruption *(75, 76)*. This likely explains the importance of rapidly induced innate responses at the time of viral recrudescence (*77*) and the resulting striking IFN-I resistance of rebound viruses (Fig. 4). The fact that rebound viruses were uniformly IFN-I resistant, irrespective of the clinical trial or type of treatment interruption, supports this hypothesis (*38, 62, 64-66*). Whether the superior IFN-I resistance of rebound viruses confers enhanced transmissibility is not known, but the occurrence of unintended transmissions during treatment interruption suggests that this possibility needs to be investigated *(78, 79)*.

Recent studies by Abrahams and colleagues showed that 71% of outgrowth viruses are genetically most similar to viruses replicating in the plasma just prior to ART initiation (*80*). Since this proportion is far greater than would be expected if the reservoir formed continuously, treatment appears to alter the host environment to facilitate the formation of latently infected cells (*80*). By characterizing the pathways of viral evolution in the longitudinally studied participants, we also found that outgrowth viruses isolated years after ART suppression were genetically most similar to viruses replicating late in untreated infection (Fig. 3 and fig. S2). Moreover, these outgrowth viruses exhibited IFN-I IC_50_ values that were near identical to those of viruses circulating in the plasma just prior to ART (fig. S2). Our data thus confirm and extend these earlier findings, showing that outgrowth viruses are not only genetically, but also phenotypically, most similar to viruses replicating late in infection and provide additional evidence for the preferential entry of HIV-1 into the latent reservoir near the time of ART initiation.

The viral determinants that confer IFN-I resistance during HIV-1 infection are not known. Studying infectious molecular clones of transmitted founder and six-month consensus viruses from the same individual, a previous study failed to identify resistance or sensitivity signatures that were common among the different virus pairs (*59*). Sequence changes occurred at many sites throughout the viral genome, suggesting differential responses to numerous ISGs. Given the myriad of IFN-I effector pathways and the plasticity of the HIV-1 quasispecies, these results are not surprising. Moreover, IFN-I resistance levels can be altered by a small number of sequence changes (Fig. 5). Thus, it is unlikely that a set of universal IFN-I resistance determinants will be identified. Instead, the path to resistance for any one HIV-1 strain is likely context-dependent and driven by the particular viral and immunological circumstances.

A critical question in cure research is whether treatment interruption and the associated transient viremia have an impact on the composition and size of the latent reservoir. Comparing pre-ATI and post-ATI QVOA isolates, a previous study found only minimal changes in the abundance of replication-competent viruses, with little genetic evidence of rebound viruses significantly enriching the post-ATI reservoir (*64*). The exception was one individual (A08), who harbored a subset of post-ATI viruses that were genetically closely related to rebound viruses (*64*). Examining existing and newly derived QVOA isolates from this same individual, we found that pre-ATI and post-ATI viruses had largely similar IFN-I resistance values (Fig. 4). However, the post-ATI QVOAs included two isolates with elevated IFN-I resistance, which were genetically very similar to a subset of rebound viruses (highlighted in Fig. 5). Thus, the post-ATI outgrowth viruses in this individual comprised two viral populations, one of which represented “rebound-like” viruses based on genetic and phenotypic analyses (Fig. 4, B and D and Fig. 5).

Treatment interruptions are the gold standard to evaluate promising cure strategies (*81*) and are employed with increasing frequency. Thus, it is essential that they are safe and well designed. Consistent with results from previous studies (*64, 66*), we found that pre-ATI QVOA isolates were neither predictive nor representative of the viruses that subsequently rebounded *in vivo*. This disconnect reinforces the clinical importance of ATIs since merely testing cure strategies *in vitro* on reactivated latent viruses may generate misleading results. A comprehensive analysis of post-ATI isolates in participant A08 suggested that treatment interruption may have re-seeded the reservoir with IFN-I resistant viruses, although the existence of these variants prior to ART could not be formally excluded. Although rebound-like (IFN-resistant) viruses comprised only a minor subset of the sampled post-ATI reservoir, this possibility requires further exploration in additional participants and cohorts to assure the safety of ATIs.

The finding that rebound viruses are uniformly IFN-I resistant is also relevant to the design of cure strategies that directly or indirectly engage IFN-I pathways. These approaches attempt to harness the antiviral activity of IFN-Is either by administering these cytokines directly (*38*) or by eliciting IFN-I responses via immunomodulators, such as TLR 7 and 9 agonists (*82, 83*), and other latency reversal agents (*84*). The fact that ART cessation likely induces near-maximal IFN-I responses at the sites of virus recrudescence will need to be factored into the design of these strategies. Consistent with this, a recent study found that activation of NK cell cytotoxicity, and not the induction of interferon stimulated genes, correlated with exogenous IFNα2-mediated HIV-1 control (*85*).

A limitation of the present study is the lack of diversity among the study participants, all but one of them were men-who-have-sex-with-men from the US and western Europe. This is a longstanding issue in translational HIV-1 research that will have to be addressed by enrolling trial participants who are more representative of the gender, ethnicity, co-morbidity, disease progression and viral subtype composition of the HIV-1 pandemic. Our study also focused on INFα2 and IFNβ, because the potency of these cytokines has previously been demonstrated both *in vitro (50)* and *in vivo* (29). However, additional IFN-I subtypes will need to be analyzed to elucidate the full spectrum of relevant innate immune pressures on HIV-1. It will also be important to perform viral outgrowth assays in the presence and absence of IFN-I to see whether pre-treatment of latently infected CD4+ T cells activates viruses that genetically or phenotypically resemble rebound viruses. Similarly, integration site analysis will be important to determine whether IFN-I resistant proviruses preferentially integrate in chromatin regions that restrict viral gene expression in the absence of high levels of IFN-I. The striking IFN-I resistance of rebound viruses also reinforces the need for nonhuman primate models that faithfully recapitulate this key feature of HIV-1 persistence. Studies are underway to determine whether SIVmac or SHIV infected rhesus macaques recapitulate the IFN-I kinetics observed in humans, and if so, whether these models can be used to trace the provenance and activation requirements of rebound viruses.

In summary, we show here that IFN-I play an important role in the control of HIV-1, not only during acute, but also later stages of infection. The IFN-I resistance of plasma viruses reflects the contribution of IFN-I mediated activity relative to other antiviral forces at the site of virus production. The kinetics of the IFN-I response vary over the course of infection, from very high levels in acute infection to low levels in early infection to elevated levels prior to ART initiation and near maximal levels during viral rebound following ART cessation. Although the tissue and cell origins of rebound viruses remain to be determined, our data indicate that they arise either from a cryptic reservoir of highly IFN-I resistant viruses or rapidly evolve at the sites of viral recrudescence by acquiring IFN-I resistance.

## Materials and Methods

### Study design

The role of IFN-Is in the control of HIV-1 during infection was determined by generating 500 plasma and PBMCs derived viral isolates from 26 individuals sampled prospectively before and after antiretroviral therapy (ART) and/or during treatment interruption. Viruses were genetically characterized and their susceptibility to IFNα2 and IFNβ inhibition (IC_50_) was determined. Sample sizes were dependent on availability and not predetermined by power calculations.

### Study participants

Participants MM14, MM15, MM23, MM33, MM34, MM39, MM40, MM55, and MM62 were recruited to an acute HIV-1 infection cohort at the Mortimer Market Centre for Sexual Health and HIV Research in London, United Kingdom, while participant WEAU was followed at the 1917 Clinic of the University of Alabama at Birmingham. Six individuals who experienced a gradual loss of their CD4+ T cells, were classified as typical progressors; two individuals who maintained high CD4+ T cell counts and low viral loads, were classified as non-progressors; and two individuals, who developed AIDS within 8 and 18 months post-infection, respectively, were classified as rapid progressors (Fig. 1). Peripheral blood was collected in EDTA-containing tubes following onset of symptoms and at regular intervals thereafter (table S1). Plasma was separated by centrifugation, while PBMCs were isolated using a Histopaque 1.077 density gradient and cryopreserved. Subjects provided informed consent and were offered antiretroviral therapy based on the standard-of-care at the time. Ethical approval for the London cohort study was provided by the National Health Service Camden/Islington Community Local Research Ethics Committee. The use of stored (de-identified) samples from participant WEAU was approved by the University of Alabama at Birmingham Institutional Review Board. Plasma and viably frozen PBMC samples were also obtained from 16 individuals who participated in six different treatment interruption and latency trials (MNU-0628, NCT02825797, NCT0246322, NCT00051818, NCT02227277, NCT02588586). These materials were obtained from existing repositories and their use was approved by the respective Institutional Review Boards as previously described *(61-67)*.

### Virus isolation from plasma

For limiting dilution isolation, plasma samples were end-point diluted and used to infect 1 × 10^6^ activated CD4+ T cells from multiple healthy donors in 24-well plates (50). To generate bulk isolates, plasma aliquots containing 1,500 - 20,000 viral RNA copies were used to infect 4 × 10^6^ healthy donor CD4+ T cells in 6-well plates. Cultures were maintained for 21 days and tested weekly for the presence of p24 antigen in the supernatant. Virus positive cultures were expanded in pooled healthy donor CD4+ T cells (1 × 10^7^) and the resulting viral stocks were used for all genetic and biological analyses.

### Quantitative viral outgrowth assay

Virus isolations were performed essentially as described (*60*) with minor modifications. CD4+ T cells were isolated from viably frozen PBMC, incubated in RPMI media containing 15% FBS without IL-2 for 24 hours, and stimulated for 24 hours using a combination of anti-CD2, CD3 and CD28 antibody-coated beads (Miltenyi Biotec T Cell Activation/Expansion Kit). Cells were then cocultured with healthy donor CD4+ T cells (1:10 patient:donor cells) and cultured for 3 weeks with weekly monitoring of p24 production. Positive cultures were expanded and viral stocks generated as described above. In cases where QVOA isolates were already available *(61-64)*, supernatants were obtained and used to generate viral stocks in healthy donor CD4+ T cells (table S3).

### Single genome amplification

Single genome amplification of 3’ half genomes or viral *env* genes from plasma RNA and proviral DNA was performed as previously described *(54, 86)*. Briefly, ~20,000 copies of viral RNA were extracted from plasma using QIAamp Viral RNA kit (Qiagen) and reverse transcribed using SuperScript III Reverse Transcriptase (Invitrogen). PBMC DNA was extracted using a DNeasy Blood and Tissue kit (Qiagen). Viral cDNA and PBMC DNA were then endpoint diluted and amplified using nested PCR using primers and conditions as previously reported *(54, 86)*.

### Isolate sequencing

Viral RNA was extracted from isolate stocks, reverse-transcribed, and the resulting cDNA used to amplify overlapping 5’ and 3’ genome halves in separate PCR reactions (*50*). Amplicons were sequenced using Illumina MiSeq and paired-end reads assembled to generate a sample-specific reference sequence. Viral reads were mapped to this reference, and the extent of genetic diversity was examined for each position along the alignment. Isolates that exhibited more than 15% diversity at any one position were judged to contain more than one variant.

### Phylogeny

Plasma viral, proviral and isolate derived sequences for each subject were aligned with Clustal Omega (*87*) and inspected for misassemblies, overly truncated sequences and sequences with abundant G to A mutations (considered to be the result of APOBEC hypermutation). Alignments were trimmed to the shortest sequence and positions with gaps in any sequence were masked in all sequences. Trees were generated using RAxML v8.2.12 (*88*) with model GTRGAMMA and bootstrap values calculated from 1,000 replicates. Potential recombinants from superinfected subjects were identified in alignments using Recco (*89*) using default parameters; recombinants were reported if they had an alignment p-value <0.05 and contained at least 10 recombination induced mutations.

### Interferon IC_50_ determinations

To determine the IFNα2 and IFNβ concentrations required to inhibit virus replication by 50% (IC_50_), pooled activated CD4+ T cells were left untreated or cultured in the presence of increasing amounts of IFNα2 or IFNβ for 24 hours, essentially as described (50). Cultures (2.5×10^5^ cells) were then infected overnight with equal amounts of virus (0.25 ng of RT) and maintained for 7 days in IL-2 (30 U/ml) media, while replenishing IFN-I every 48 hours. Virus replication was measured as the amount of p24 produced at day 7 and IC_50_ was calculated as the amount of IFN necessary to reduce viral growth by 50% from levels measured in the absence of IFN-I (fig. S3).

### Statistical analyses

We developed hierarchical Bayesian models to analyze the longitudinal changes of interferon resistance while avoiding bias from differential sampling *(50, 58)*. These models assumed that the data were log normally distributed with the resistance observed in each individual deriving from a common population-level distribution (Supplementary Methods). For the longitudinal analysis, IFN-I resistance was assumed to start at an acute infection level, fall (or rise) to some nadir change point, and then rise (or fall) based on changes in CD4+ T cell counts (fig. S4). For comparisons of outgrowth and rebound viruses, each isolate was assumed to be a random sample from the viruses circulating within an individual, and each individual was assumed to be representative of a larger population. Outgrowth isolates were assumed to represent mixed populations that could include rebound-like viruses (Supplementary Methods). Data was processed and analyzed using R v3.4.4 (90). Posterior probability distributions were estimated using Markov chain Monte Carlo sampling as implemented in Stan v2.23 (91). Statistical significance was assessed by determining if the 95% credible interval overlapped the null hypothesis.

## Data Availability

HIV-1 sequences have been deposited in GenBank as listed in table S5. Analysis code is available at https://github.com/sherrillmix/ifnDynamicsModels and will be archived on Zenodo prior to publication.

## Acknowledgments

We thank G. H. Learn for sequence analysis, S.S. Iyer for expertise in IFN-I resistance testing, K. Ruffin for preparation of the manuscript and the participants who generously volunteered for these studies.

## Author contributions

M.V.P.G., S.S.-M., F.B.-R., R.M.R., G.M.S., P.B., K.J.B. and B.H.H. conceived and planned the study; M.V.P.G., F.B.-R., S.T., A.G.S., Y.L., W.L., A.N.A., J.D., and J.C. performed sequence analyses; M.V.P.G, S.S.-M., F.B.-R., R.M.R. and S.T. isolated virus and determined IFN-I IC_50_ values; L.J.P. and P.M.S. performed evolutionary analyses; S.S.-M. performed statistical analyses; A.E.F-M., P.P., I.W., E.P., J.C.C.L., D.B.S., F. M., M.A.M., Y.Z.C., S.H., M.S.G., L.J.M., R.G.C., J.M.S., R.F.S., M.C., M.C.N., P.B., K.J.B. conducted clinical trials, provided patient material, performed serological analyses, and performed outgrowth assays; M.V.P.G., S.S.-M. P.B., K.J.B. and B.H.H. coordinated the contributions of all authors and wrote the manuscript.

## Supplementary Materials

### Supplementary Methods

**Fig. S1.**
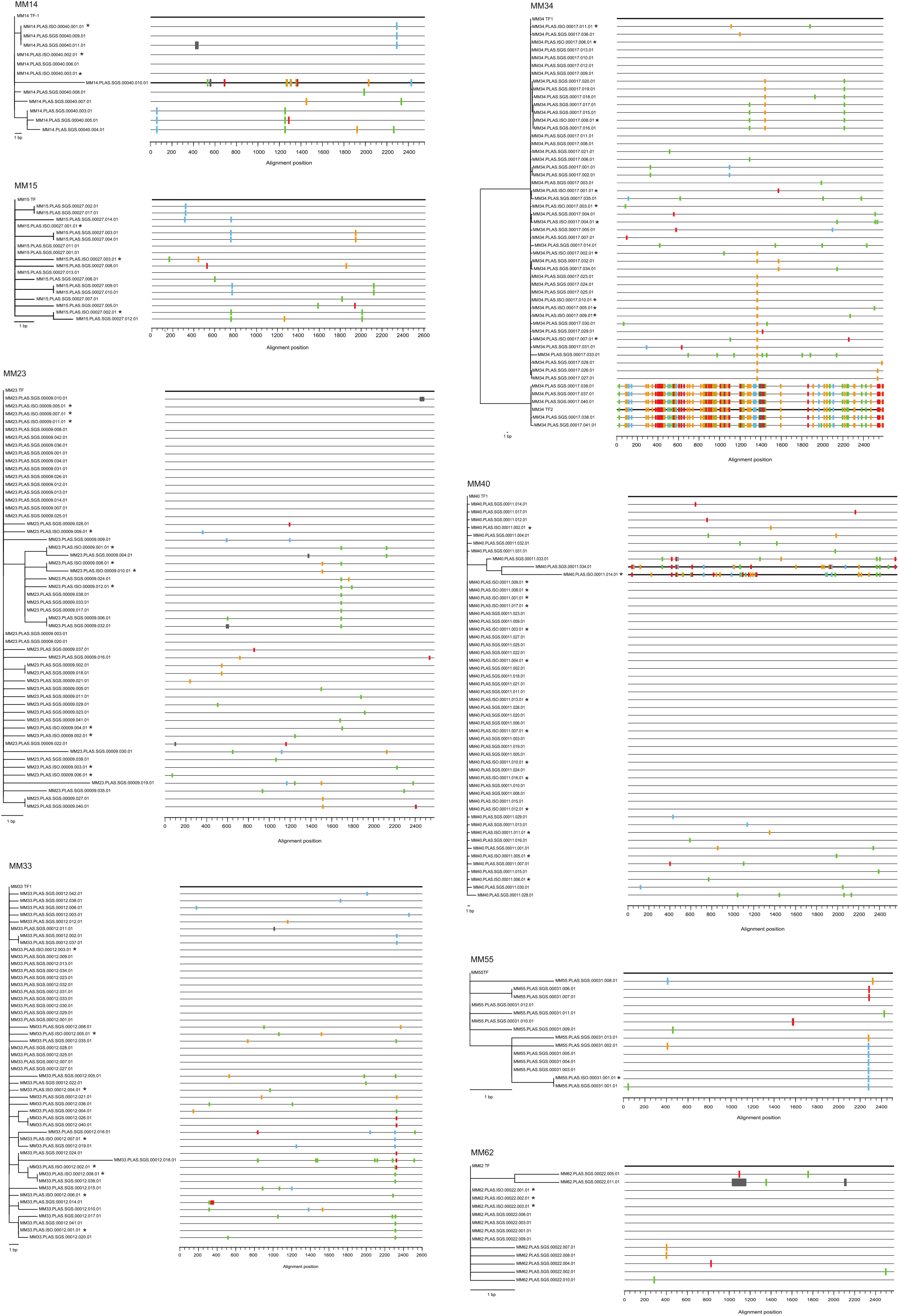
Transmitted founder virus inference and enumeration. Highlighter (v2.1.1) plots and associated phylogenetic trees of SGA and limiting dilution isolate derived *env* gene sequences are shown for the first available plasma sample of eight study participants. Tick marks indicate differences from the inferred TF sequence depicted in bold on top (red, T; green, A; blue, C; orange, G; gray, indel). Participants MM14, MM34 and MM40 were infected with more than one transmitted founder virus (highlighted in bold). Trees were constructed using the neighbor-pining method implemented in the Highlighter v2.1.1 software (hiv.lanl.gov). Sequences obtained from plasma isolates are indicated by an asterisk. The scale bar indicated 1 base pair. Participants MM39 and WEAU were each infected with a single transmitted founder virus as reported previously *(54, 57)*.

**Fig. S2.**
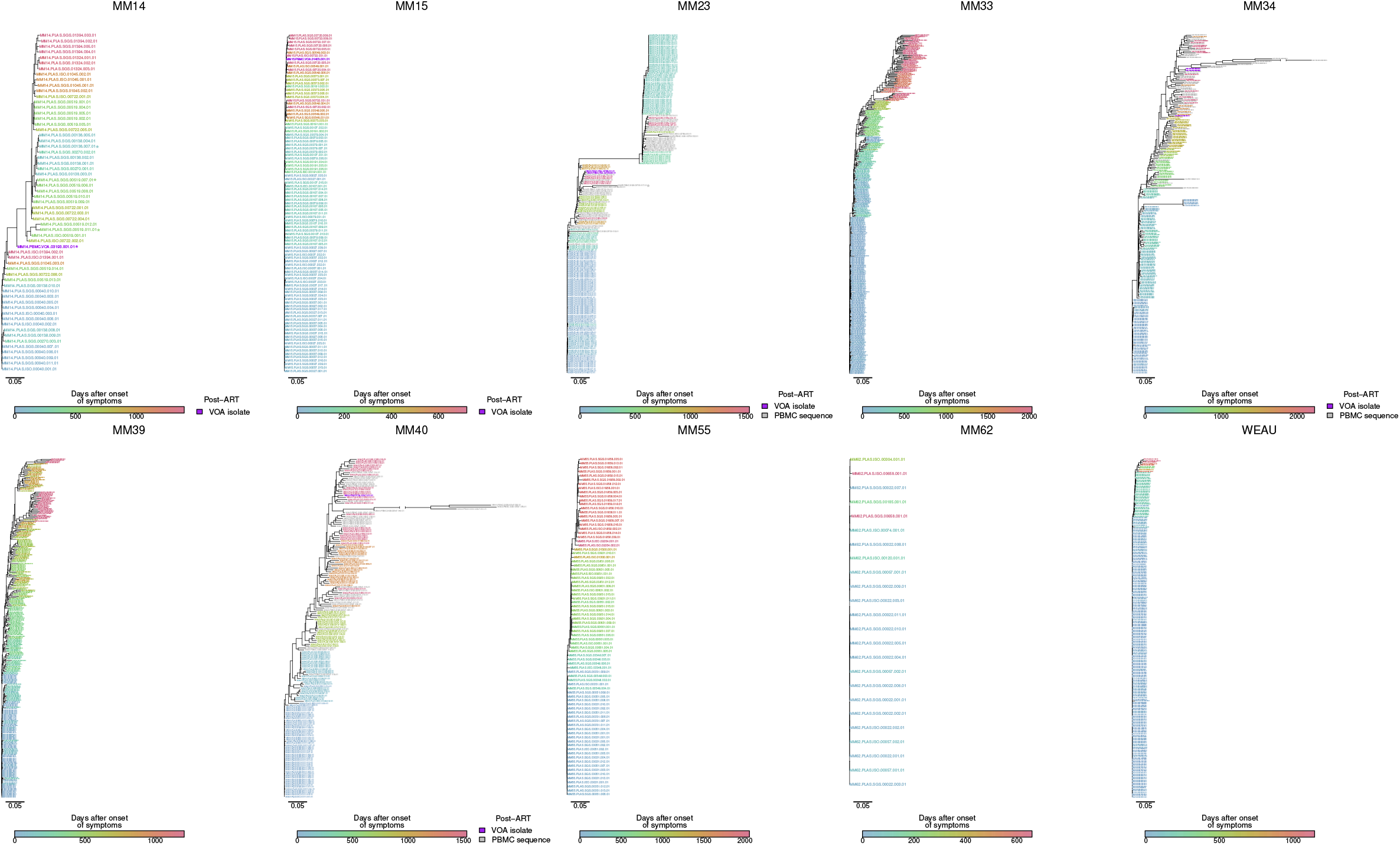
HIV-1 quasispecies diversification over time. A maximum likelihood tree depicting the phylogenetic relationships of *env* nucleotide sequences amplified either directly from uncultured plasma (PLAS.SGS) or PBMCs (PBMC.SGS), or generated from limiting dilution- derived plasma (PLAS.ISO) and PBMC (PBMC.QVOA) viral isolates, is shown for each of 10 prospectively sampled individuals. Sequences are colored by time point (indicated in the sequence name), with blue (earliest samples) transitioning to red (latest samples) as in Fig. 3. Purple leaves indicates PBMC derived outgrowth viruses obtained after years of suppressive ART, while grey leaves indicate SGA sequences amplified directly from the corresponding PBMCs. Each leaf is labeled to indicate the individual, source, days post onset of symptoms, and a sequence or isolate identifier (see tables S1 and S2 for details). Asterisks indicate recombinants. A gap in a branch accommodates hypermutated sequences. The scale bar indicates 0.05 substitutions per site.

**Fig. S3.**
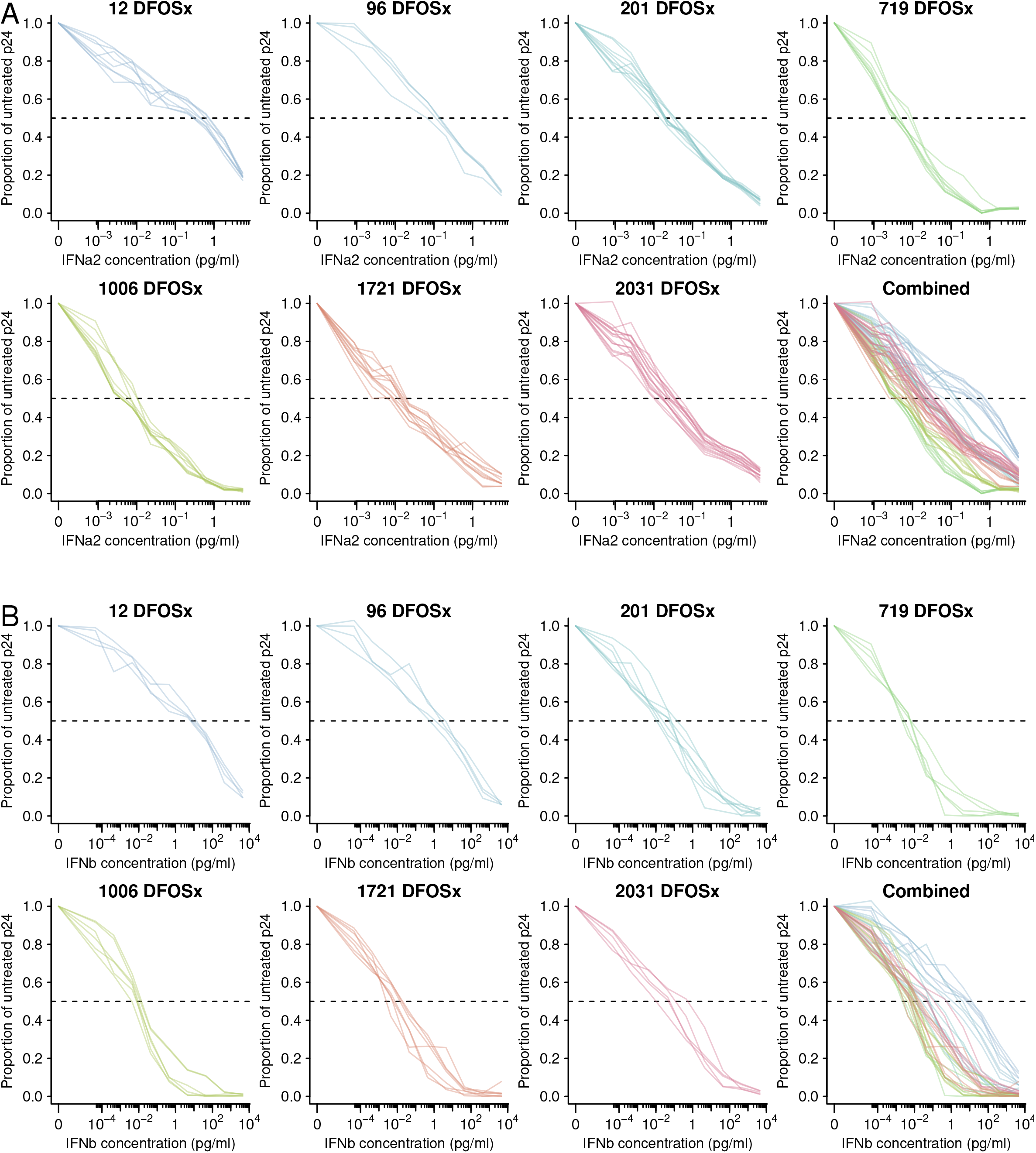
Determination of IFN-I IC_50_ values. Dose response curves showing the effect of increasing amounts of IFNα2 (A) and IFNβ (B) on the replication potential of limiting dilution plasma viral isolates from participant MM33 are shown for individual timepoints with days following onset of symptoms (DFOXs) indicated or combined (last panel). Curves are colored by timepoint. Dashed horizontal lines indicate the half-maximal inhibitory concentration (IC_50_).

**Fig. S4.**
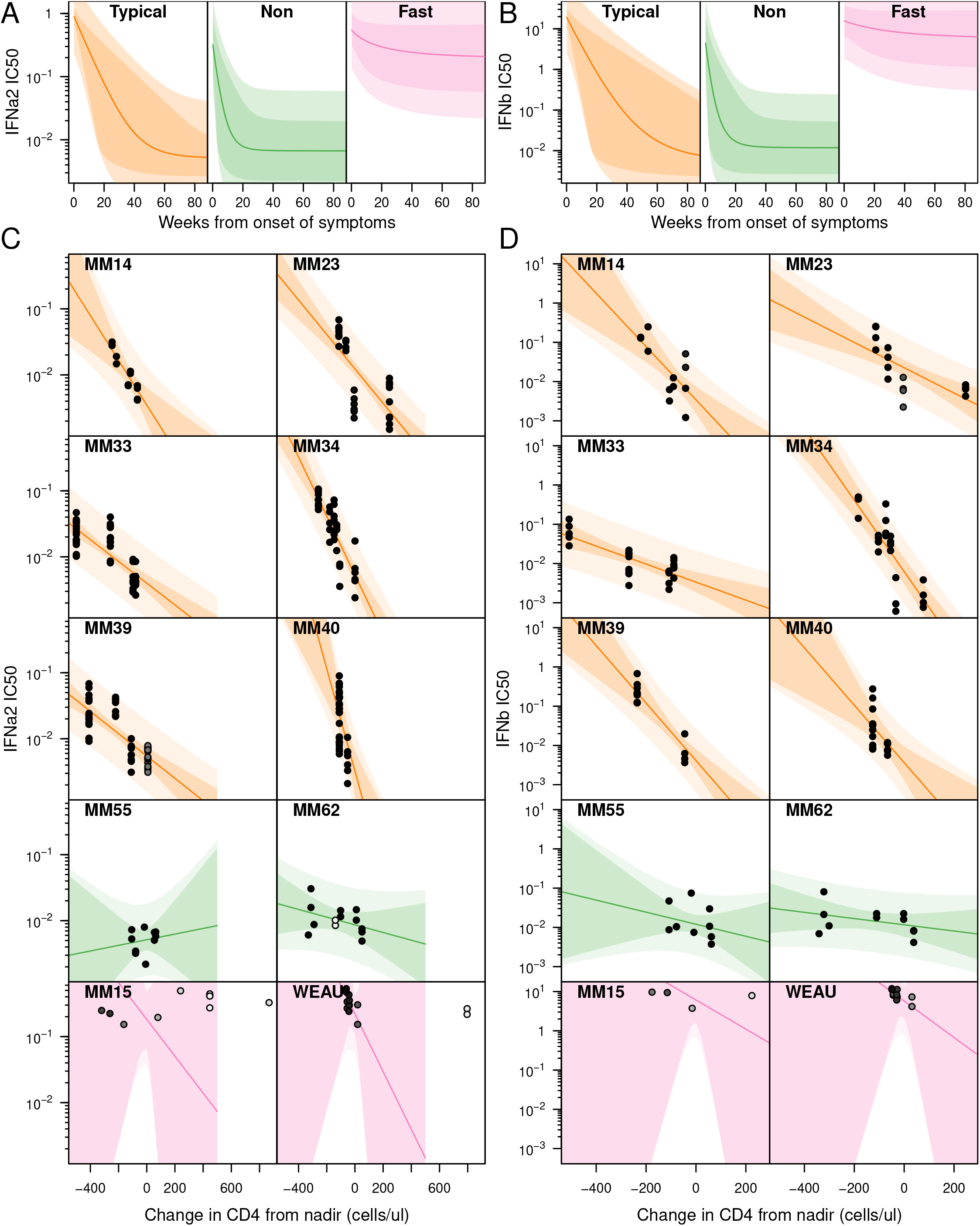
Dynamic changes in IFN-I resistance over the course of HIV-1 infection. A hierarchical Bayesian change point model was used to estimate the IFN-I resistance of plasma viral isolates in 10 longitudinally sampled HIV-1 infected study participants. The model allowed the inference of longitudinal patterns of IFNα2 and IFNβ IC_50_ values across individuals by first predicting resistance as a fall from acute levels to a nadir point and then as variation from nadir levels based on changes in CD4+ T cell counts. (A-B) Predicted mean IFNα2 (A) or IFNβ (B) IC_50_ values (lines) are shown for plasma virus isolates modelled for typical progressor (orange), non-progressor (green) and rapid progressor (pink) participants from the time of transmission over the early course of infection (the x-axis shows weeks from onset of symptoms), along with corresponding 95% credible (darker shading) and prediction (lighter shading) intervals. (C-D) Predicted mean IFNα2 (C) or IFNβ (B) IC_50_ values (lines) are shown for plasma virus isolates modeled for individual typical progressor (orange), non-progressor (green) and rapid progressor (pink) participants based on changes in CD4+ T cell counts following the nadir (the x-axis shows decreases and increases from the nadir which is set to 0), along with corresponding 95% credible (darker shading) and prediction (lighter shading) intervals. Individual data points indicate virus isolates from the respective individuals, with shading indicating the estimated posterior probability that the time of nadir preceded the plasma collection time point (from white, probability of 0, to black, probability of 1). Isolates estimated to have less than 5% probability of following the time of nadir are not shown. For display, the nadir CD4+ T cell count was estimated as the posterior mean for that individual.

**Fig. S5.**
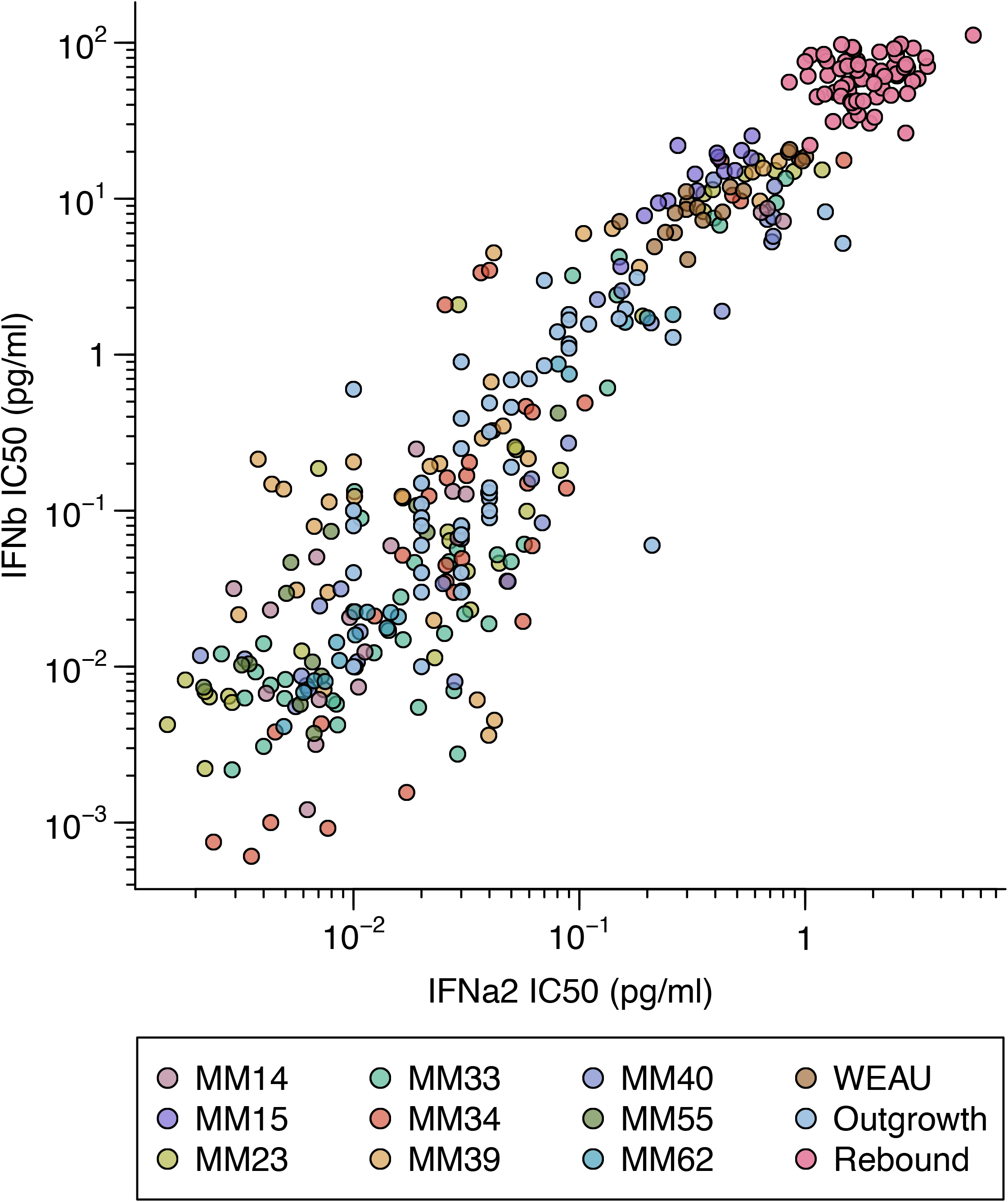
Correlation of IFNα2 and IFNβ IC_50_ values. The IFNα2 and IFNβ IC_50_ of all longitudinal, outgrowth and rebound isolates are compared. Isolates from longitudinally studied individuals as well as participants in latency and/or treatment interruption trials are represented by points colored as indicated in the legend.

**Fig. S6.**
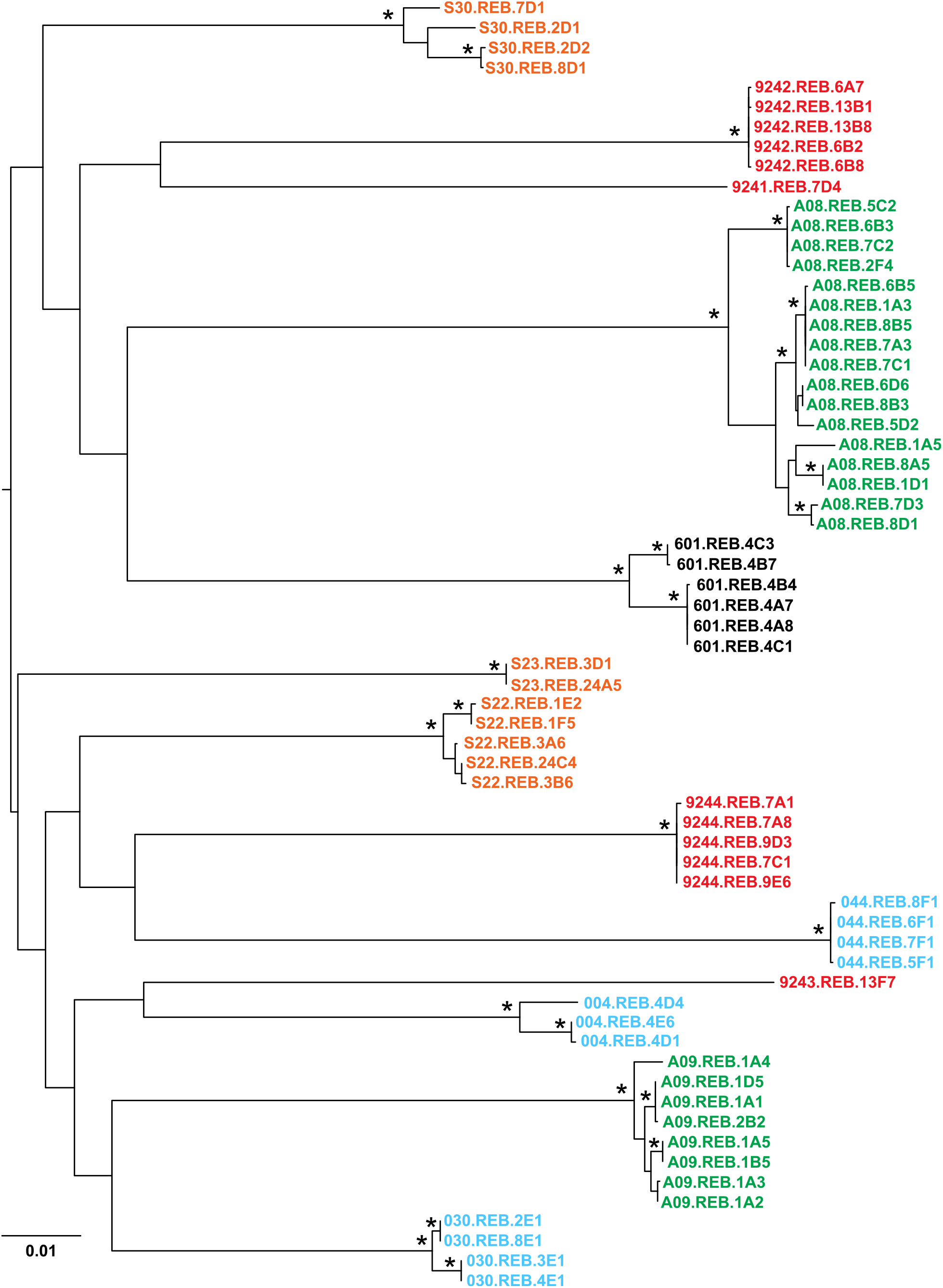
Phylogenetic relationships of rebound isolates. A maximum likelihood tree depicting the phylogenetic relationships of 3’ half-genome sequences of rebound isolates from different study participants is shown. Isolates are colored according to the respective study (red, NCT02825797; green, NCT02463227; orange, NCT00051818; turquoise, NCT02227277; black, NCT02588586) as indicated in tables S3 and S4. Asterisks indicate bootstrap values of >90%; the scale bar indicates 0.01 substitution per site. Isolate 004.REB.4E3 was not sequenced.

**Table S1.**
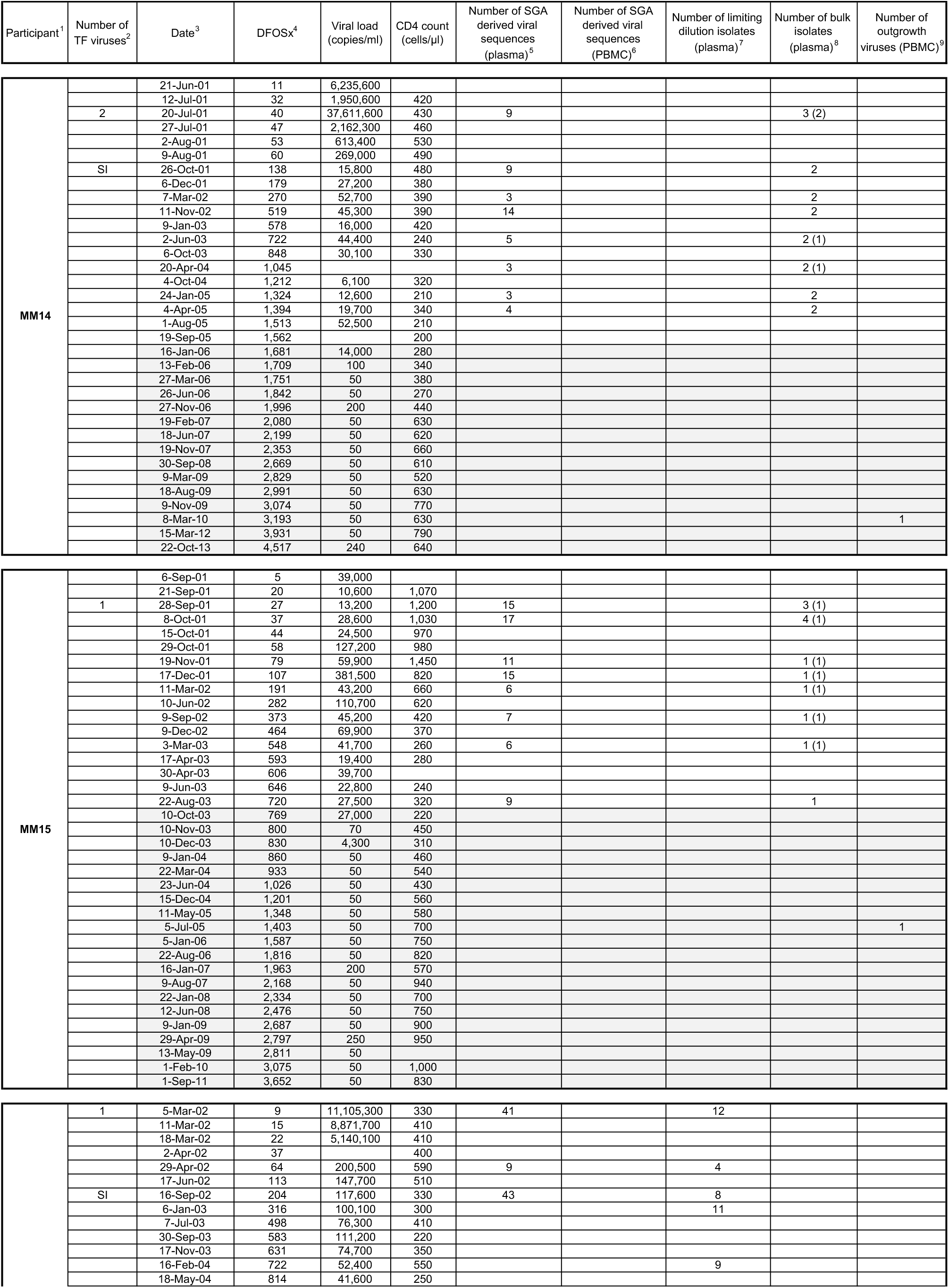
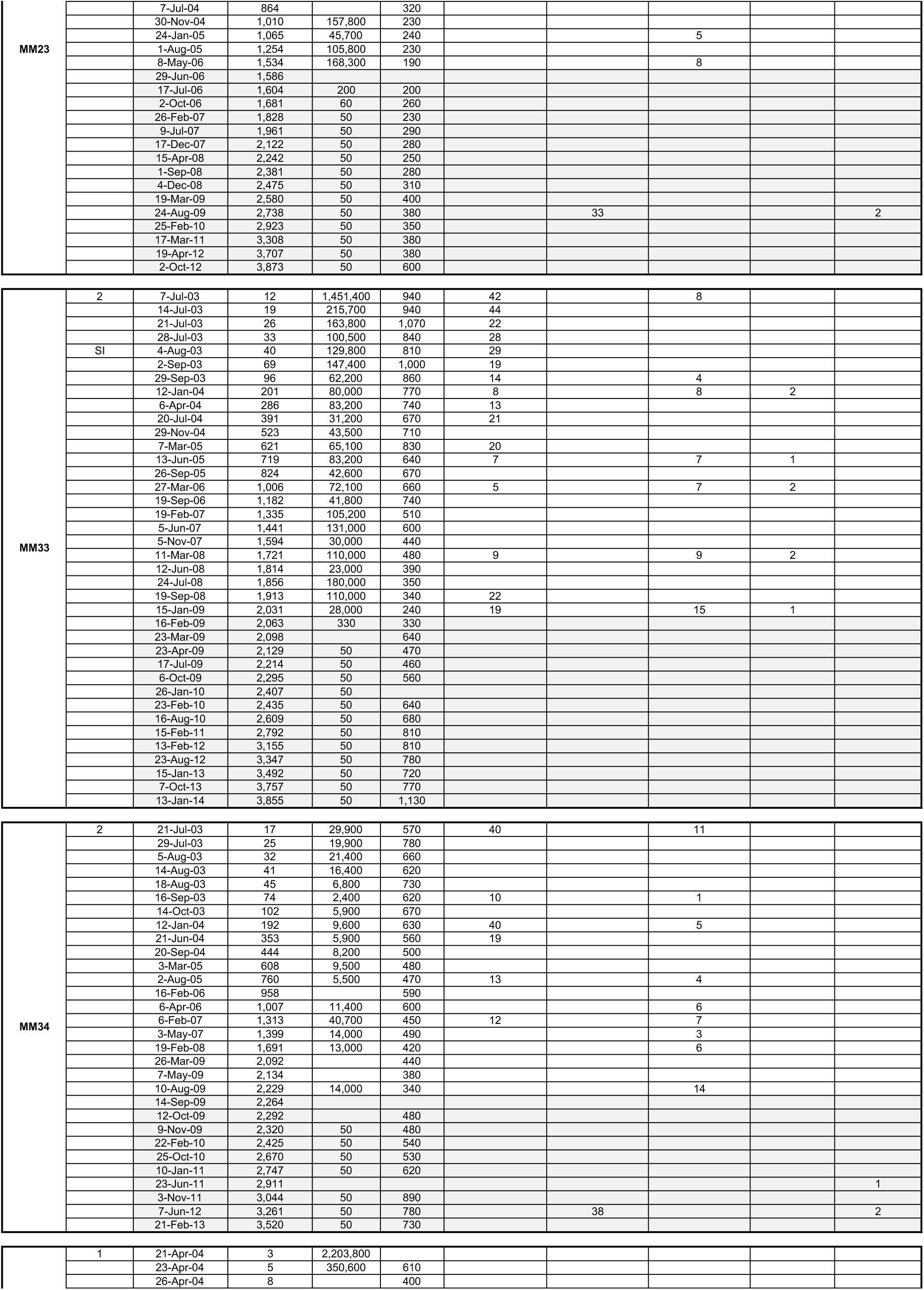
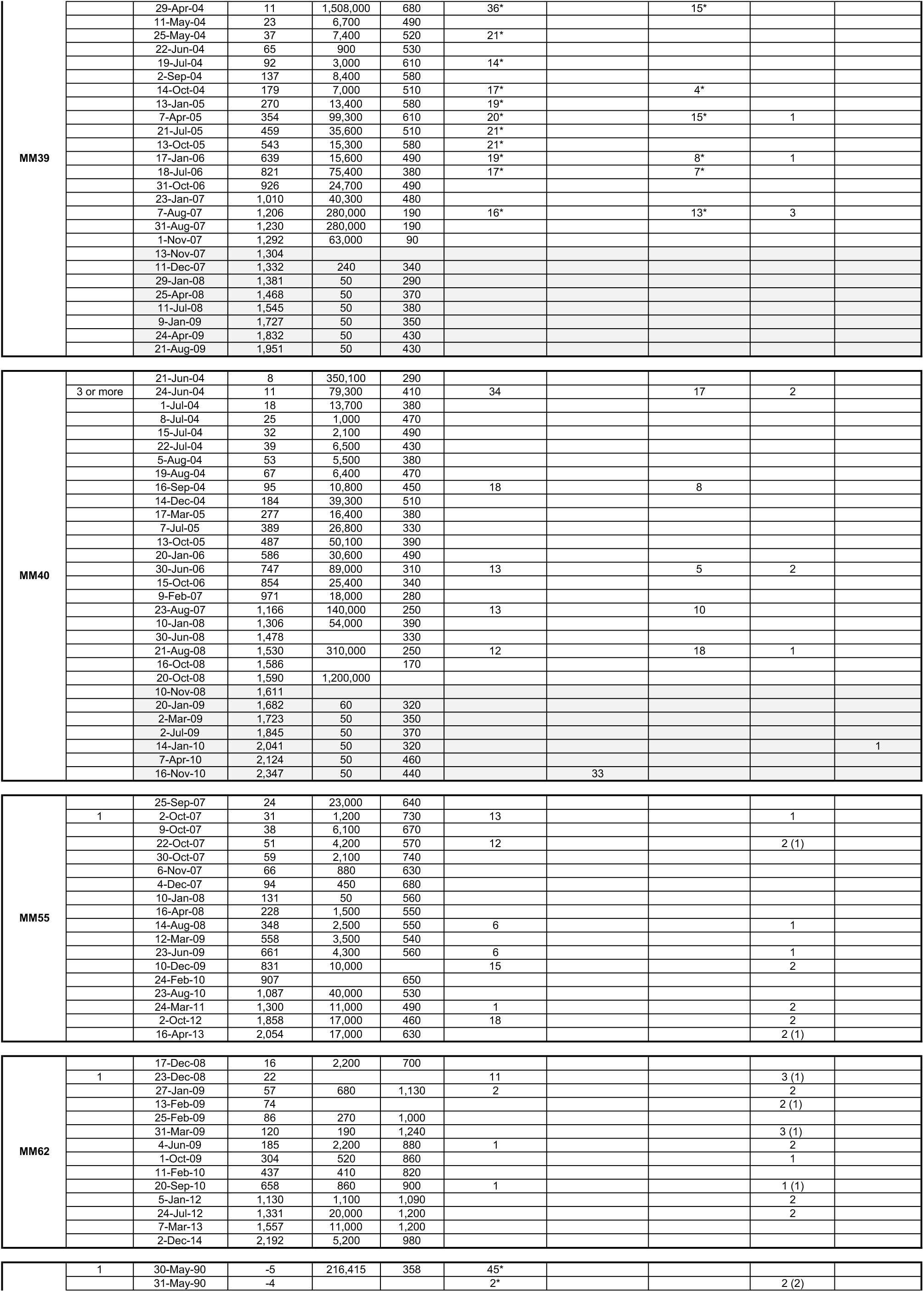
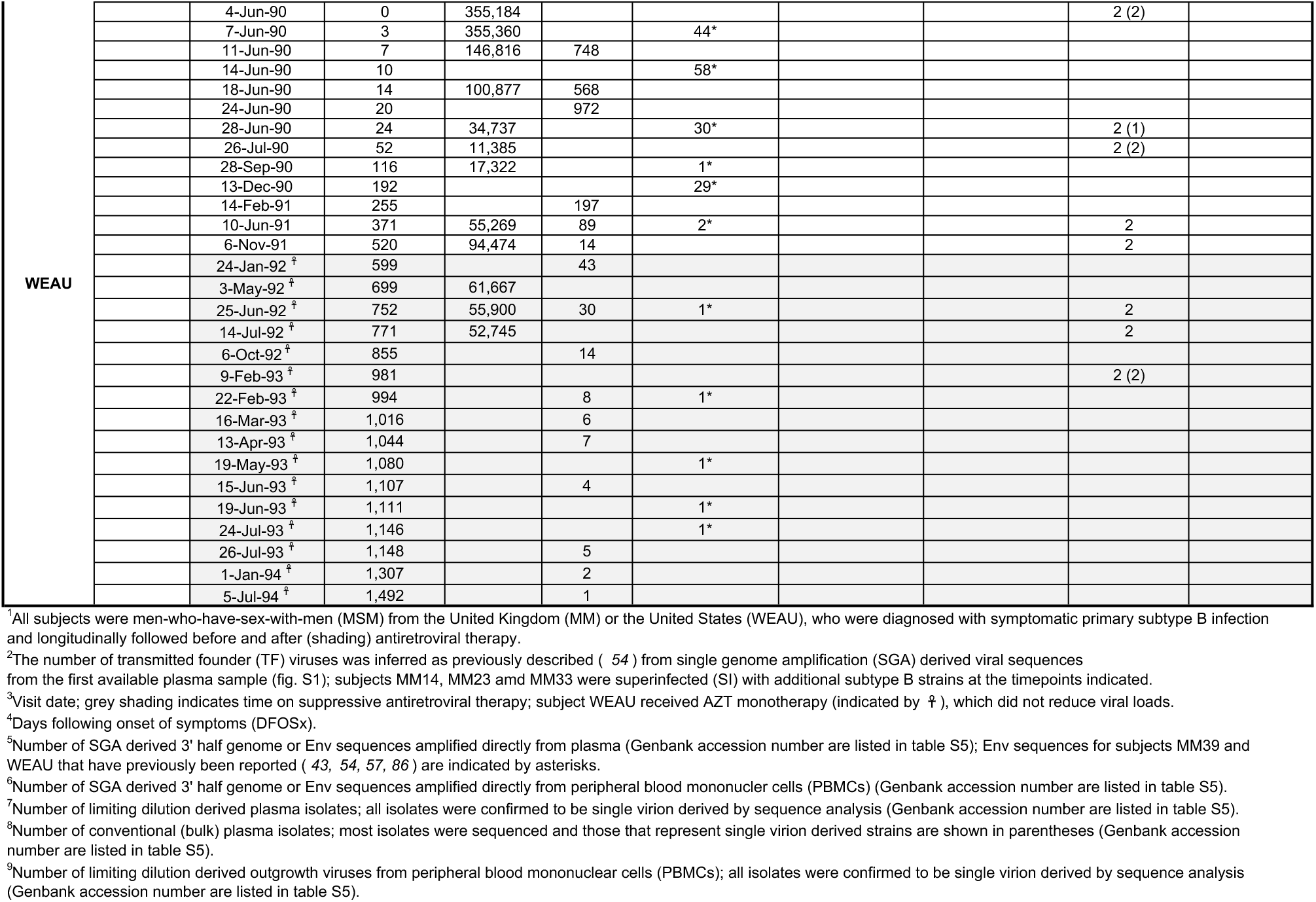
Generation of HIV-1 isolates from plasma and peripheral blood mononuclear cells of 10 individuals sampled from acute infection throughout their clinical course.

**Table S2.** IFN-I resistance of plasma and QVOA isolates from longitudinally sampled study participants.

| Isolate | Sequence ID<sup>1</sup> | DFOSx<sup>2</sup> | Replicative capacity<br>(ng p24/ml)<sup>3</sup> | IFNα2 IC50<br>(pg/ml)<sup>4</sup> | IFNβ IC50<br>(pg/ml)<sup>4</sup> |
| --- | --- | --- | --- | --- | --- |
| MM14.02.2C3.bulk | MM14.PLAS.ISO.00040.003 | 40 | 1,148 | 0.639 | 8.160 |
| MM14.02.2B4.bulk | MM14.PLAS.ISO.00040.002 | 40 | 899 | 0.682 | 8.666 |
| MM14.02.2A1.bulk | MM14.PLAS.ISO.00040.001 | 40 | 924 | 0.800 | 7.161 |
| MM14.06.2B1.bulk | MM14.PLAS.ISO.00138.001 | 138 | 525 | 0.010 | 0.021 |
| MM14.06.2B2.bulk | MM14.PLAS.ISO.00138.002 | 138 | 394 | 0.003 | 0.032 |
| MM14.08.2B3.bulk | MM14.PLAS.ISO.00270.001 | 270 | 447 | 0.007 | 0.050 |
| MM14.08.2B4.bulk | MM14.PLAS.ISO.00270.002 | 270 | 511 | 0.004 | 0.023 |
| MM14.10.2C1.bulk | MM14.PLAS.ISO.00519.001 | 519 | 449 | 0.004 | 0.007 |
| MM14.10.2C2.bulk | MM14.PLAS.ISO.00519.002 | 519 | 251 | 0.006 | 0.001 |
| MM14.12.1A1.bulk | MM14.PLAS.ISO.00722.001 | 722 | 414 | 0.015 | 0.060 |
| MM14.12.1A2.bulk | MM14.PLAS.ISO.00722.002 | 722 | 476 | 0.019<sup>5</sup> | 0.248<sup>5</sup> |
| MM14.14.1A1.bulk | MM14.PLAS.ISO.01045.001 | 1,045 | 273 | 0.007 | 0.006 |
| MM14.14.1A2.bulk | MM14.PLAS.ISO.01045.002 | 1,045 | 215 | 0.007 | 0.003 |
| MM14.16.1B1.bulk | MM14.PLAS.ISO.01324.001 | 1,324 | 688 | 0.028 | 0.133 |
| MM14.16.1B2.bulk | MM14.PLAS.ISO.01324.002 | 1,324 | 725 | 0.032<sup>5</sup> | 0.128<sup>5</sup> |
| MM14.17.1B3.bulk | MM14.PLAS.ISO.01394.001 | 1,394 | 304 | 0.011 | 0.007 |
| MM14.17.1B4.bulk | MM14.PLAS.ISO.01394.002 | 1,394 | 253 | 0.011<sup>5</sup> | 0.012<sup>5</sup> |
| MM14.23.1A3.QVOA | MM14.PBMC.QVOA.03193.001 | 3,193 | 347 | 0.029 | 0.067 |
| MM15.02.2B5.bulk | MM15.PLAS.ISO.00027.002 | 27 | 876 | 0.577<sup>5</sup> | 18.246<sup>5</sup> |
| MM15.02.2A2.bulk | MM15.PLAS.ISO.00027.001 | 27 | 904 | 0.523 | 20.371 |
| MM15.02.2A4.bulk | MM15.PLAS.ISO.00027.003 | 27 | 489 | 0.582 | 25.291 |
| MM15.03.2B6.bulk | MM15.PLAS.ISO.00037.002 | 37 | 729 | 0.412 | 18.431 |
| MM15.03.2A3.bulk | MM15.PLAS.ISO.00037.001 | 37 | 571 | 0.439 | 14.984 |
| MM15.03.1B2.bulk | MM15.PLAS.ISO.00037.003 | 37 | 387 | 0.407 | 19.592 |
| MM15.03.2B1.bulk | MM15.PLAS.ISO.00037.004 | 37 | 389 | 0.273 | 21.927 |
| MM15.06.2A1.bulk | MM15.PLAS.ISO.00079.001 | 79 | 603 | 0.326 | 14.380 |
| MM15.07.2A3.bulk | MM15.PLAS.ISO.00107.001 | 107 | 1,317 | 0.489 | 15.166 |
| MM15.08.2B1.bulk | MM15.PLAS.ISO.00191.001 | 191 | 668 | 0.194 | 7.782 |
| MM15.11.2B4.bulk | MM15.PLAS.ISO.00373.001 | 373 | 645 | 0.153 | 3.676 |
| MM15.13.2C1.bulk | MM15.PLAS.ISO.00548.001 | 548 | 879 | 0.247<sup>5</sup> | 9.677<sup>5</sup> |
| MM15.14.2C3.bulk | MM15.PLAS.ISO.00720.001 | 720 | 589 | 0.224 | 9.385 |
| MM15.16.1B2.QVOA | MM15.PBMC.QVOA.01403.001 | 1,403 | 467 | 0.332 | 11.236 |
| MM23.01.2A3 | MM23.PLAS.ISO.00009.011 | 9 | 142 | 1.114 |  |
| MM23.01.2C2 | MM23.PLAS.ISO.00009.012 | 9 | 186 | 0.925 |  |
| MM23.01.11A3 | MM23.PLAS.ISO.00009.001 | 9 | 283 | 0.635 |  |
| MM23.01.11A6 | MM23.PLAS.ISO.00009.002 | 9 | 249 | 0.784<sup>5</sup> |  |
| MM23.01.11B5 | MM23.PLAS.ISO.00009.003 | 9 | 232 | 0.730 |  |
| MM23.01.11C4 | MM23.PLAS.ISO.00009.004 | 9 | 168 | 0.765<sup>5</sup> |  |
| MM23.01.13A4 | MM23.PLAS.ISO.00009.005 | 9 | 321 | 0.708<sup>5</sup> |  |
| MM23.01.13A5 | MM23.PLAS.ISO.00009.006 | 9 | 331 | 0.614 | 17.458 |
| MM23.01.13B3 | MM23.PLAS.ISO.00009.007 | 9 | 524 | 0.735 | 15.166 |
| MM23.01.13C2 | MM23.PLAS.ISO.00009.008 | 9 | 960 | 1.189 | 15.293 |
| MM23.01.13D2 | MM23.PLAS.ISO.00009.009 | 9 | 926 | 0.891 | 14.955 |
| MM23.01.14D5 | MM23.PLAS.ISO.00009.010 | 9 | 1,392 | 0.673 |  |
| MM23.05.2C1 | MM23.PLAS.ISO.00064.004 | 64 | 245 | 0.542 | 14.380 |
| MM23.05.14B3 | MM23.PLAS.ISO.00064.001 | 64 | 591 | 0.356 | 10.750 |
| MM23.05.14C3 | MM23.PLAS.ISO.00064.002 | 64 | 218 | 0.389 | 11.426 |
| MM23.05.14D2 | MM23.PLAS.ISO.00064.003 | 64 | 317 | 0.355 | 8.262 |

**Table S2**
|  |  |  |  |  |  |
| --- | --- | --- | --- | --- | --- |
| MM23.07.1C3 | MM23.PLAS.ISO.00204.001 | 204 | 499 | 0.042 |  |
| MM23.07.2A2 | MM23.PLAS.ISO.00204.002 | 204 | 119 | 0.048 |  |
| MM23.07.2A4 | MM23.PLAS.ISO.00204.003 | 204 | 151 | 0.083 | 0.181 |
| MM23.07.2A6 | MM23.PLAS.ISO.00204.004 | 204 | 292 | 0.191 | 1.766 |
| MM23.07.2B2 | MM23.PLAS.ISO.00204.005 | 204 | 177 | 0.029 | 2.090 |
| MM23.07.2B3 | MM23.PLAS.ISO.00204.006 | 204 | 100 | 0.257 |  |
| MM23.07.2B5 | MM23.PLAS.ISO.00204.007 | 204 | 526 | 0.007 | 0.186 |
| MM23.07.2C6 | MM23.PLAS.ISO.00204.008 | 204 | 397 | 0.187 |  |
| MM23.08.13C6 | MM23.PLAS.ISO.00316.001 | 316 | 168 | 0.006 | 0.013 |
| MM23.08.21B6 | MM23.PLAS.ISO.00316.004 | 316 | 362 | 0.004 |  |
| MM23.08.21C2 | MM23.PLAS.ISO.00316.005 | 316 | 144 | 0.003 | 0.006 |
| MM23.08.22A3 | MM23.PLAS.ISO.00316.006 | 316 | 434 | 0.003 | 0.006 |
| MM23.08.22B4 | MM23.PLAS.ISO.00316.007 | 316 | 477 | 0.004 |  |
| MM23.08.22B5 | MM23.PLAS.ISO.00316.008 | 316 | 431 | 0.002 | 0.002 |
| MM23.08.22C6 | MM23.PLAS.ISO.00316.009 | 316 | 427 | 0.003 |  |
| MM23.11.1A1 | MM23.PLAS.ISO.00722.001 | 722 | 291 | 0.007 |  |
| MM23.11.1A5 | MM23.PLAS.ISO.00722.002 | 722 | 358 | 0.002 | 0.006 |
| MM23.11.1C3 | MM23.PLAS.ISO.00722.003 | 722 | 361 | 0.004 |  |
| MM23.11.1D3 | MM23.PLAS.ISO.00722.004 | 722 | 157 | 0.002 | 0.007 |
| MM23.11.1D4 | MM23.PLAS.ISO.00722.005 | 722 | 165 | 0.002 | 0.004 |
| MM23.11.1D6 | MM23.PLAS.ISO.00722.006 | 722 | 128 | 0.002 | 0.008 |
| MM23.11.2B2 | MM23.PLAS.ISO.00722.007 | 722 | 403 | 0.008<sup>5</sup> |  |
| MM23.11.2B3 | MM23.PLAS.ISO.00722.008 | 722 | 268 | 0.006<sup>5</sup> |  |
| MM23.11.2C2 | MM23.PLAS.ISO.00722.009 | 722 | 290 | 0.009 |  |
| MM23.12.21B1 | MM23.PLAS.ISO.01065.001 | 1,065 | 300 | 0.023 | 0.011 |
| MM23.12.22A3 | MM23.PLAS.ISO.01065.002 | 1,065 | 284 | 0.033 | 0.023 |
| MM23.12.22C1 | MM23.PLAS.ISO.01065.003 | 1,065 | 231 | 0.026 | 0.073 |
| MM23.12.22C6 | MM23.PLAS.ISO.01065.004 | 1,065 | 323 | 0.032 | 0.041 |
| MM23.13.1B4 | MM23.PLAS.ISO.01534.001 | 1,534 | 335 | 0.068<sup>5</sup> |  |
| MM23.13.2D1 | MM23.PLAS.ISO.01534.008 | 1,534 | 157 | 0.053<sup>5</sup> | 0.246 |
| MM23.13.21A3 | MM23.PLAS.ISO.01534.002 | 1,534 | 290 | 0.045<sup>5</sup> |  |
| MM23.13.21B5 | MM23.PLAS.ISO.01534.003 | 1,534 | 374 | 0.040<sup>5</sup> | 0.130 |
| MM23.13.21B6 | MM23.PLAS.ISO.01534.004 | 1,534 | 272 | 0.052 | 0.256 |
| MM23.13.21D4 | MM23.PLAS.ISO.01534.005 | 1,534 | 284 | 0.038 |  |
| MM23.13.22B3 | MM23.PLAS.ISO.01534.006 | 1,534 | 302 | 0.050 |  |
| MM23.13.22C5 | MM23.PLAS.ISO.01534.007 | 1,534 | 427 | 0.027 | 0.064 |
| MM23.18.1A1.QVOA | MM23.PBMC.QVOA.02738.001 | 2,738 | 354 | 0.059 | 0.099 |
| MM23.18.2A1.QVOA | MM23.PBMC.QVOA.02738.002 | 2,738 | 284 | 0.044 | 0.046 |
| MM33.01.1C1 | MM33.PLAS.ISO.00012.007 | 12 | 418 | 0.734<sup>5</sup> |  |
| MM33.01.13C1 | MM33.PLAS.ISO.00012.001 | 12 | 1,013 | 0.820 | 13.488 |
| MM33.01.14C4 | MM33.PLAS.ISO.00012.002 | 12 | 558 | 0.394<sup>5</sup> | 7.507 |
| MM33.01.16A5 | MM33.PLAS.ISO.00012.003 | 12 | 582 | 0.744<sup>5</sup> | 9.406 |
| MM33.01.16C3 | MM33.PLAS.ISO.00012.004 | 12 | 1,035 | 0.739 |  |
| MM33.01.18A5 | MM33.PLAS.ISO.00012.005 | 12 | 552 | 0.581<sup>5</sup> |  |
| MM33.01.19C4 | MM33.PLAS.ISO.00012.006 | 12 | 91 | 0.419<sup>5</sup> | 6.764 |
| MM33.01.20D1 | MM33.PLAS.ISO.00012.008 | 12 | 1,020 | 0.295 |  |
| MM33.07.1A2 | MM33.PLAS.ISO.00096.001 | 96 | 471 | 0.150<sup>5</sup> | 4.223 |
| MM33.07.1B6 | MM33.PLAS.ISO.00096.002 | 96 | 137 | 0.093<sup>5</sup> | 3.217 |
| MM33.07.1D4 | MM33.PLAS.ISO.00096.003 | 96 | 367 | 0.133<sup>5</sup> | 0.612 |
| MM33.07.2D2 | MM33.PLAS.ISO.00096.004 | 96 | 757 | 0.147<sup>5</sup> | 2.419 |
| MM33.08.1A2 | MM33.PLAS.ISO.00201.001 | 201 | 390 | 0.019<sup>5</sup> | 0.047 |
| MM33.08.1A3 | MM33.PLAS.ISO.00201.002 | 201 | 372 | 0.050<sup>5</sup> | 0.047 |
| MM33.08.1A4 | MM33.PLAS.ISO.00201.003 | 201 | 320 | 0.045<sup>5</sup> |  |
| MM33.08.1D2 | MM33.PLAS.ISO.00201.004 | 201 | 312 | 0.012<sup>5</sup> | 0.012 |

**Table S2**
|  |  |  |  |  |  |
| --- | --- | --- | --- | --- | --- |
| MM33.08.1D4 | MM33.PLAS.ISO.00201.005 | 201 | 533 | 0.057<sup>5</sup> | 0.061 |
| MM33.08.1D5 | MM33.PLAS.ISO.00201.006 | 201 | 791 | 0.043<sup>5</sup> |  |
| MM33.08.2A3 | MM33.PLAS.ISO.00201.007 | 201 | 436 | 0.018<sup>5</sup> |  |
| MM33.08.2B6 | MM33.PLAS.ISO.00201.008 | 201 | 587 | 0.043<sup>5</sup> | 0.052 |
| MM33.08.1A4.bulk | MM33.PLAS.ISO.00201.009 | 201 | 313 | 0.030 | 0.031 |
| MM33.08.1A3.bulk | MM33.PLAS.ISO.00201.010 | 201 | 344 | 0.025 | 0.016 |
| MM33.13.1D3 | MM33.PLAS.ISO.00719.004 | 719 | 681 | 0.008 | 0.006 |
| MM33.13.1D6 | MM33.PLAS.ISO.00719.005 | 719 | 244 | 0.004<sup>5</sup> |  |
| MM33.13.2A4 | MM33.PLAS.ISO.00719.006 | 719 | 507 | 0.004 | 0.003 |
| MM33.13.2D6 | MM33.PLAS.ISO.00719.007 | 719 | 174 | 0.005<sup>5</sup> | 0.006 |
| MM33.13.11B1 | MM33.PLAS.ISO.00719.001 | 719 | 608 | 0.003 | 0.002 |
| MM33.13.11B5 | MM33.PLAS.ISO.00719.002 | 719 | 684 | 0.009 |  |
| MM33.13.12A4 | MM33.PLAS.ISO.00719.003 | 719 | 598 | 0.003 |  |
| MM33.13.1.bulk | MM33.PLAS.ISO.00719.008 | 719 | 121 | 0.003 | 0.006 |
| MM33.14.11A3 | MM33.PLAS.ISO.01006.001 | 1,006 | 682 | 0.009<sup>5</sup> | 0.004 |
| MM33.14.11B1 | MM33.PLAS.ISO.01006.002 | 1,006 | 682 | 0.005 |  |
| MM33.14.11B4 | MM33.PLAS.ISO.01006.003 | 1,006 | 645 | 0.004 |  |
| MM33.14.11C6 | MM33.PLAS.ISO.01006.004 | 1,006 | 690 | 0.004 |  |
| MM33.14.12A2 | MM33.PLAS.ISO.01006.005 | 1,006 | 580 | 0.004 | 0.008 |
| MM33.14.12A4 | MM33.PLAS.ISO.01006.006 | 1,006 | 559 | 0.004<sup>5</sup> | 0.009 |
| MM33.14.12C6 | MM33.PLAS.ISO.01006.007 | 1,006 | 639 | 0.005<sup>5</sup> | 0.008 |
| MM33.14.1A1.bulk | MM33.PLAS.ISO.01006.008 | 1,006 | 106 | 0.004 | 0.014 |
| MM33.14.1A2.bulk | MM33.PLAS.ISO.01006.009 | 1,006 | 536 | 0.003 | 0.012 |
| MM33.17.1D2 | MM33.PLAS.ISO.01721.009 | 1,721 | 493 | 0.019 | 0.005 |
| MM33.17.1D6 | MM33.PLAS.ISO.01721.001 | 1,721 | 260 | 0.014 | 0.017 |
| MM33.17.2A4 | MM33.PLAS.ISO.01721.002 | 1,721 | 639 | 0.017<sup>5</sup> | 0.015 |
| MM33.17.2B3 | MM33.PLAS.ISO.01721.003 | 1,721 | 140 | 0.019<sup>5</sup> |  |
| MM33.17.2B4 | MM33.PLAS.ISO.01721.004 | 1,721 | 59 | 0.028<sup>5</sup> | 0.007 |
| MM33.17.2B5 | MM33.PLAS.ISO.01721.005 | 1,721 | 114 | 0.017<sup>5</sup> |  |
| MM33.17.2C3 | MM33.PLAS.ISO.01721.006 | 1,721 | 50 | 0.009 |  |
| MM33.17.2D4 | MM33.PLAS.ISO.01721.007 | 1,721 | 356 | 0.029<sup>5</sup> | 0.003 |
| MM33.17.2D5 | MM33.PLAS.ISO.01721.008 | 1,721 | 484 | 0.008 | 0.006 |
| MM33.17.1A1.bulk | MM33.PLAS.ISO.01721.010 | 1,721 | 381 | 0.031 | 0.022 |
| MM33.17.1A2.bulk | MM33.PLAS.ISO.01721.011 | 1,721 | 615 | 0.040 | 0.019 |
| MM33.19.1A2 | MM33.PLAS.ISO.02031.001 | 2,031 | 565 | 0.015 |  |
| MM33.19.1A4 | MM33.PLAS.ISO.02031.002 | 2,031 | 331 | 0.018 |  |
| MM33.19.1B1 | MM33.PLAS.ISO.02031.003 | 2,031 | 616 | 0.017 |  |
| MM33.19.1C2 | MM33.PLAS.ISO.02031.004 | 2,031 | 274 | 0.011 | 0.089 |
| MM33.19.2A4 | MM33.PLAS.ISO.02031.005 | 2,031 | 244 | 0.027 | 0.047 |
| MM33.19.2B1 | MM33.PLAS.ISO.02031.006 | 2,031 | 224 | 0.016 | 0.028 |
| MM33.19.2B5 | MM33.PLAS.ISO.02031.007 | 2,031 | 689 | 0.010 | 0.133 |
| MM33.19.2C1 | MM33.PLAS.ISO.02031.008 | 2,031 | 621 | 0.018 |  |
| MM33.19.2D2 | MM33.PLAS.ISO.02031.009 | 2,031 | 591 | 0.032 |  |
| MM33.19.3A1 | MM33.PLAS.ISO.02031.010 | 2,031 | 220 | 0.036 |  |
| MM33.19.3B4 | MM33.PLAS.ISO.02031.011 | 2,031 | 242 | 0.035 |  |
| MM33.19.3D2 | MM33.PLAS.ISO.02031.012 | 2,031 | 551 | 0.024 |  |
| MM33.19.4A5 | MM33.PLAS.ISO.02031.013 | 2,031 | 149 | 0.022 |  |
| MM33.19.4B5 | MM33.PLAS.ISO.02031.014 | 2,031 | 655 | 0.033 |  |
| MM33.19.4D1 | MM33.PLAS.ISO.02031.015 | 2,031 | 628 | 0.046 |  |
| MM33.19.1B1.bulk | MM33.PLAS.ISO.02031.016 | 2,031 | 267 | 0.029 | 0.057 |
| MM34.01.1A5 | MM34.PLAS.ISO.00017.002 | 17 | 176 | 0.518 | 9.675 |
| MM34.01.1B3 | MM34.PLAS.ISO.00017.003 | 17 | 154 | 0.425 | 17.497 |
| MM34.01.1B6 | MM34.PLAS.ISO.00017.004 | 17 | 279 | 0.485 |  |
| MM34.01.1C2 | MM34.PLAS.ISO.00017.001 | 17 | 135 | 1.485 | 17.631 |

**Table S2**
|  |  |  |  |  |  |
| --- | --- | --- | --- | --- | --- |
| MM34.01.1D1 | MM34.PLAS.ISO.00017.006 | 17 | 128 | 1.094 |  |
| MM34.01.2A4 | MM34.PLAS.ISO.00017.007 | 17 | 237 | 0.725 |  |
| MM34.01.2B3 | MM34.PLAS.ISO.00017.008 | 17 | 343 | 0.386 |  |
| MM34.01.2C2 | MM34.PLAS.ISO.00017.009 | 17 | 200 | 0.944 | 18.025 |
| MM34.01.2D4 | MM34.PLAS.ISO.00017.010 | 17 | 218 | 0.691 |  |
| MM34.01.2D5 | MM34.PLAS.ISO.00017.011 | 17 | 503 | 0.821 |  |
| MM34.06.21C6 | MM34.PLAS.ISO.00074.001 | 74 | 266 | 0.478 | 10.531 |
| MM34.08.11B5 | MM34.PLAS.ISO.00192.001 | 192 | 392 | 0.032<sup>5</sup> | 0.168<sup>5</sup> |
| MM34.08.11C2 | MM34.PLAS.ISO.00192.002 | 192 | 439 | 0.037<sup>5</sup> | 3.350<sup>5</sup> |
| MM34.08.11D3 | MM34.PLAS.ISO.00192.004 | 192 | 409 | 0.04 | 3.475<sup>5</sup> |
| MM34.08.21A3 | MM34.PLAS.ISO.00192.005 | 192 | 624 | 0.025 | 2.088<sup>5</sup> |
| MM34.12.21A1 | MM34.PLAS.ISO.00760.001 | 760 | 340 | 0.026 | 0.035 |
| MM34.12.21C5 | MM34.PLAS.ISO.00760.002 | 760 | 236 | 0.012 | 0.021 |
| MM34.12.21D1 | MM34.PLAS.ISO.00760.003 | 760 | 59 | 0.030 | 0.049 |
| MM34.12.21D3 | MM34.PLAS.ISO.00760.004 | 760 | 100 | 0.028 | 0.030 |
| MM34.13.11A1 | MM34.PLAS.ISO.01007.001 | 1,007 | 66 | 0.005 | 0.004 |
| MM34.13.11A2 | MM34.PLAS.ISO.01007.002 | 1,007 | 268 | 0.006 |  |
| MM34.13.11B1 | MM34.PLAS.ISO.01007.003 | 1,007 | 154 | 0.004 | 0.001 |
| MM34.13.11C5 | MM34.PLAS.ISO.01007.004 | 1,007 | 640 | 0.017 | 0.002 |
| MM34.13.11D2 | MM34.PLAS.ISO.01007.005 | 1,007 | 143 | 0.002 | 0.001 |
| MM34.13.11D5 | MM34.PLAS.ISO.01007.006 | 1,007 | 214 | 0.007 |  |
| MM34.14.1A4 | MM34.PLAS.ISO.01313.001 | 1,313 | 482 | 0.072 |  |
| MM34.14.1B2 | MM34.PLAS.ISO.01313.002 | 1,313 | 389 | 0.018 |  |
| MM34.14.1B3 | MM34.PLAS.ISO.01313.003 | 1,313 | 354 | 0.062 | 0.060 |
| MM34.14.1C6 | MM34.PLAS.ISO.01313.004 | 1,313 | 646 | 0.036 |  |
| MM34.14.1D4 | MM34.PLAS.ISO.01313.005 | 1,313 | 478 | 0.022 | 0.124 |
| MM34.14.2A1 | MM34.PLAS.ISO.01313.006 | 1,313 | 596 | 0.041<sup>5</sup> | 0.326<sup>5</sup> |
| MM34.14.2B1 | MM34.PLAS.ISO.01313.007 | 1,313 | 153 |  | 0.050 |
| MM34.15.11A1 | MM34.PLAS.ISO.01399.001 | 1,399 | 188 | 0.007 | 0.004 |
| MM34.15.11C1 | MM34.PLAS.ISO.01399.002 | 1,399 | 242 | 0.008 | 0.001 |
| MM34.15.11D3 | MM34.PLAS.ISO.01399.003 | 1,399 | 384 | 0.004<sup>5</sup> | 0.001<sup>5</sup> |
| MM34.16.11A5 | MM34.PLAS.ISO.01691.002 | 1,691 | 414 | 0.017 | 0.052 |
| MM34.16.11C1 | MM34.PLAS.ISO.01691.003 | 1,691 | 278 | 0.048 | 0.035 |
| MM34.16.11C3 | MM34.PLAS.ISO.01691.004 | 1,691 | 591 | 0.032 |  |
| MM34.16.11D1 | MM34.PLAS.ISO.01691.005 | 1,691 | 413 | 0.026 | 0.045 |
| MM34.16.11D2 | MM34.PLAS.ISO.01691.006 | 1,691 | 255 | 0.056 | 0.019 |
| MM34.18.1A4 | MM34.PLAS.ISO.02229.003 | 2,229 | 619 | 0.072 |  |
| MM34.18.1A6 | MM34.PLAS.ISO.02229.004 | 2,229 | 588 | 0.088 | 0.139 |
| MM34.18.1C5 | MM34.PLAS.ISO.02229.006 | 2,229 | 232 | 0.098 |  |
| MM34.18.2A4 | MM34.PLAS.ISO.02229.009 | 2,229 | 274 | 0.058 | 0.466 |
| MM34.18.2A6 | MM34.PLAS.ISO.02229.010 | 2,229 | 102 | 0.106 | 0.490 |
| MM34.18.2C2 | MM34.PLAS.ISO.02229.011 | 2,229 | 258 | 0.052 |  |
| MM34.18.2D1 | MM34.PLAS.ISO.02229.013 | 2,229 | 118 | 0.073 |  |
| MM34.18.2D2 | MM34.PLAS.ISO.02229.014 | 2,229 | 276 | 0.062 | 0.429 |
| MM34.23.1B2.QVOA | MM34.PBMC.QVOA.02911.001 | 2,911 | 392 | 0.059 | 0.150 |
| MM34.25.3A5.QVOA | MM34.PBMC.QVOA.03261.002 | 3,261 | 303 | 0.033 | 0.204 |
| MM34.25.2B5.QVOA | MM34.PBMC.QVOA.03261.001 | 3,261 | 254 | 0.026 | 0.162 |
| MM39.02.1B4 | MM39.PLAS.ISO.00011.001 | 11 | 424 | 0.584<sup>5</sup> | 14.857 |
| MM39.02.2A2 | MM39.PLAS.ISO.00011.002 | 11 | 180 | 0.756<sup>5</sup> |  |
| MM39.02.2B5 | MM39.PLAS.ISO.00011.003 | 11 | 276 | 0.700<sup>5</sup> |  |
| MM39.02.13B5 | MM39.PLAS.ISO.00011.004 | 11 | 437 | 0.799<sup>5</sup> |  |
| MM39.02.13C3 | MM39.PLAS.ISO.00011.005 | 11 | 326 | 0.796<sup>5</sup> |  |
| MM39.02.13D2 | MM39.PLAS.ISO.00011.006 | 11 | 275 | 0.631<sup>5</sup> | 9.659 |
| MM39.02.13D3 | MM39.PLAS.ISO.00011.007 | 11 | 249 | 0.620 |  |

**Table S2**
|  |  |  |  |  |  |
| --- | --- | --- | --- | --- | --- |
| MM39.02.13D6 | MM39.PLAS.ISO.00011.008 | 11 | 453 | 0.731<sup>5</sup> |  |
| MM39.02.14C1 | MM39.PLAS.ISO.00011.010 | 11 | 637 | 0.897<sup>5</sup> |  |
| MM39.02.14C2 | MM39.PLAS.ISO.00011.011 | 11 | 566 | 0.787<sup>5</sup> |  |
| MM39.02.14C3 | MM39.PLAS.ISO.00011.012 | 11 | 828 | 0.767<sup>5</sup> | 17.371 |
| MM39.02.14C5 | MM39.PLAS.ISO.00011.013 | 11 | 758 | 0.652<sup>5</sup> | 15.713 |
| MM39.02.14D4 | MM39.PLAS.ISO.00011.014 | 11 | 132 | 0.929<sup>5</sup> |  |
| MM39.02.14D5 | MM39.PLAS.ISO.00011.015 | 11 | 228 | 0.850<sup>5</sup> |  |
| MM39.07.11A3 | MM39.PLAS.ISO.00179.001 | 179 | 288 | 0.104<sup>5</sup> | 5.980 |
| MM39.07.11A4 | MM39.PLAS.ISO.00179.002 | 179 | 178 | 0.042 | 4.497 |
| MM39.07.11C1 | MM39.PLAS.ISO.00179.003 | 179 | 117 | 0.185<sup>5</sup> | 3.633 |
| MM39.07.11C2 | MM39.PLAS.ISO.00179.004 | 179 | 195 | 0.140<sup>5</sup> | 6.435 |
| MM39.10.11B3 | MM39.PLAS.ISO.00354.001 | 354 | 94 | 0.007<sup>5</sup> |  |
| MM39.10.11B4 | MM39.PLAS.ISO.00354.002 | 354 | 298 | 0.007<sup>5</sup> |  |
| MM39.10.11D1 | MM39.PLAS.ISO.00354.003 | 354 | 415 | 0.006<sup>5</sup> |  |
| MM39.10.11D4 | MM39.PLAS.ISO.00354.004 | 354 | 198 | 0.004<sup>5</sup> | 0.148 |
| MM39.10.11D6 | MM39.PLAS.ISO.00354.005 | 354 | 129 | 0.007 |  |
| MM39.10.12A2 | MM39.PLAS.ISO.00354.006 | 354 | 127 | 0.007 |  |
| MM39.10.12A6 | MM39.PLAS.ISO.00354.007 | 354 | 143 | 0.003 |  |
| MM39.10.12B5 | MM39.PLAS.ISO.00354.008 | 354 | 91 | 0.007 |  |
| MM39.10.12C3 | MM39.PLAS.ISO.00354.009 | 354 | 121 | 0.004 |  |
| MM39.10.12C4 | MM39.PLAS.ISO.00354.010 | 354 | 130 | 0.005<sup>5</sup> | 0.137<sup>5</sup> |
| MM39.10.12D3 | MM39.PLAS.ISO.00354.011 | 354 | 271 | 0.008 | 0.114 |
| MM39.10.12D4 | MM39.PLAS.ISO.00354.012 | 354 | 80 | 0.005 |  |
| MM39.10.14A3 | MM39.PLAS.ISO.00354.015 | 354 | 295 | 0.003 |  |
| MM39.10.14B4 | MM39.PLAS.ISO.00354.013 | 354 | 86 | 0.007 | 0.079 |
| MM39.10.14C5 | MM39.PLAS.ISO.00354.014 | 354 | 206 | 0.005 |  |
| MM39.10.1C1.bulk | MM39.PLAS.ISO.00354.016 | 354 | 121 | 0.004<sup>5</sup> | 0.214<sup>5</sup> |
| MM39.13.1B1 | MM39.PLAS.ISO.00639.001 | 639 | 594 | 0.008 | 0.030 |
| MM39.13.11A2 | MM39.PLAS.ISO.00639.002 | 639 | 180 | 0.006 |  |
| MM39.13.11A4 | MM39.PLAS.ISO.00639.003 | 639 | 349 | 0.007 |  |
| MM39.13.11B1 | MM39.PLAS.ISO.00639.004 | 639 | 188 | 0.005 |  |
| MM39.13.11B2 | MM39.PLAS.ISO.00639.008 | 639 | 124 | 0.007 | 0.007 |
| MM39.13.11C3 | MM39.PLAS.ISO.00639.005 | 639 | 379 | 0.006 | 0.031 |
| MM39.13.11C5 | MM39.PLAS.ISO.00639.006 | 639 | 167 | 0.010 | 0.023 |
| MM39.13.11D1 | MM39.PLAS.ISO.00639.007 | 639 | 247 | 0.005 |  |
| MM39.13.1C3.bulk | MM39.PLAS.ISO.00639.009 | 639 | 66 | 0.003 | 0.022 |
| MM39.14.1A2 | MM39.PLAS.ISO.00821.001 | 821 | 107 | 0.042 | 0.005 |
| MM39.14.1A5 | MM39.PLAS.ISO.00821.002 | 821 | 339 | 0.040 | 0.004 |
| MM39.14.1B3 | MM39.PLAS.ISO.00821.003 | 821 | 547 | 0.039 |  |
| MM39.14.2A3 | MM39.PLAS.ISO.00821.004 | 821 | 233 | 0.026 |  |
| MM39.14.2B1 | MM39.PLAS.ISO.00821.005 | 821 | 455 | 0.035<sup>5</sup> | 0.006<sup>5</sup> |
| MM39.14.2B5 | MM39.PLAS.ISO.00821.007 | 821 | 318 | 0.023 | 0.020 |
| MM39.14.2D4 | MM39.PLAS.ISO.00821.006 | 821 | 93 | 0.022 |  |
| MM39.16.1A4 | MM39.PLAS.ISO.01206.002 | 1,206 | 189 | 0.009 |  |
| MM39.16.1B2 | MM39.PLAS.ISO.01206.003 | 1,206 | 239 | 0.024<sup>5</sup> | 0.200 |
| MM39.16.1B4 | MM39.PLAS.ISO.01206.004 | 1,206 | 167 | 0.017<sup>5</sup> | 0.121 |
| MM39.16.1C4 | MM39.PLAS.ISO.01206.005 | 1,206 | 107 | 0.016<sup>5</sup> | 0.123 |
| MM39.16.2A5 | MM39.PLAS.ISO.01206.006 | 1,206 | 172 | 0.022<sup>5</sup> | 0.192 |
| MM39.16.2C3 | MM39.PLAS.ISO.01206.001 | 1,206 | 539 | 0.020<sup>5</sup> |  |
| MM39.16.2D2 | MM39.PLAS.ISO.01206.007 | 1,206 | 635 | 0.018<sup>5</sup> |  |
| MM39.16.13A2 | MM39.PLAS.ISO.01206.013 | 1,206 | 426 | 0.068 |  |
| MM39.16.13A5 | MM39.PLAS.ISO.01206.008 | 1,206 | 113 | 0.037 | 0.291 |
| MM39.16.13B1 | MM39.PLAS.ISO.01206.009 | 1,206 | 117 | 0.059 | 0.216 |
| MM39.16.13B3 | MM39.PLAS.ISO.01206.010 | 1,206 | 270 | 0.020 |  |
| MM39.16.14B3 | MM39.PLAS.ISO.01206.011 | 1,206 | 183 | 0.023 |  |

**Table S2**
|  |  |  |  |  |  |
| --- | --- | --- | --- | --- | --- |
| MM39.16.14C1 | MM39.PLAS.ISO.01206.012 | 1,206 | 617 | 0.010 | 0.123 |
| MM39.16.1A1.bulk | MM39.PLAS.ISO.01206.014 | 1,206 | 80 | 0.010 | 0.205 |
| MM39.16.1A2.bulk | MM39.PLAS.ISO.01206.015 | 1,206 | 493 | 0.041 | 0.669 |
| MM39.16.1A3.bulk | MM39.PLAS.ISO.01206.016 | 1,206 | 628 | 0.046 | 0.349 |
| MM40.01.1A1 | MM40.PLAS.ISO.00011.001 | 11 | 518 | 0.712<sup>5</sup> | 5.287 |
| MM40.01.1A2 | MM40.PLAS.ISO.00011.002 | 11 | 736 | 0.741<sup>5</sup> |  |
| MM40.01.1A4 | MM40.PLAS.ISO.00011.003 | 11 | 557 | 0.736<sup>5</sup> |  |
| MM40.01.1A5 | MM40.PLAS.ISO.00011.004 | 11 | 709 | 0.722 |  |
| MM40.01.1A6 | MM40.PLAS.ISO.00011.005 | 11 | 654 | 0.782 |  |
| MM40.01.1B1 | MM40.PLAS.ISO.00011.006 | 11 | 315 | 0.723<sup>5</sup> | 5.738 |
| MM40.01.1B2 | MM40.PLAS.ISO.00011.007 | 11 | 727 | 0.999 |  |
| MM40.01.1B3 | MM40.PLAS.ISO.00011.008 | 11 | 532 | 0.653 |  |
| MM40.01.1B4 | MM40.PLAS.ISO.00011.009 | 11 | 132 | 0.233 |  |
| MM40.01.1B5 | MM40.PLAS.ISO.00011.010 | 11 | 685 | 0.627 |  |
| MM40.01.1C6 | MM40.PLAS.ISO.00011.011 | 11 | 538 | 0.528 |  |
| MM40.01.1D1 | MM40.PLAS.ISO.00011.012 | 11 | 484 | 0.652 |  |
| MM40.01.2A4 | MM40.PLAS.ISO.00011.013 | 11 | 629 | 0.821 |  |
| MM40.01.2B5 | MM40.PLAS.ISO.00011.014 | 11 | 312 | 0.679 |  |
| MM40.01.2C2 | MM40.PLAS.ISO.00011.015 | 11 | 366 | 0.681 | 7.361 |
| MM40.01.2D4 | MM40.PLAS.ISO.00011.016 | 11 | 657 | 0.722 | 7.624 |
| MM40.01.2D5 | MM40.PLAS.ISO.00011.017 | 11 | 417 | 0.782 |  |
| MM40.01.2A1.bulk | MM40.PLAS.ISO.00011.018 | 11 | 1,254 | 0.732 | 11.993 |
| MM40.01.2A2.bulk | MM40.PLAS.ISO.00011.019 | 11 | 583 | 0.394 | 13.197 |
| MM40.07.11A3 | MM40.PLAS.ISO.00095.004 | 95 | 421 | 0.429 | 1.899 |
| MM40.07.11A4 | MM40.PLAS.ISO.00095.001 | 95 | 319 | 0.590 |  |
| MM40.07.11B1 | MM40.PLAS.ISO.00095.002 | 95 | 314 | 0.750 |  |
| MM40.07.11B3 | MM40.PLAS.ISO.00095.005 | 95 | 387 | 0.701 |  |
| MM40.07.11B4 | MM40.PLAS.ISO.00095.003 | 95 | 287 | 0.149 |  |
| MM40.07.11B5 | MM40.PLAS.ISO.00095.006 | 95 | 266 | 0.208 | 1.598 |
| MM40.07.11C1 | MM40.PLAS.ISO.00095.008 | 95 | 200 | 0.120 | 2.256 |
| MM40.07.12B1 | MM40.PLAS.ISO.00095.007 | 95 | 233 | 0.154 | 2.568 |
| MM40.12.13A6 | MM40.PLAS.ISO.00747.001 | 747 | 166 | 0.006 | 0.008 |
| MM40.12.13C1 | MM40.PLAS.ISO.00747.002 | 747 | 176 | 0.002 | 0.012 |
| MM40.12.13D1 | MM40.PLAS.ISO.00747.003 | 747 | 178 | 0.004 |  |
| MM40.12.14C5 | MM40.PLAS.ISO.00747.004 | 747 | 192 | 0.006<sup>5</sup> | 0.006 |
| MM40.12.14D2 | MM40.PLAS.ISO.00747.005 | 747 | 219 | 0.006<sup>5</sup> | 0.007 |
| MM40.12.1A1.bulk | MM40.PLAS.ISO.00747.006 | 747 | 267 | 0.010 | 0.011 |
| MM40.12.1A2.bulk | MM40.PLAS.ISO.00747.007 | 747 | 529 | 0.003 | 0.011 |
| MM40.13.11A1 | MM40.PLAS.ISO.01166.002 | 1,166 | 289 | 0.009<sup>5</sup> |  |
| MM40.13.11A5 | MM40.PLAS.ISO.01166.003 | 1,166 | 237 | 0.017<sup>5</sup> |  |
| MM40.13.11B3 | MM40.PLAS.ISO.01166.004 | 1,166 | 208 | 0.008<sup>5</sup> |  |
| MM40.13.11D4 | MM40.PLAS.ISO.01166.005 | 1,166 | 238 | 0.011<sup>5</sup> | 0.017 |
| MM40.13.12A1 | MM40.PLAS.ISO.01166.006 | 1,166 | 182 | 0.007<sup>5</sup> | 0.025 |
| MM40.13.12B2 | MM40.PLAS.ISO.01166.007 | 1,166 | 189 | 0.006<sup>5</sup> | 0.009 |
| MM40.13.12B5 | MM40.PLAS.ISO.01166.008 | 1,166 | 195 | 0.009<sup>5</sup> |  |
| MM40.13.12C2 | MM40.PLAS.ISO.01166.009 | 1,166 | 263 | 0.010<sup>5</sup> | 0.010 |
| MM40.13.12C3 | MM40.PLAS.ISO.01166.010 | 1,166 | 251 | 0.006<sup>5</sup> |  |
| MM40.13.13D5 | MM40.PLAS.ISO.01166.001 | 1,166 | 162 | 0.028<sup>5</sup> | 0.008 |
| MM40.15.1A3 | MM40.PLAS.ISO.01530.001 | 1,530 | 192 | 0.048<sup>5</sup> |  |
| MM40.15.1A5 | MM40.PLAS.ISO.01530.002 | 1,530 | 241 | 0.032<sup>5</sup> |  |
| MM40.15.1B1 | MM40.PLAS.ISO.01530.003 | 1,530 | 230 | 0.025<sup>5</sup> | 0.034 |
| MM40.15.1B6 | MM40.PLAS.ISO.01530.004 | 1,530 | 251 | 0.042 |  |
| MM40.15.1C1 | MM40.PLAS.ISO.01530.005 | 1,530 | 423 | 0.067<sup>5</sup> |  |
| MM40.15.2A2 | MM40.PLAS.ISO.01530.006 | 1,530 | 126 | 0.043<sup>5</sup> |  |

**Table S2**
|  |  |  |  |  |  |
| --- | --- | --- | --- | --- | --- |
| MM40.15.2B1 | MM40.PLAS.ISO.01530.007 | 1,530 | 257 | 0.051<sup>5</sup> |  |
| MM40.15.2B5 | MM40.PLAS.ISO.01530.008 | 1,530 | 247 | 0.025<sup>5</sup> |  |
| MM40.15.2B6 | MM40.PLAS.ISO.01530.009 | 1,530 | 606 | 0.033<sup>5</sup> |  |
| MM40.15.2C1 | MM40.PLAS.ISO.01530.010 | 1,530 | 235 | 0.009<sup>5</sup> | 0.031 |
| MM40.15.2C3 | MM40.PLAS.ISO.01530.011 | 1,530 | 465 | 0.069<sup>5</sup> | 0.084 |
| MM40.15.2D1 | MM40.PLAS.ISO.01530.012 | 1,530 | 290 | 0.042<sup>5</sup> |  |
| MM40.15.2D6 | MM40.PLAS.ISO.01530.013 | 1,530 | 267 | 0.048<sup>5</sup> | 0.035 |
| MM40.15.13B6 | MM40.PLAS.ISO.01530.014 | 1,530 | 217 | 0.041 |  |
| MM40.15.13C1 | MM40.PLAS.ISO.01530.018 | 1,530 | 210 | 0.043 |  |
| MM40.15.14B3 | MM40.PLAS.ISO.01530.015 | 1,530 | 212 | 0.061 | 0.159 |
| MM40.15.14C3 | MM40.PLAS.ISO.01530.016 | 1,530 | 273 | 0.034 |  |
| MM40.15.14D5 | MM40.PLAS.ISO.01530.017 | 1,530 | 175 | 0.029 |  |
| MM40.15.1.bulk | MM40.PLAS.ISO.01530.020 | 1,530 | 144 | 0.090 | 0.271 |
| MM40.18.1C7.QVOA | MM40.PBMC.QVOA.02041.001 | 2,041 | 373 | 0.030 | 0.066 |
| MM55.01.1A3.bulk | MM55.PLAS.ISO.00031.001 | 31 | 210 | 0.019<sup>5</sup> | 0.108<sup>5</sup> |
| MM55.03.2B4.bulk | MM55.PLAS.ISO.00051.002 | 51 | 265 | 0.021<sup>5</sup> | 0.073<sup>5</sup> |
| MM55.03.2A1.bulk | MM55.PLAS.ISO.00051.001 | 51 | 310 | 0.081<sup>5</sup> | 0.422<sup>5</sup> |
| MM55.09.2A1.bulk | MM55.PLAS.ISO.00348.001 | 348 | 184 | 0.008 | 0.074 |
| MM55.11.2A3.bulk | MM55.PLAS.ISO.00661.001 | 661 | 133 | 0.002 | 0.007 |
| MM55.12.2B1.bulk | MM55.PLAS.ISO.00831.001 | 831 | 174 | 0.005<sup>5</sup> | 0.029<sup>5</sup> |
| MM55.12.2B2.bulk | MM55.PLAS.ISO.00831.002 | 831 | 181 | 0.007 | 0.011 |
| MM55.14.1B4.bulk | MM55.PLAS.ISO.01300.003 | 1,300 | 475 | 0.003 | 0.010 |
| MM55.14.2B5.bulk | MM55.PLAS.ISO.01300.001 | 1,300 | 477 | 0.003 | 0.010 |
| MM55.15.1A3.bulk | MM55.PLAS.ISO.01858.001 | 1,858 | 222 | 0.007 | 0.009 |
| MM55.15.1A4.bulk | MM55.PLAS.ISO.01858.002 | 1,858 | 209 | 0.005 | 0.047 |
| MM55.16.1C2.bulk | MM55.PLAS.ISO.02054.001 | 2,054 | 402 | 0.006 | 0.006 |
| MM55.16.1E1.bulk | MM55.PLAS.ISO.02054.002 | 2,054 | 513 | 0.007 | 0.004 |
| MM62.01.1A3.bulk | MM62.PLAS.ISO.00022.003 | 22 | 404 | 0.200<sup>5</sup> | 1.723<sup>5</sup> |
| MM62.01.1C4.bulk | MM62.PLAS.ISO.00022.001 | 22 | 227 | 0.160 | 1.614 |
| MM62.01.2C6.bulk | MM62.PLAS.ISO.00022.002 | 22 | 238 | 0.260 | 1.808 |
| MM62.02.1B1.bulk | MM62.PLAS.ISO.00057.002 | 57 | 295 | 0.090 | 0.751 |
| MM62.02.2D1.bulk | MM62.PLAS.ISO.00057.001 | 57 | 223 | 0.081 | 0.871 |
| MM62.03.1B3.bulk | MM62.PLAS.ISO.00074.001 | 74 | 255 | 0.008 | 0.014 |
| MM62.03.2D2.bulk | MM62.PLAS.ISO.00074.002 | 74 | 245 | 0.010 | 0.022 |
| MM62.04.1C1.bulk | MM62.PLAS.ISO.00120.001 | 120 | 298 | 0.005<sup>5</sup> | 0.004<sup>5</sup> |
| MM62.04.1D3.bulk | MM62.PLAS.ISO.00120.002 | 120 | 243 | 0.007 | 0.008 |
| MM62.04.1E5.bulk | MM62.PLAS.ISO.00120.003 | 120 | 279 | 0.007 | 0.008 |
| MM62.05.2C1.bulk | MM62.PLAS.ISO.00185.001 | 185 | 358 | 0.016<sup>5</sup> | 0.021<sup>5</sup> |
| MM62.05.2D5.bulk | MM62.PLAS.ISO.00185.002 | 185 | 374 | 0.030 | 0.079 |
| MM62.06.1C3.bulk | MM62.PLAS.ISO.00304.001 | 304 | 267 | 0.006 | 0.007 |
| MM62.08.2A1.bulk | MM62.PLAS.ISO.00658.001 | 658 | 362 | 0.009 | 0.011 |
| MM62.10.1D6.bulk | MM62.PLAS.ISO.01130.001 | 1,130 | 258 | 0.012 | 0.022 |
| MM62.10.1G6.bulk | MM62.PLAS.ISO.01130.002 | 1,130 | 246 | 0.014 | 0.018 |
| MM62.11.2B5.bulk | MM62.PLAS.ISO.01331.001 | 1,331 | 298 | 0.010 | 0.016 |
| MM62.11.2A2.bulk | MM62.PLAS.ISO.01331.002 | 1,331 | 307 | 0.015 | 0.022 |
| WEAU.02.1A1.bulk | WEAU.PLAS.ISO.-0004.001 | -4 | 257 | 0.843 | 19.873 |
| WEAU.02.1A2.bulk | WEAU.PLAS.ISO.-0004.002 | -4 | 464 | 0.854 | 20.646 |
| WEAU.03.1A4.bulk | WEAU.PLAS.ISO.00000.001 | 0 | 260 | 1.006 | 18.580 |
| WEAU.03.1A5.bulk | WEAU.PLAS.ISO.00000.002 | 0 | 146 | 0.972 | 17.459 |
| WEAU.08.1B1.bulk | WEAU.PLAS.ISO.00024.001 | 24 | 283 | 0.302 | 9.336 |
| WEAU.08.1B2.bulk | WEAU.PLAS.ISO.00024.002 | 24 | 295 | 0.298 | 8.530 |
| WEAU.11.1B5.bulk | WEAU.PLAS.ISO.00052.001 | 52 | 228 | 0.264 | 6.061 |

**Table S2**
|  |  |  |  |  |  |
| --- | --- | --- | --- | --- | --- |
| WEAU.11.1B6.bulk | WEAU.PLAS.ISO.00052.002 | 52 | 287 | 0.215 | 4.924 |
| WEAU.14.1C1.bulk | WEAU.PLAS.ISO.00371.001 | 371 | 306 | 0.151<sup>5</sup> | 7.139<sup>5</sup> |
| WEAU.14.1C3.bulk | WEAU.PLAS.ISO.00371.002 | 371 | 158 | 0.302 | 4.073 |
| WEAU.15.1C4.bulk | WEAU.PLAS.ISO.00520.001 | 520 | 140 | 0.266 | 8.100 |
| WEAU.15.1C5.bulk | WEAU.PLAS.ISO.00520.002 | 520 | 121 | 0.333 | 8.758 |
| WEAU.17.1D1.bulk | WEAU.PLAS.ISO.00752.001 | 752 | 146 | 0.298 | 11.100 |
| WEAU.17.1D2.bulk | WEAU.PLAS.ISO.00752.002 | 752 | 189 | 0.353 | 7.279 |
| WEAU.18.1D4.bulk | WEAU.PLAS.ISO.00771.001 | 771 | 245 | 0.241 | 6.080 |
| WEAU.18.1D5.bulk | WEAU.PLAS.ISO.00771.002 | 771 | 178 | 0.431 | 8.211 |
| WEAU.19.1A1.bulk | WEAU.PLAS.ISO.00981.001 | 981 | 299 | 0.466 | 11.906 |
| WEAU.19.1A3.bulk | WEAU.PLAS.ISO.00981.002 | 981 | 380 | 0.533 | 11.227 |

<sup>1</sup>For GenBank accession numbers see Table S5.
<sup>2</sup>Days following onset of symptoms
<sup>3</sup>Concentration of p24 in culture supernatant at day 7 in the absence of IFN treatment.
<sup>4</sup>Concentration of IFNα2 and IFNβ that reduced virus replication by 50%.
<sup>5</sup>Average of multiple biological replicates.

**Table S3.**
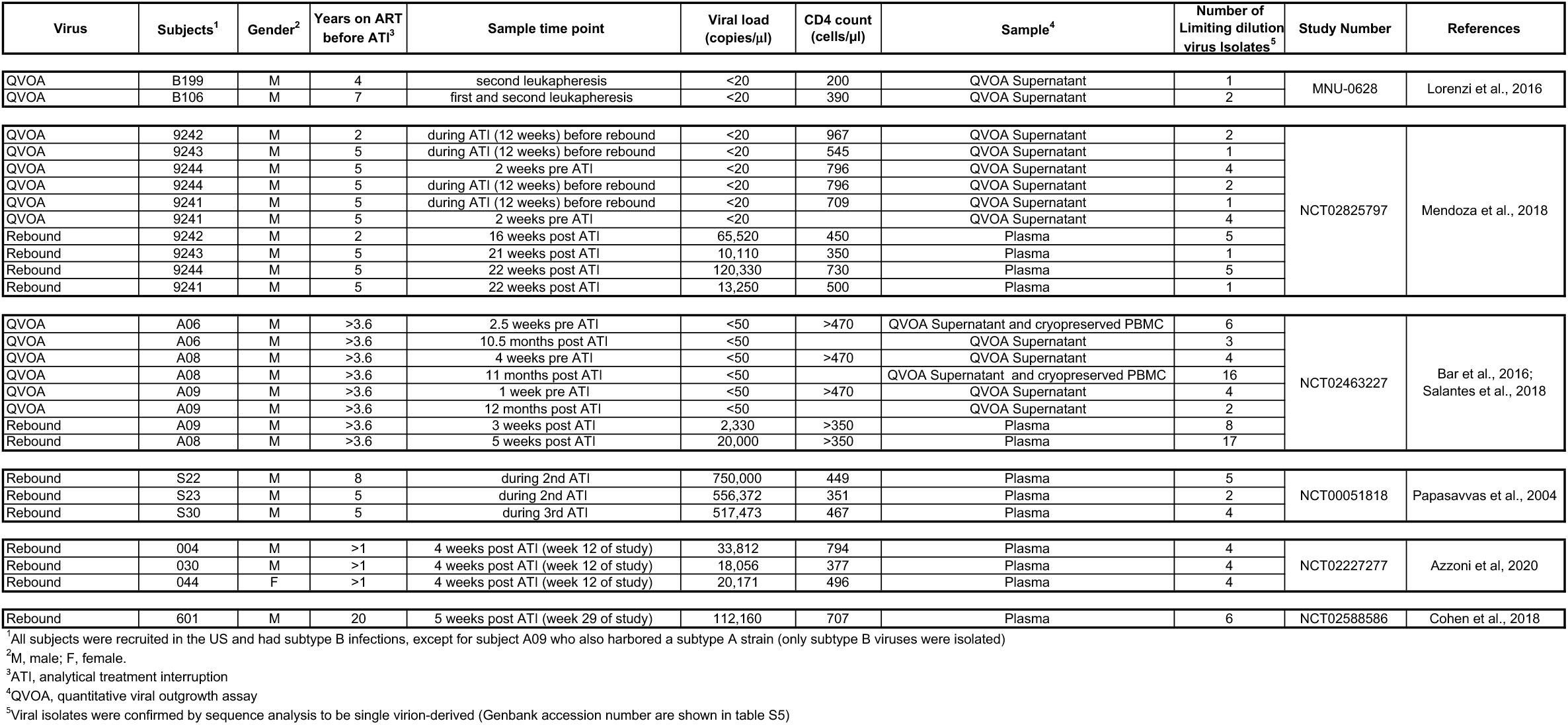
Generation of viral isolates from ART suppressed individuals with and without treatment interruption.

**Table S4.**
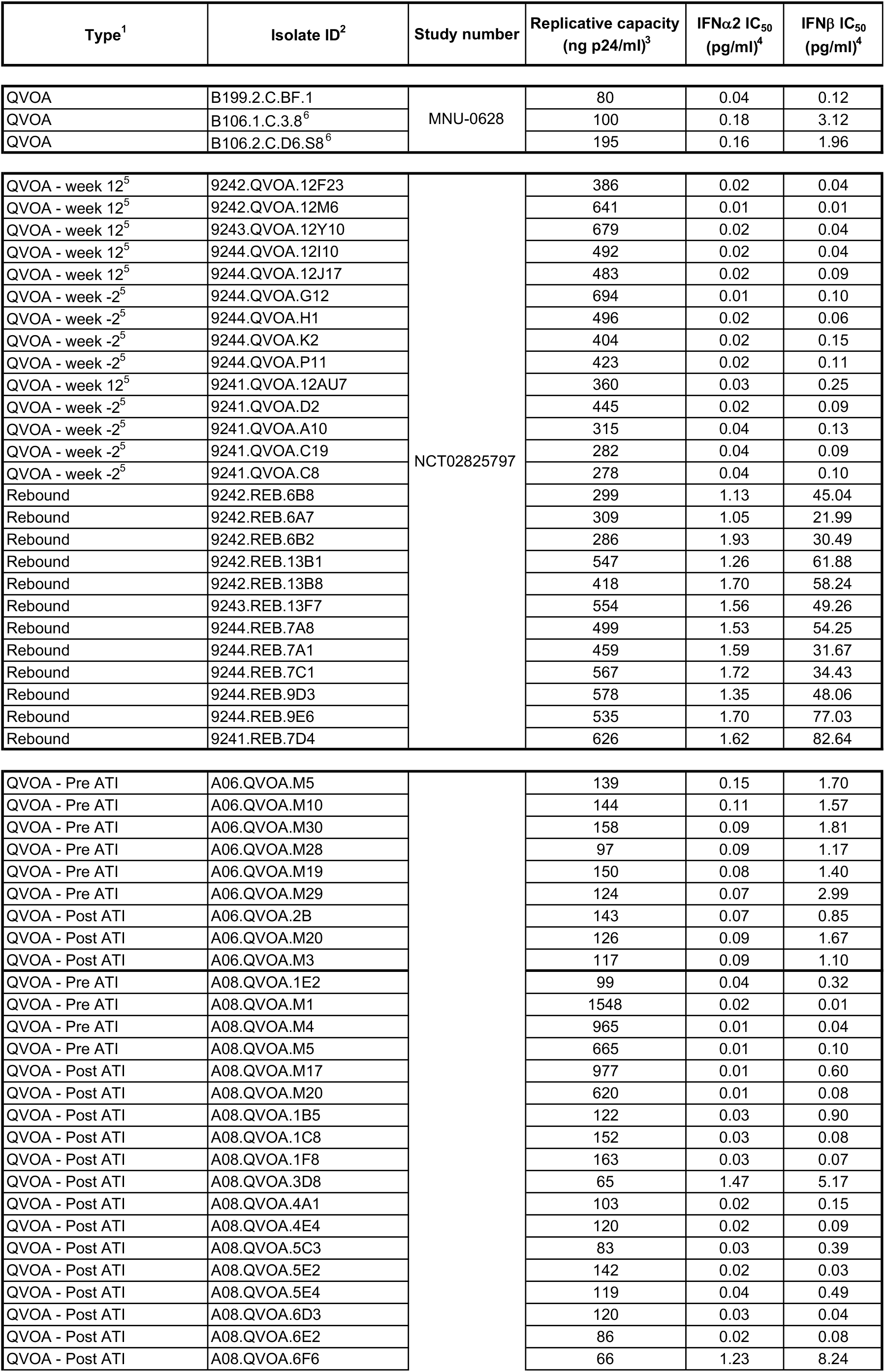
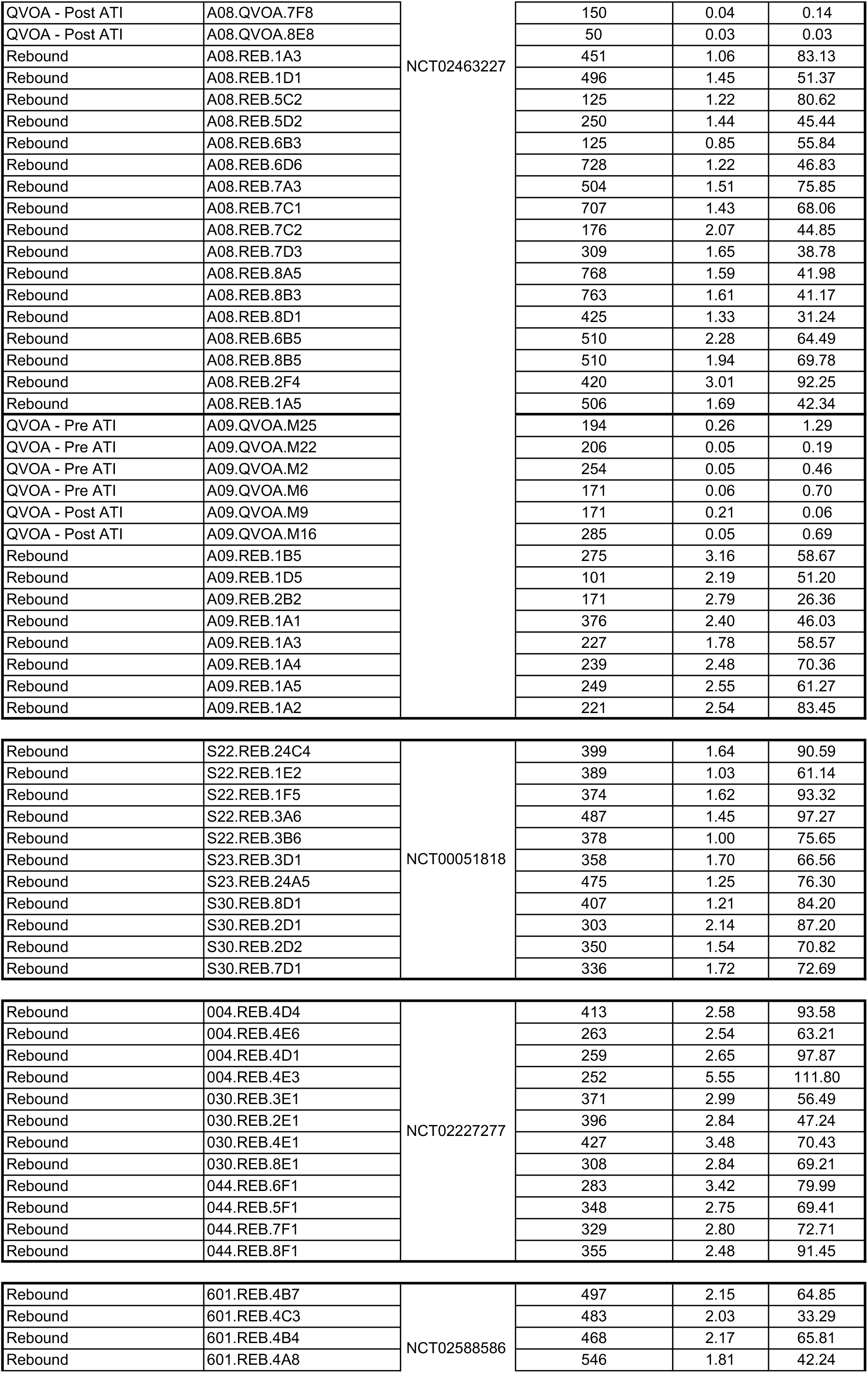
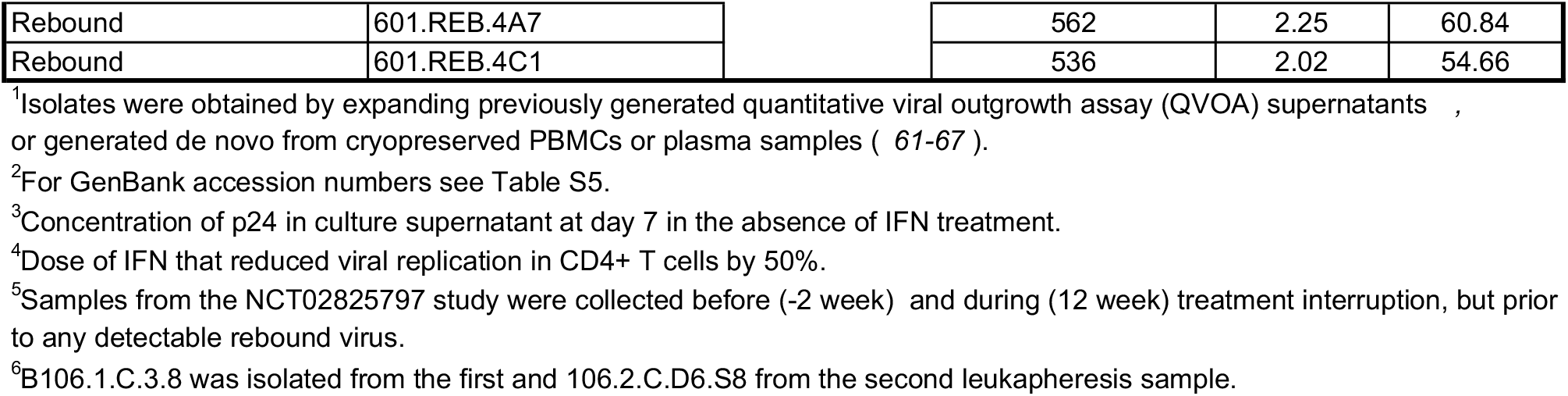
IFN-I resistance of viral outgrowth and rebound isolates.

**Table S5.**
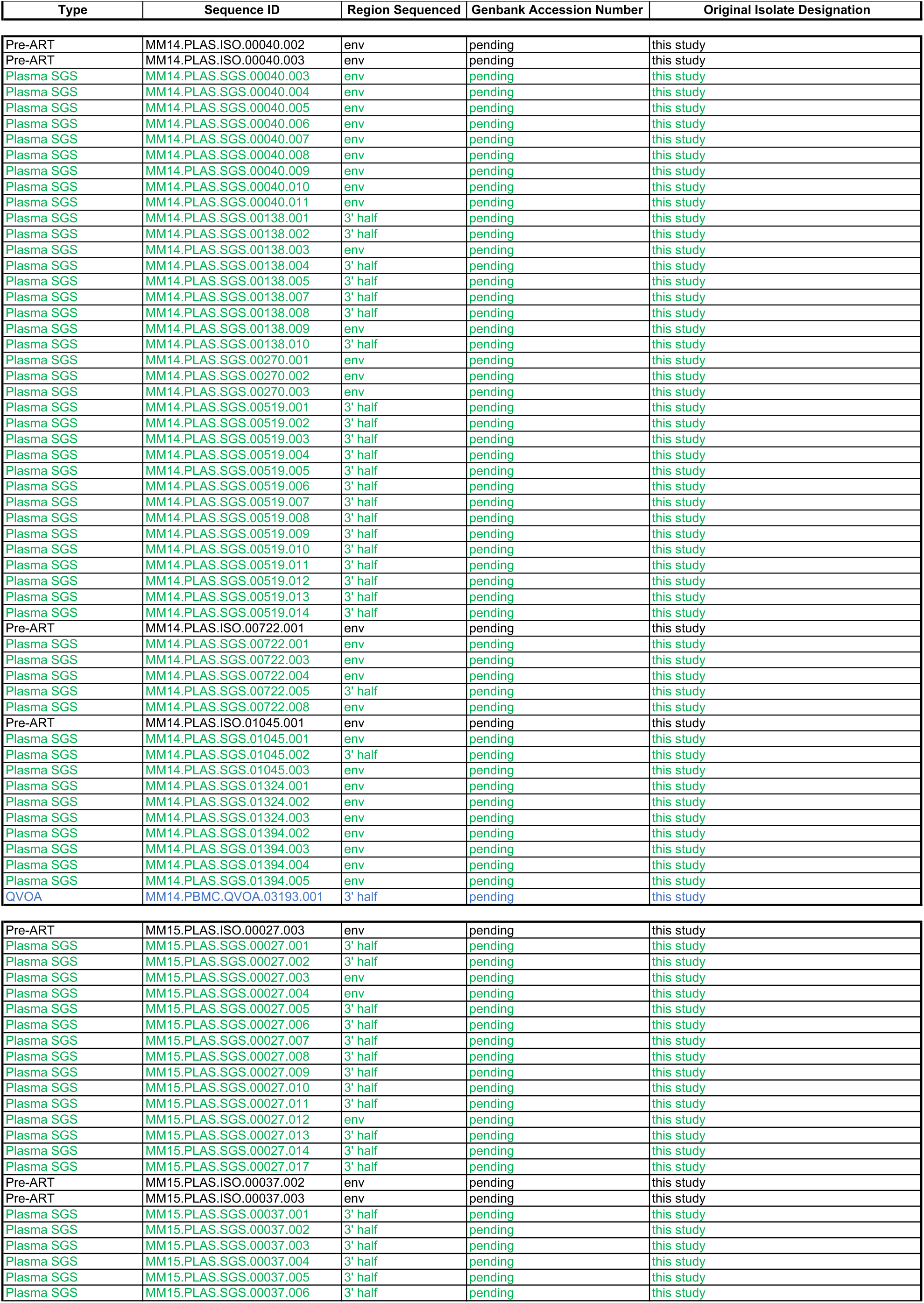
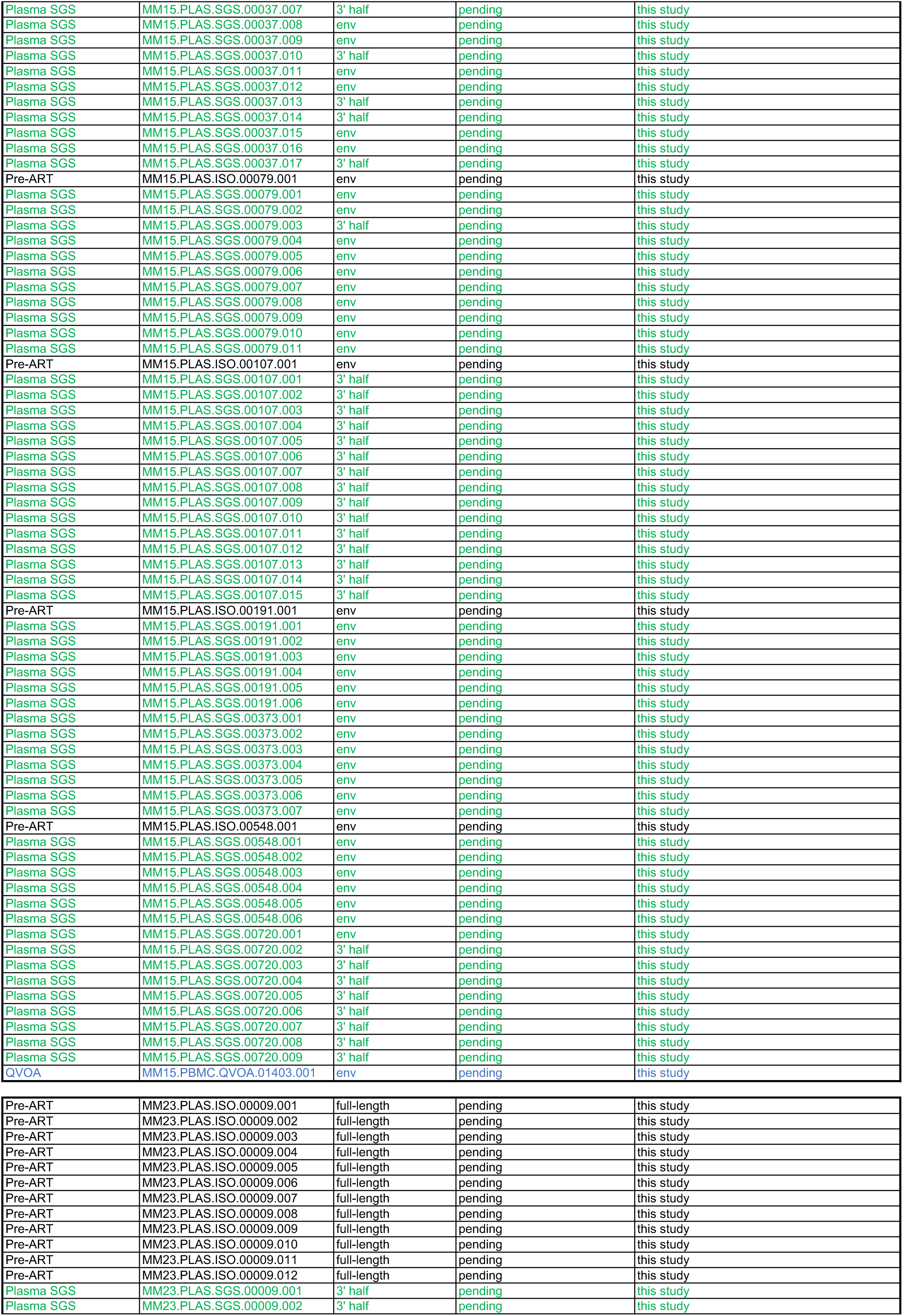
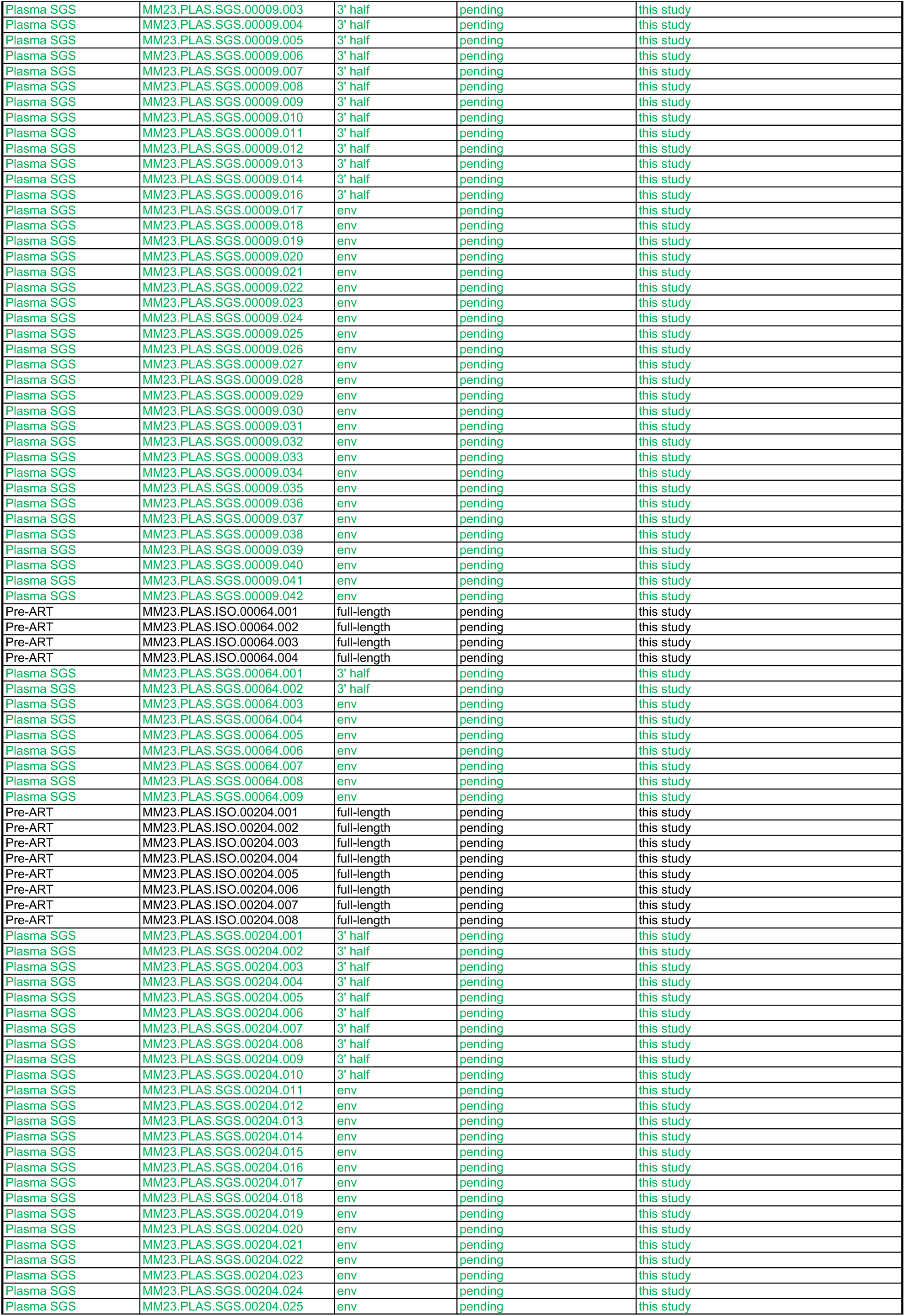
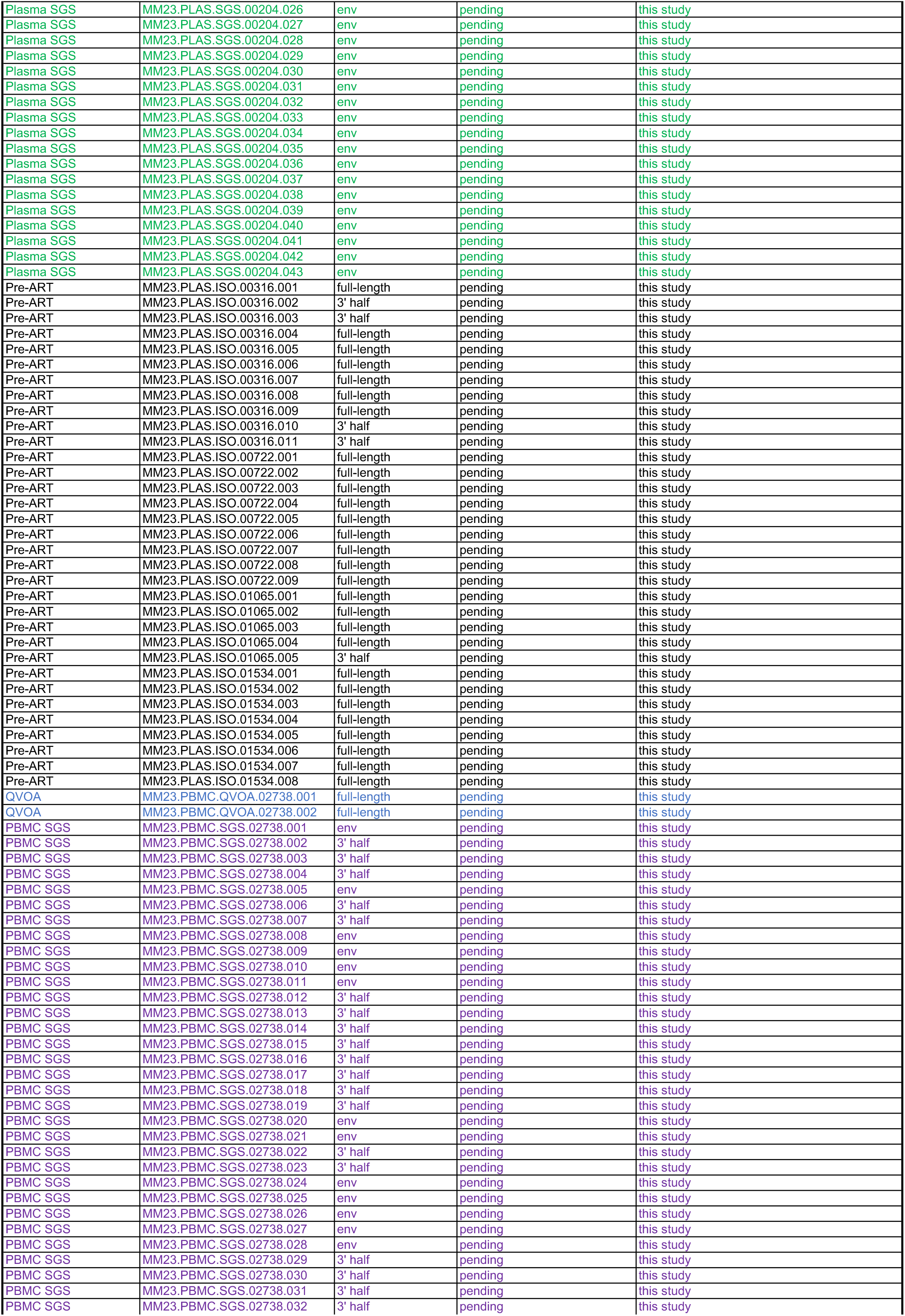
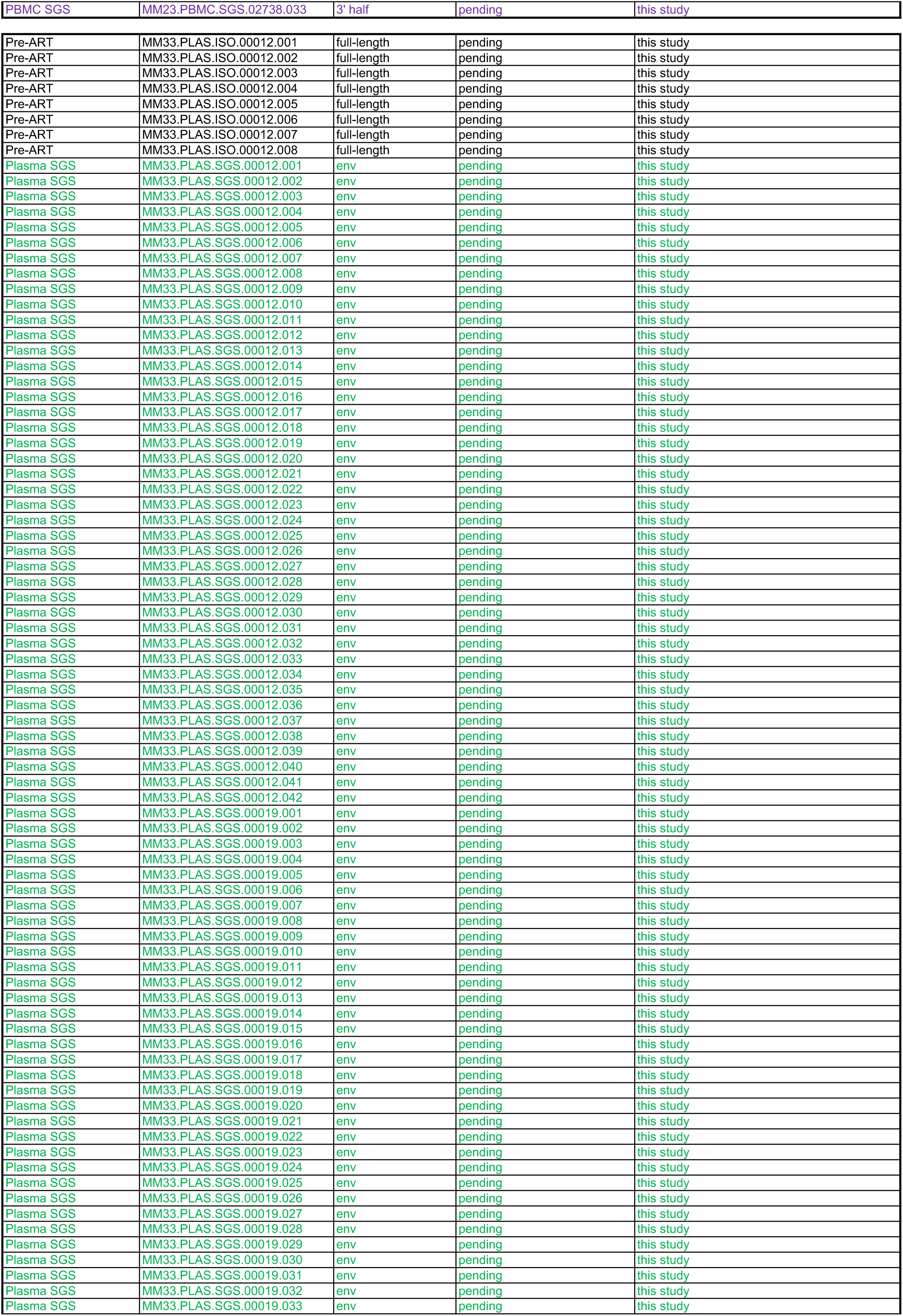
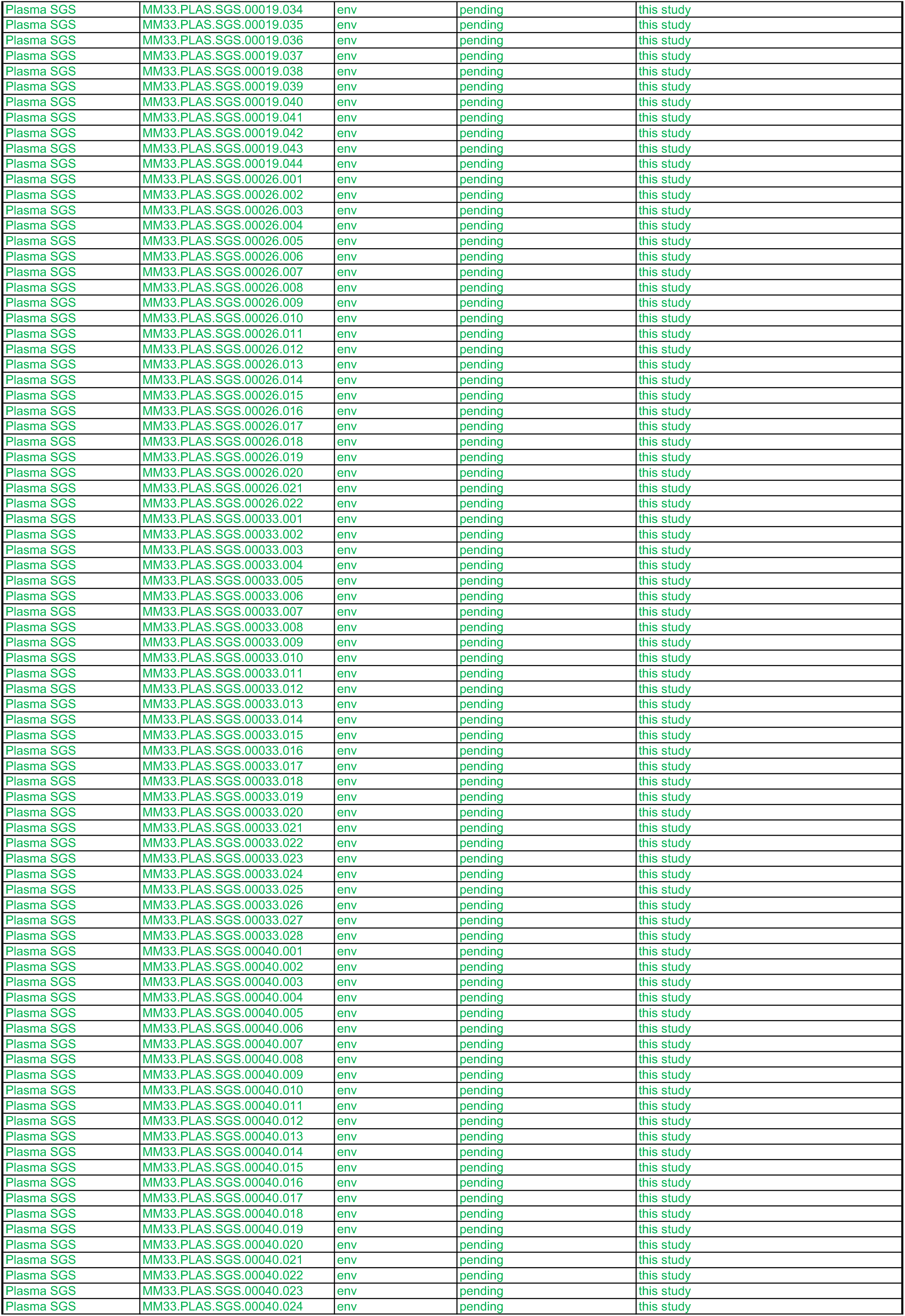
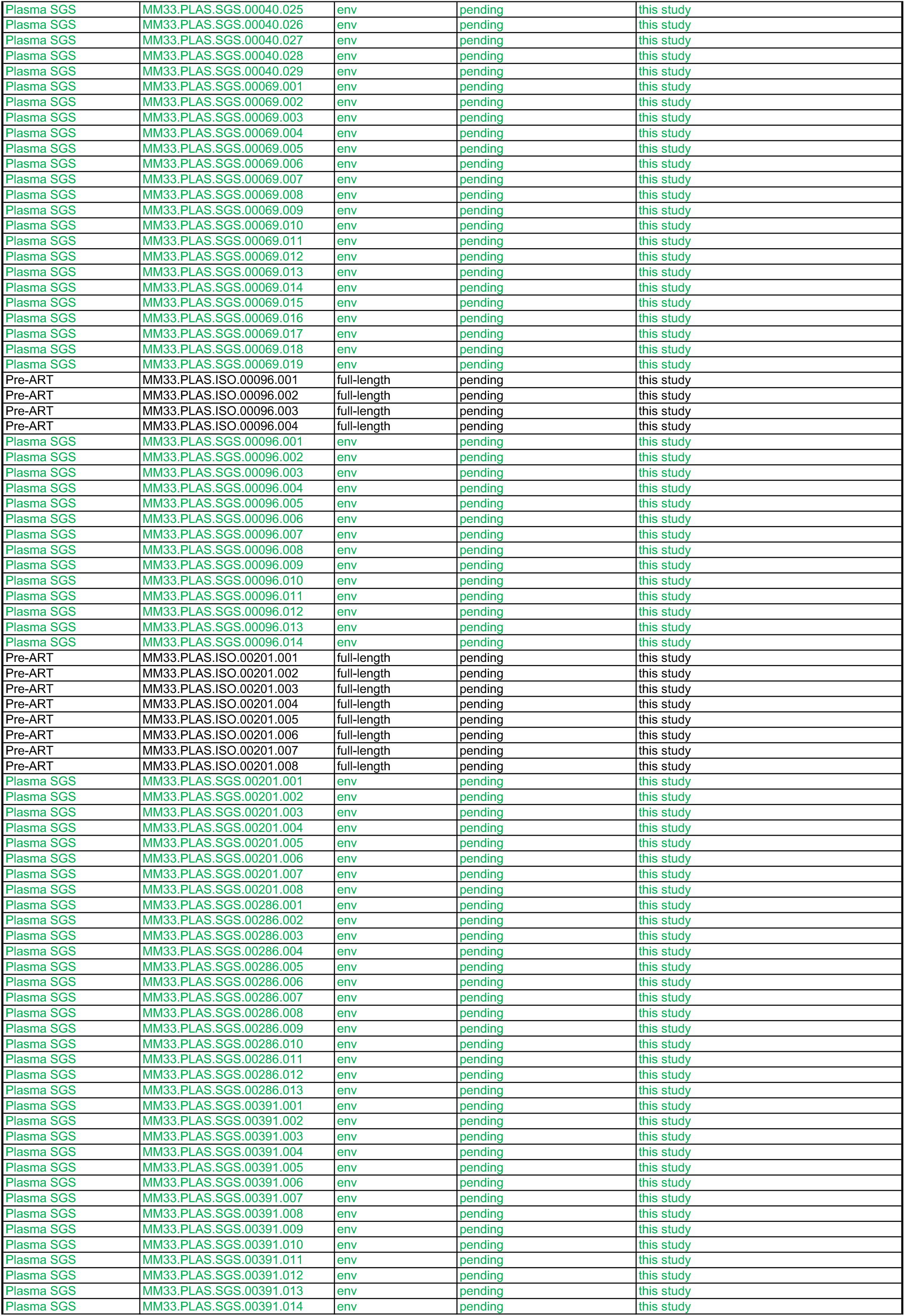
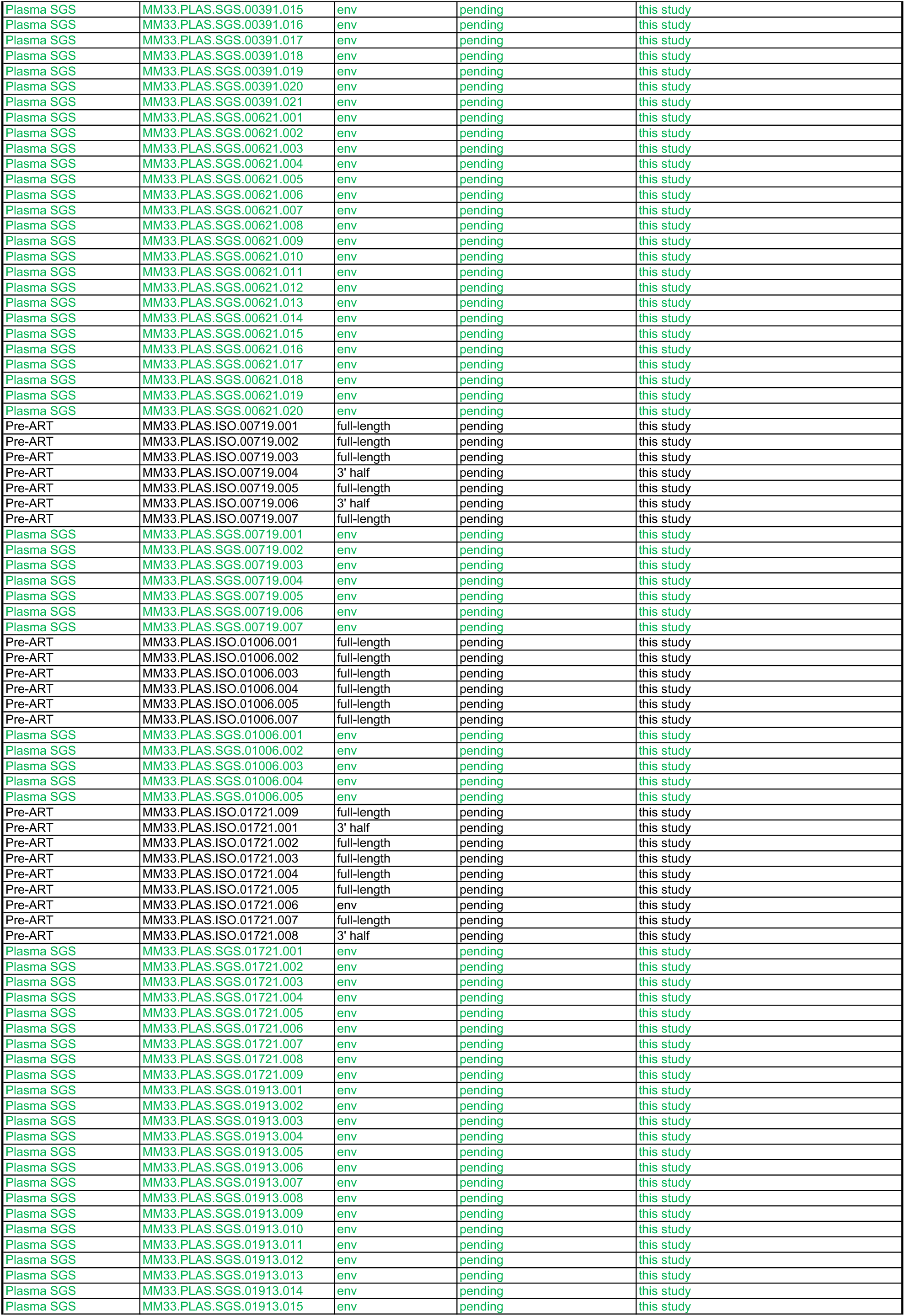
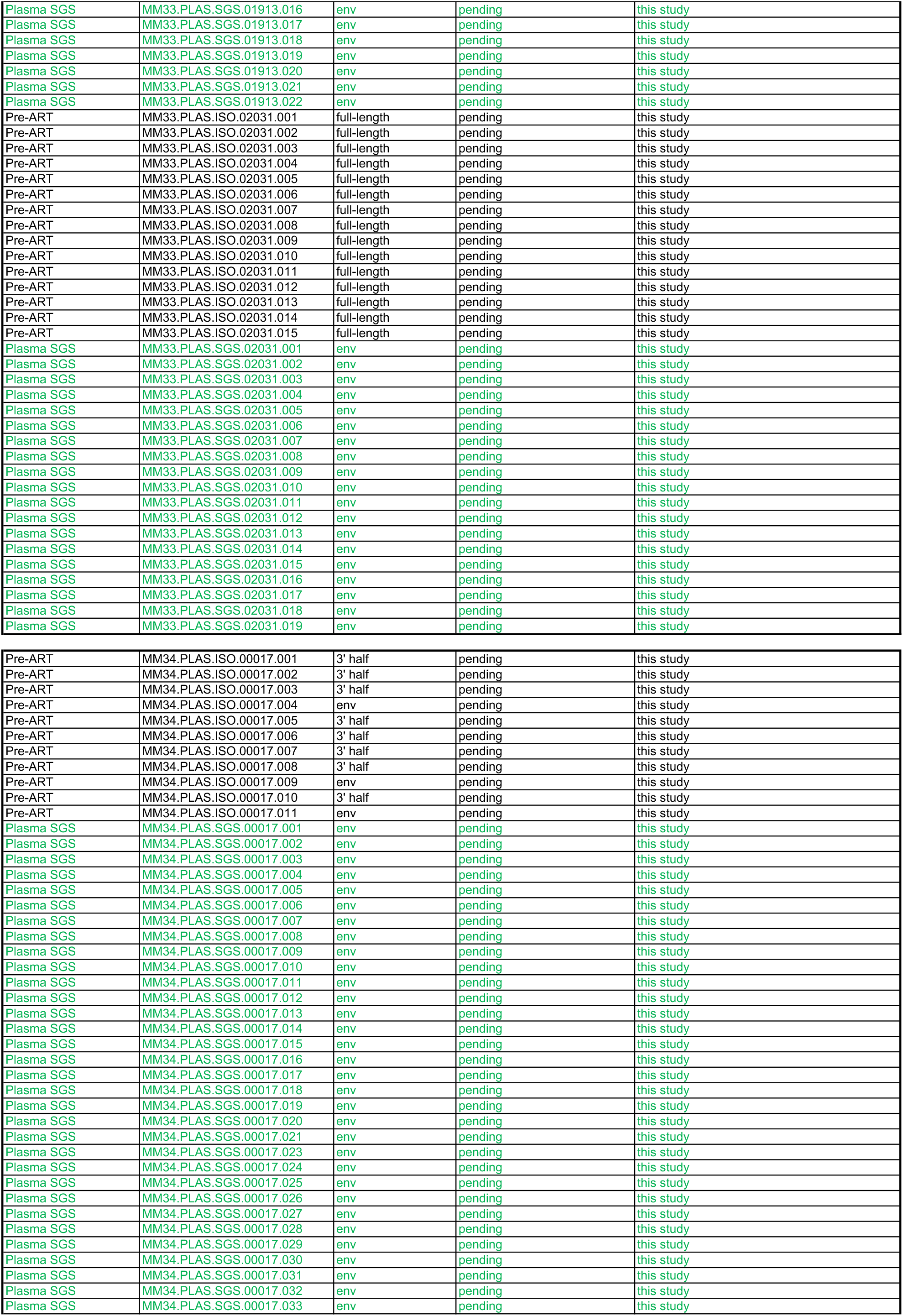
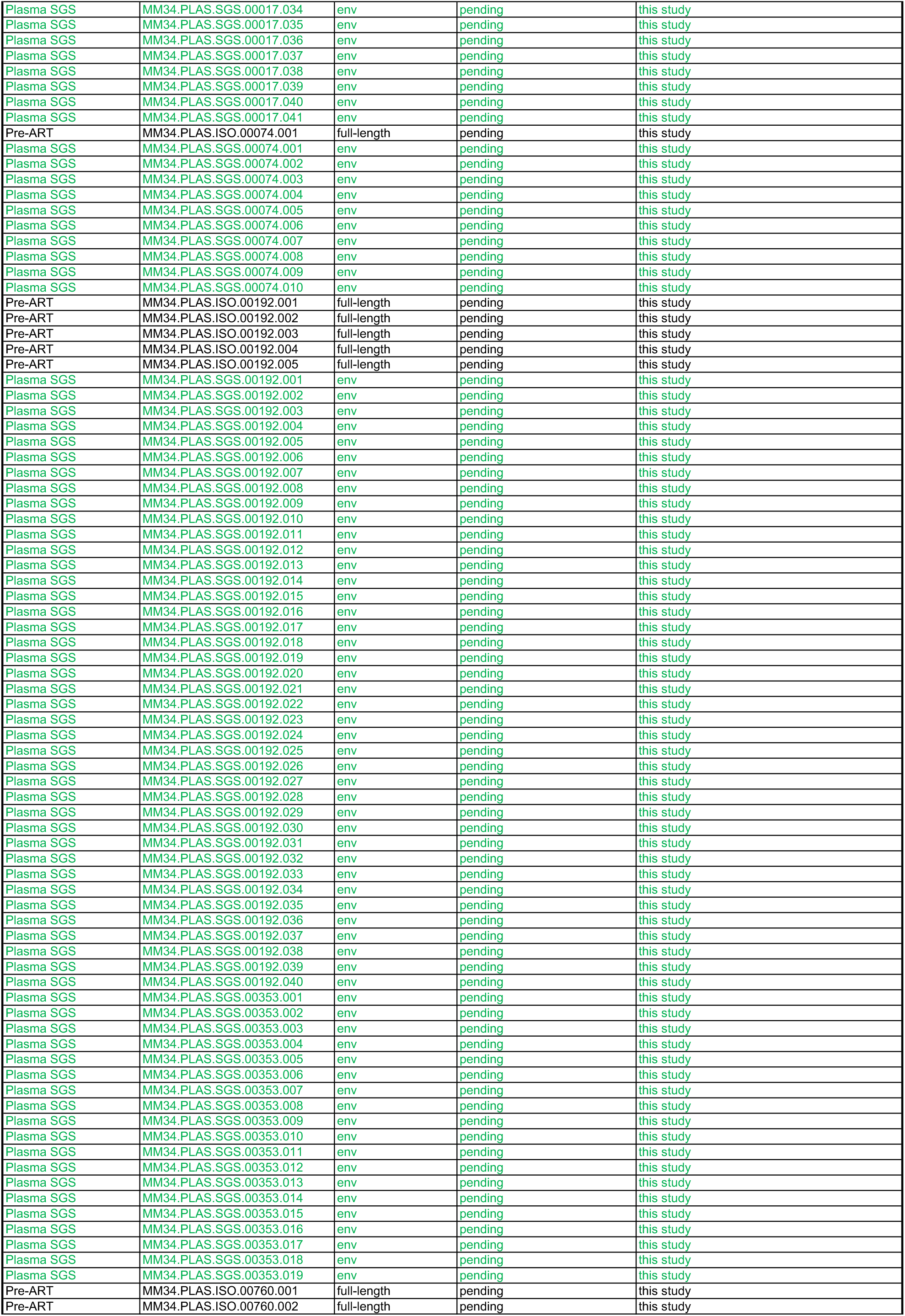
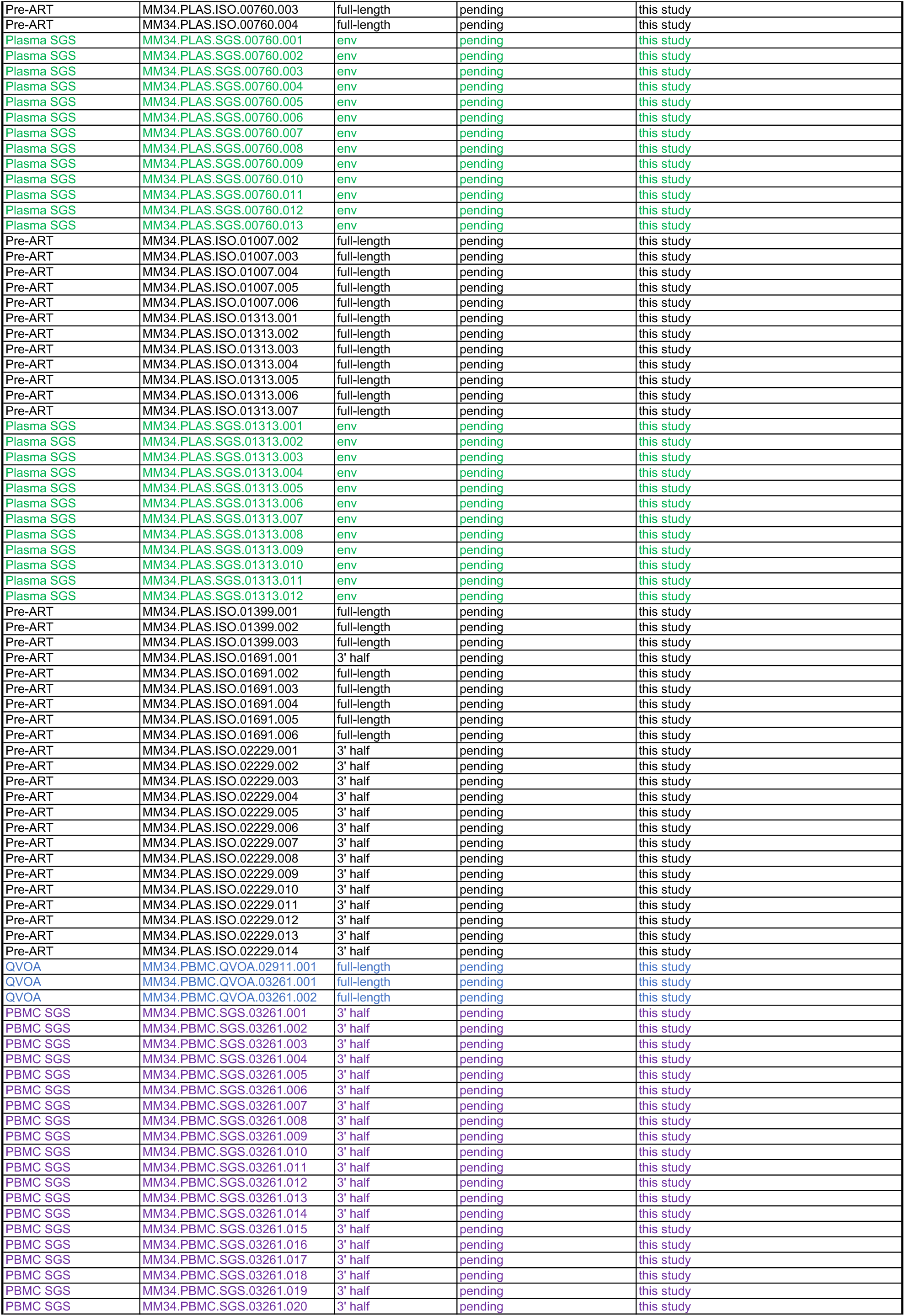
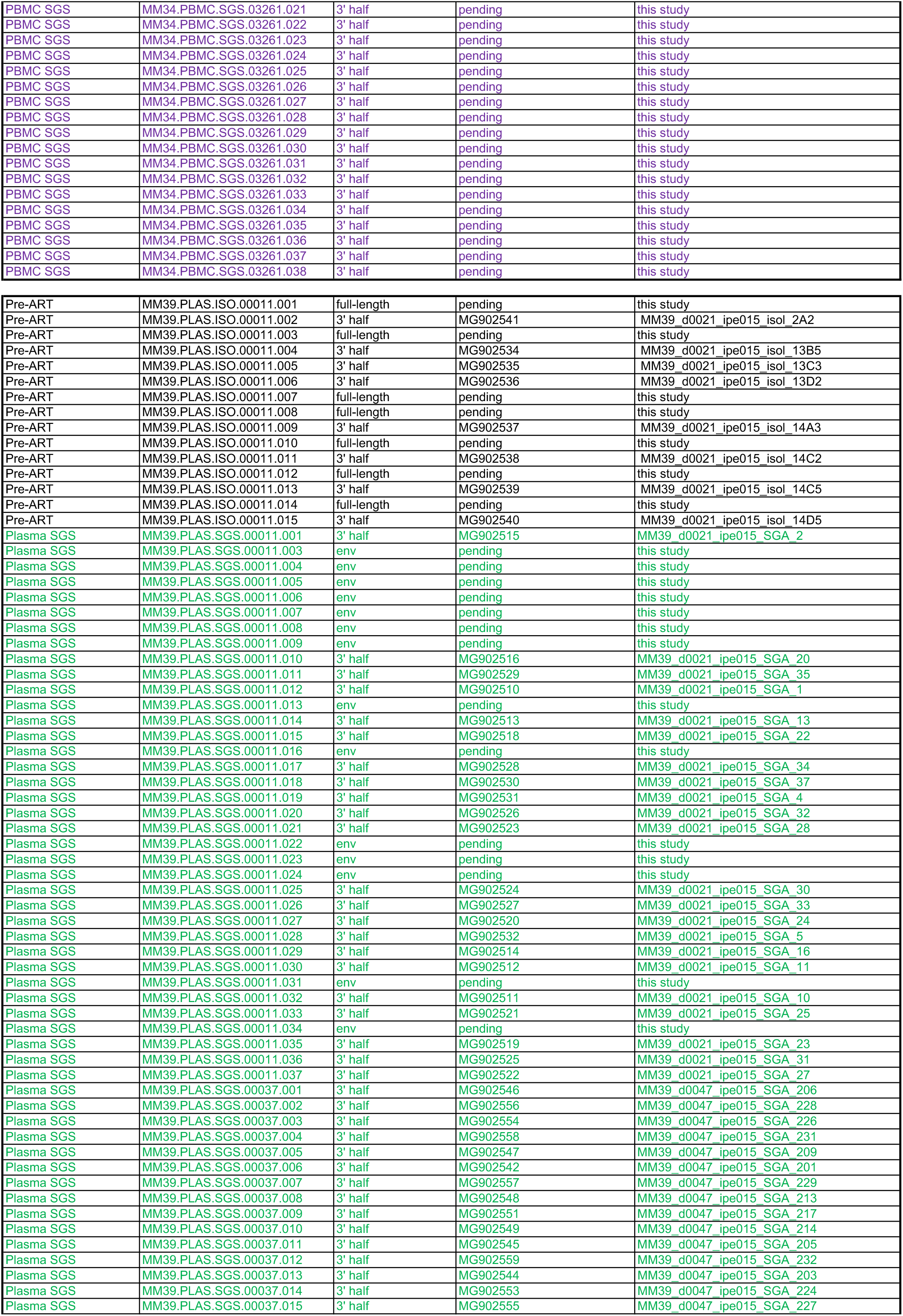
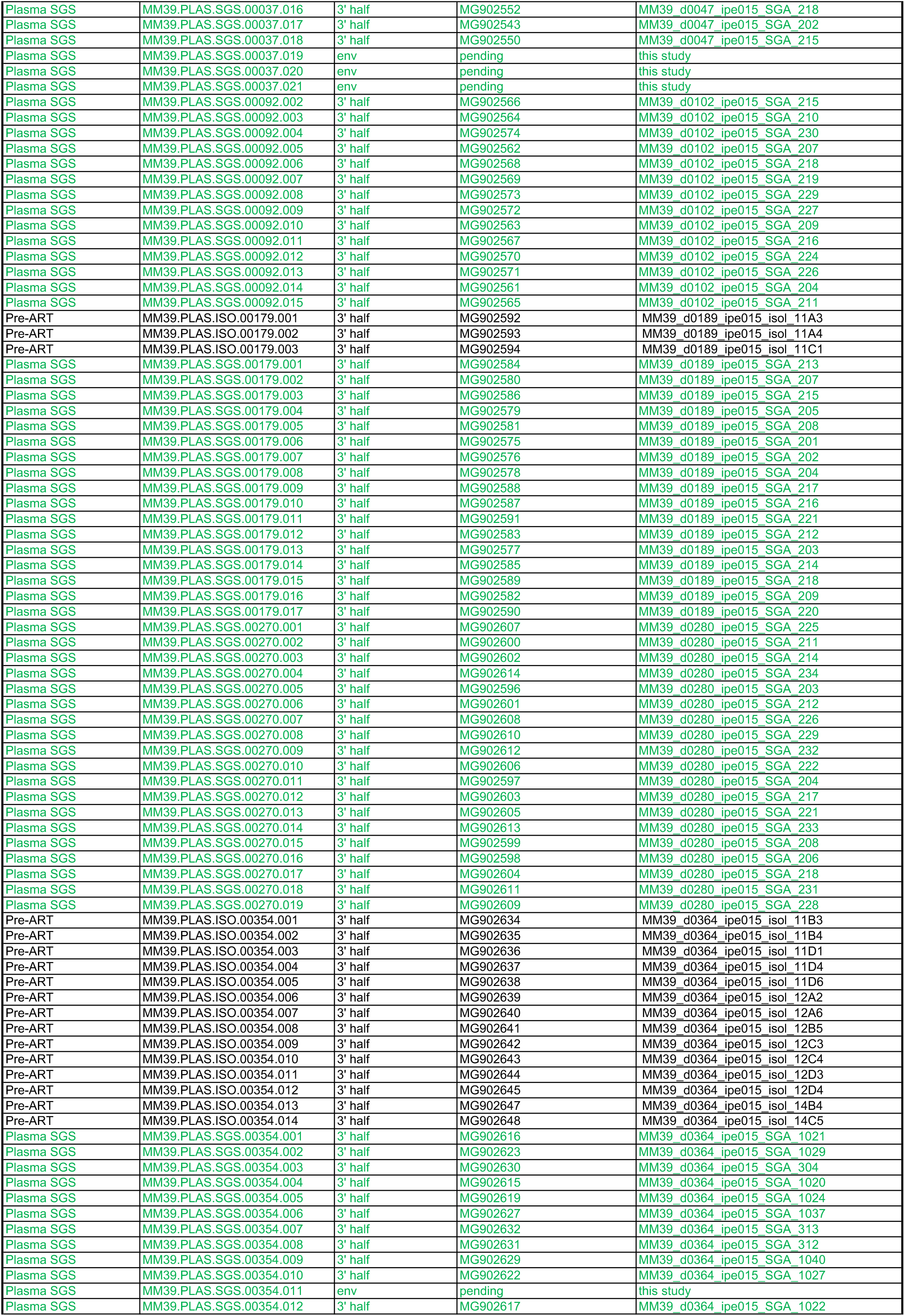
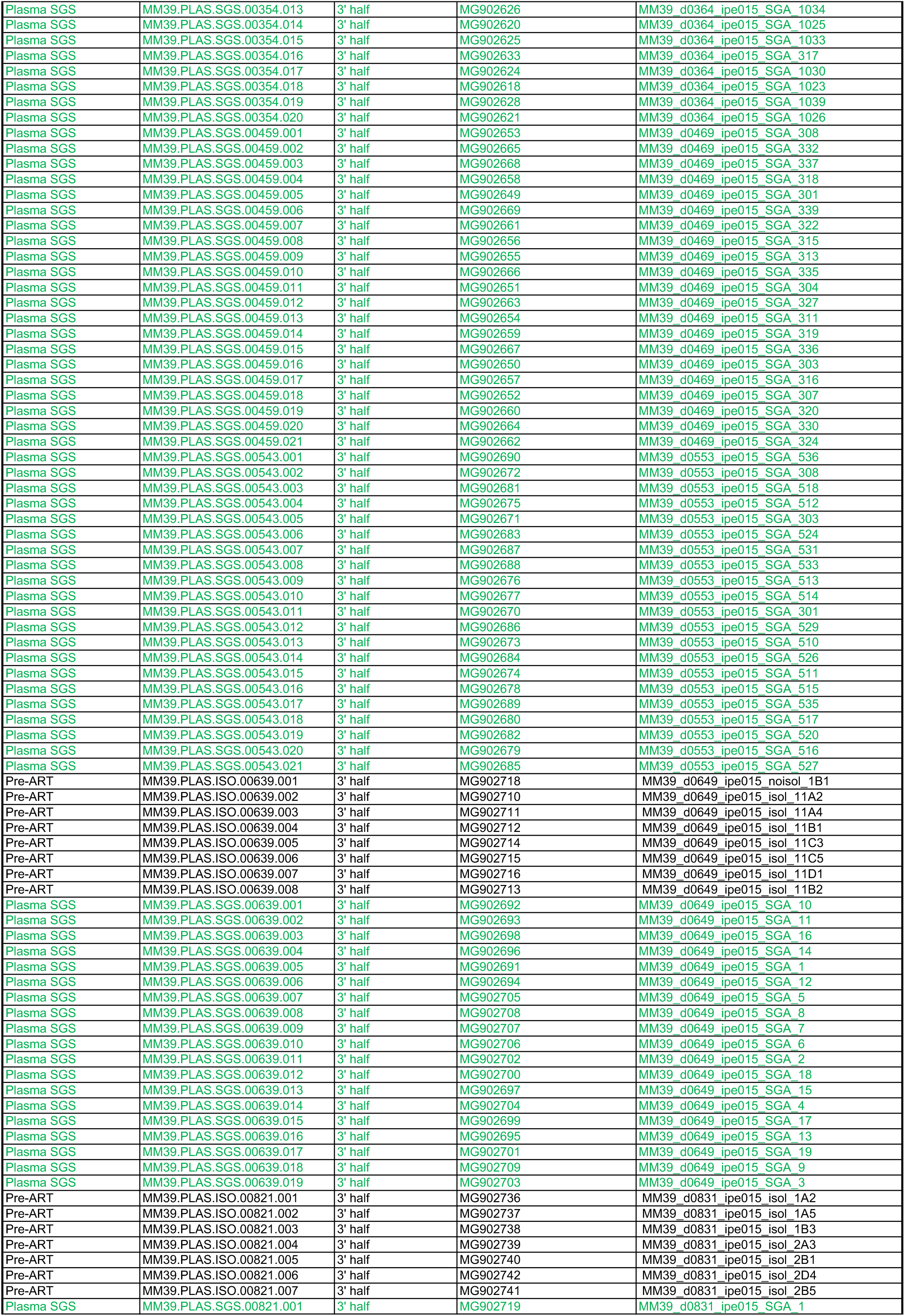
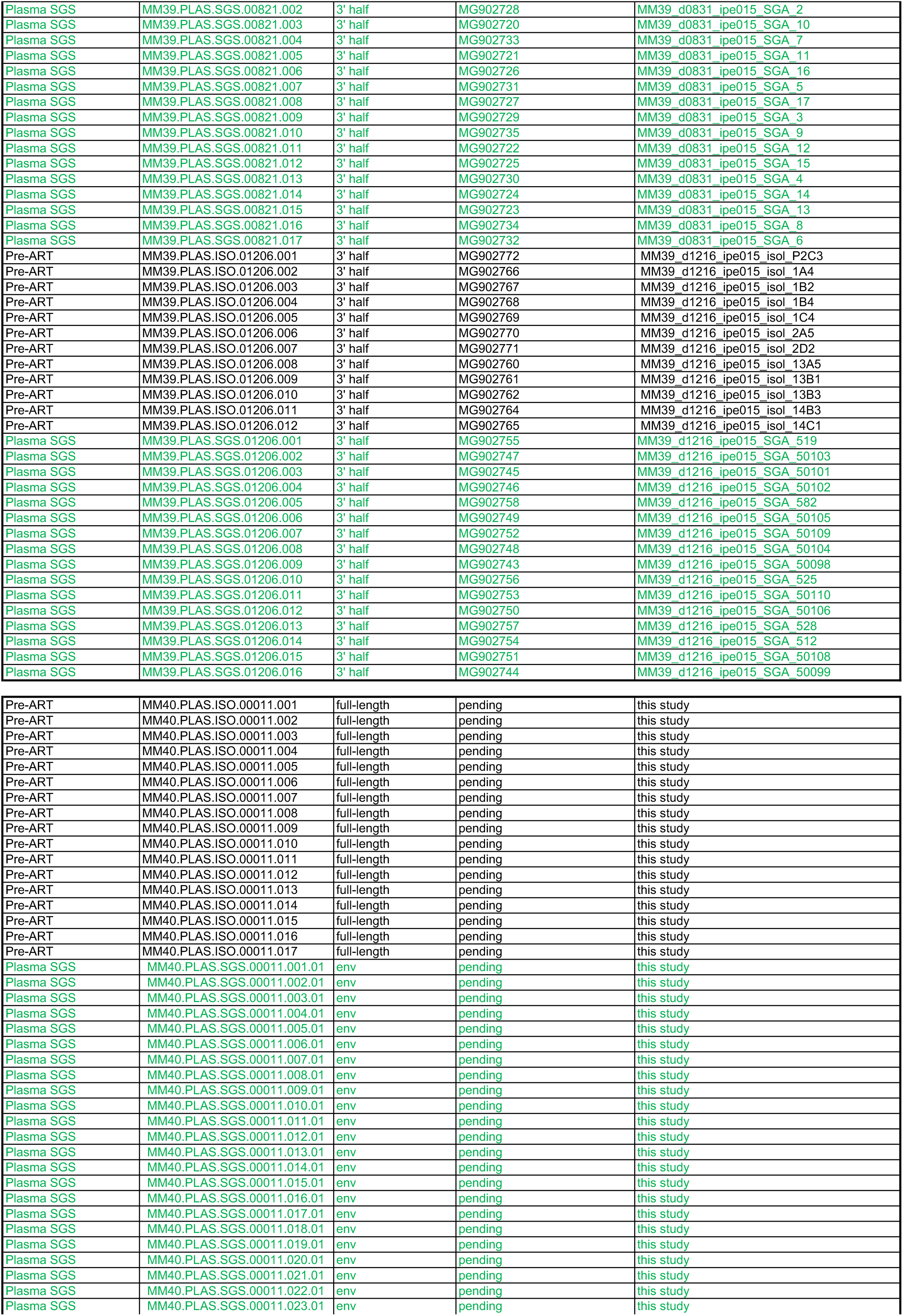
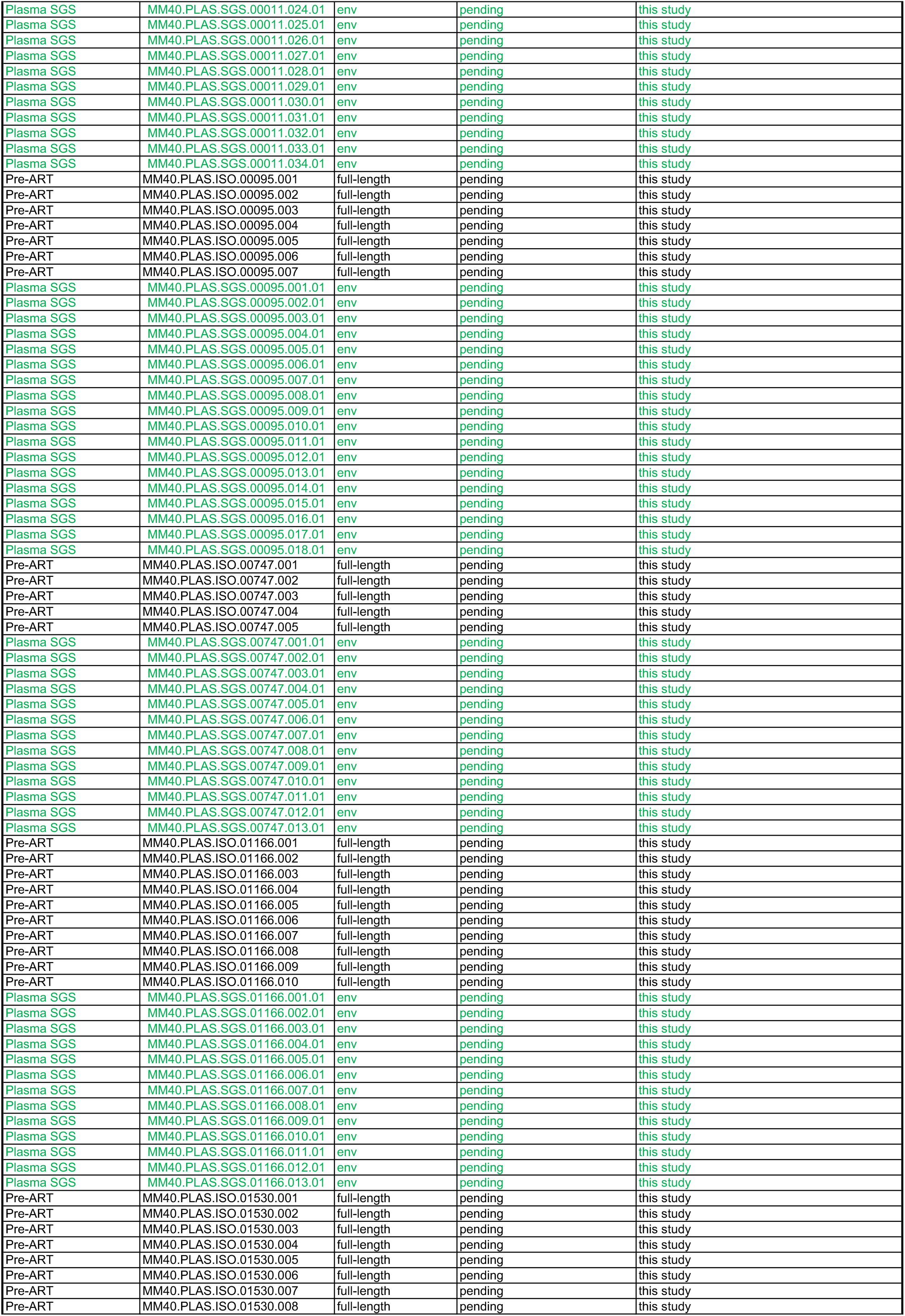
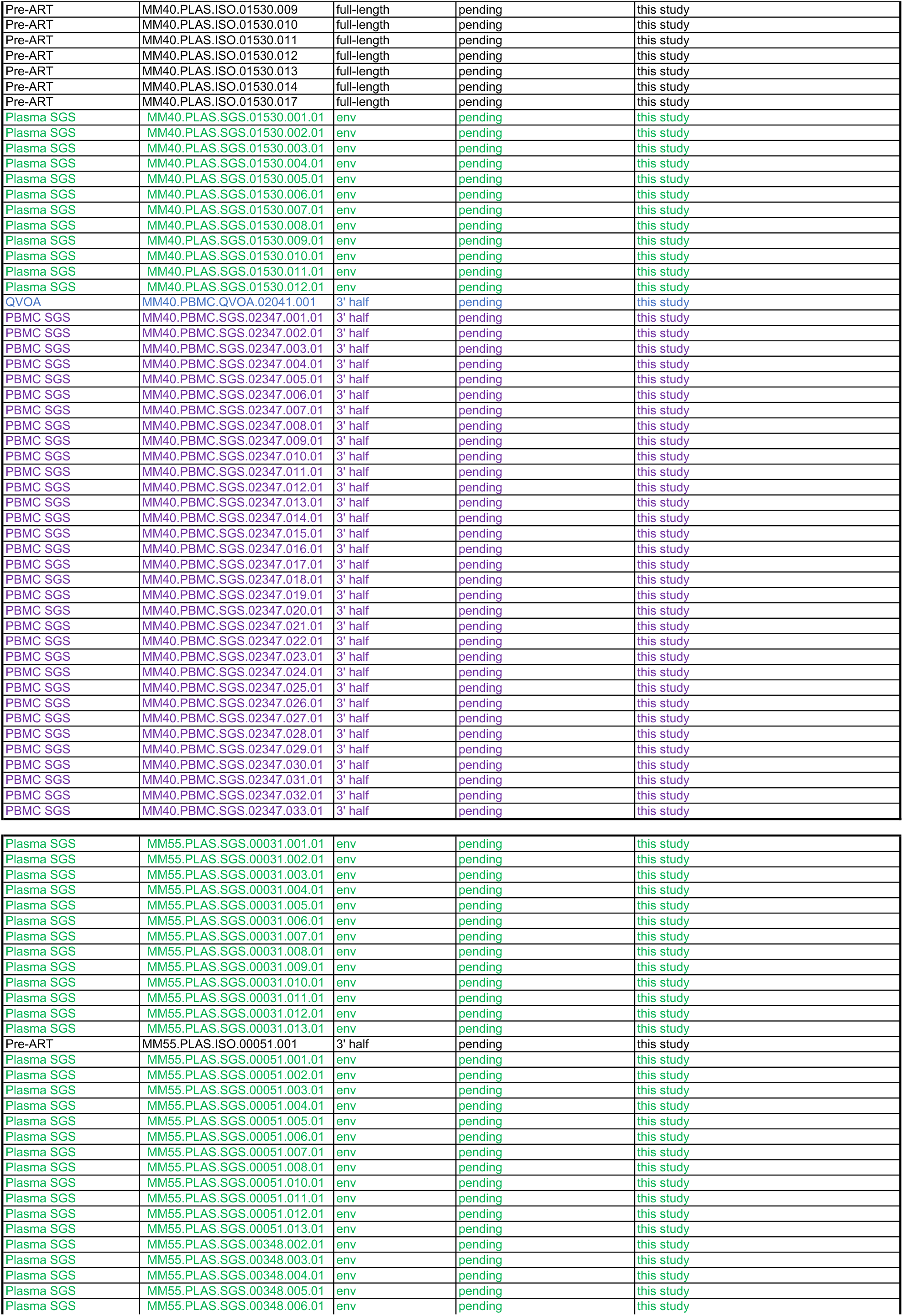
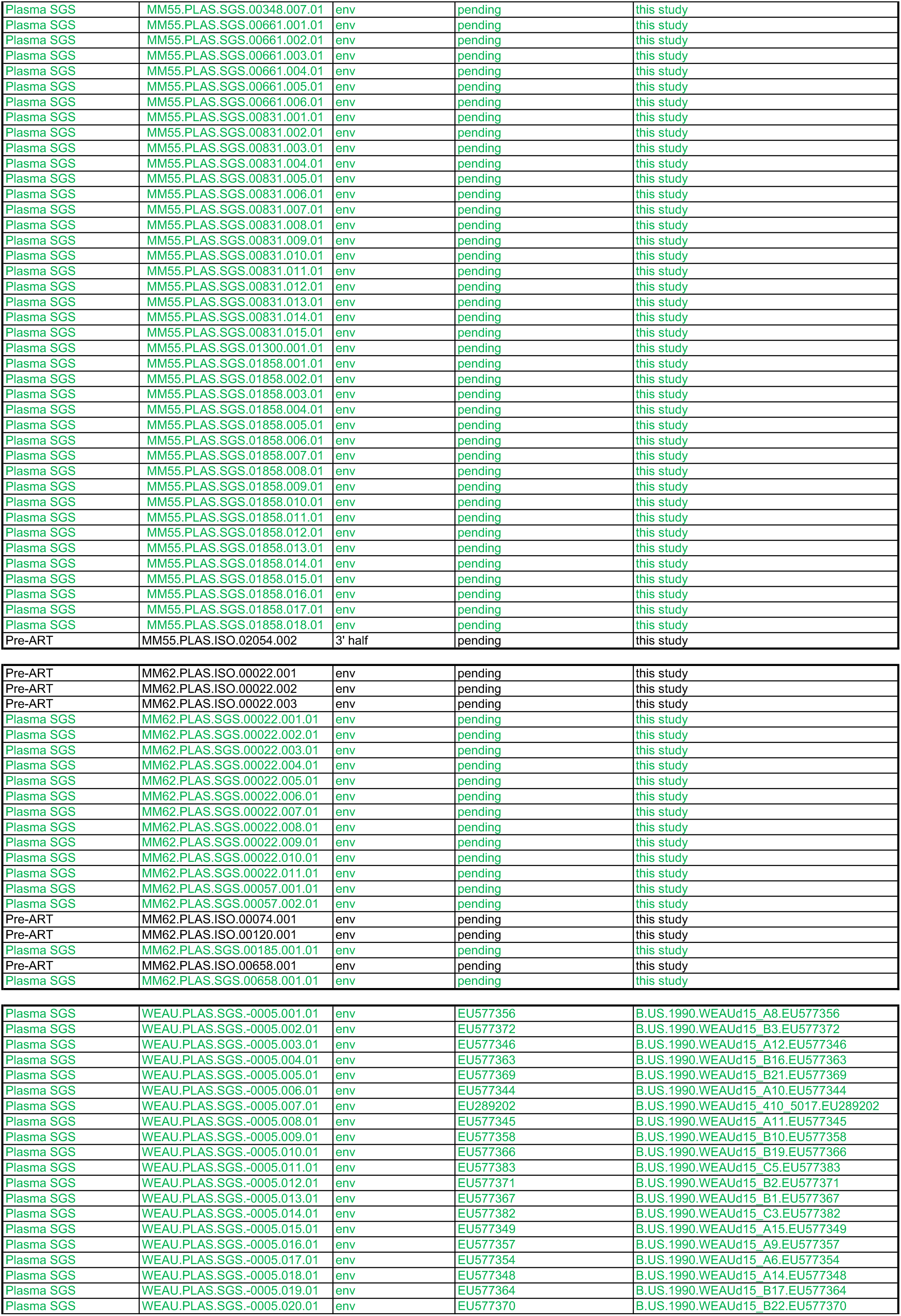
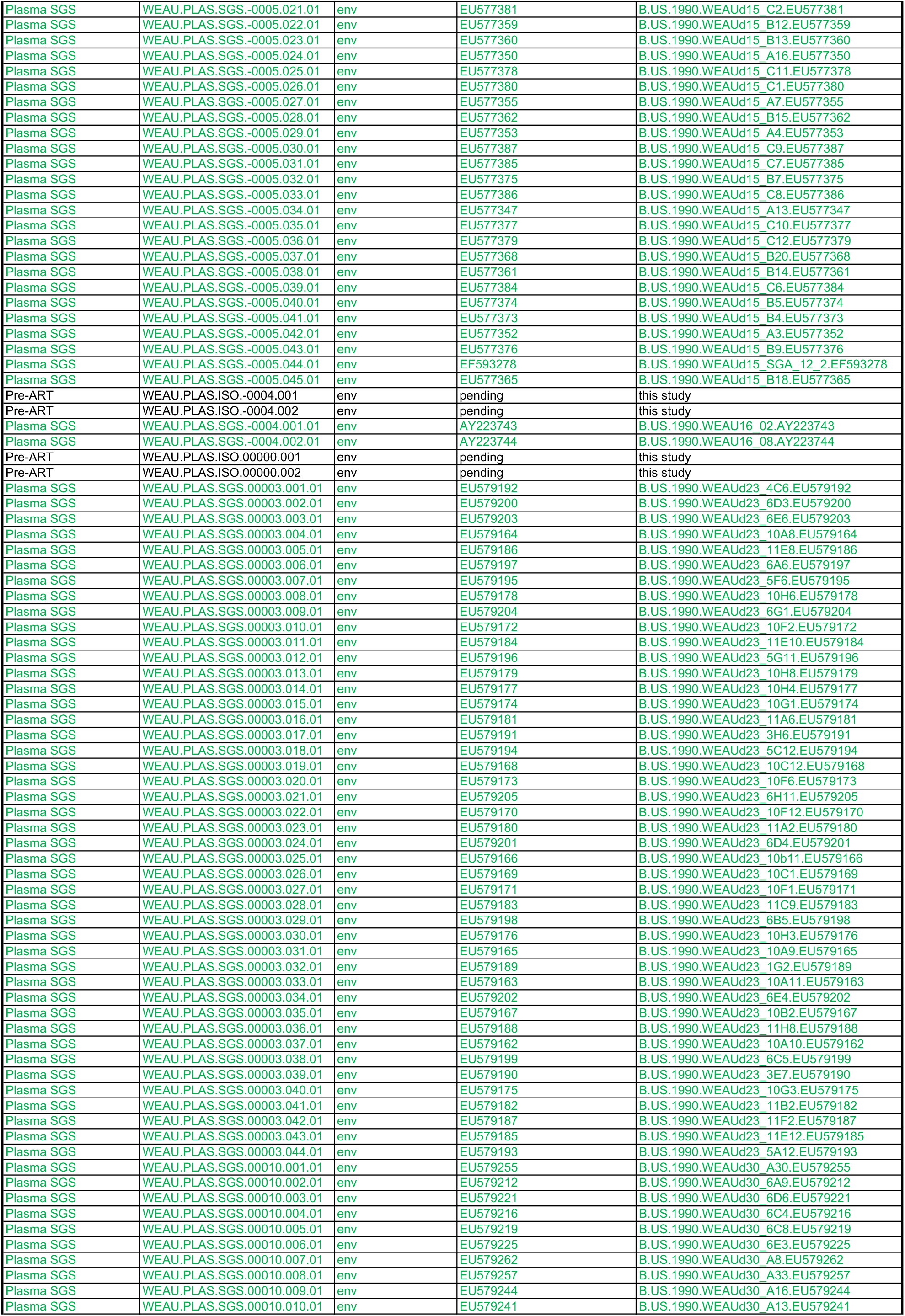
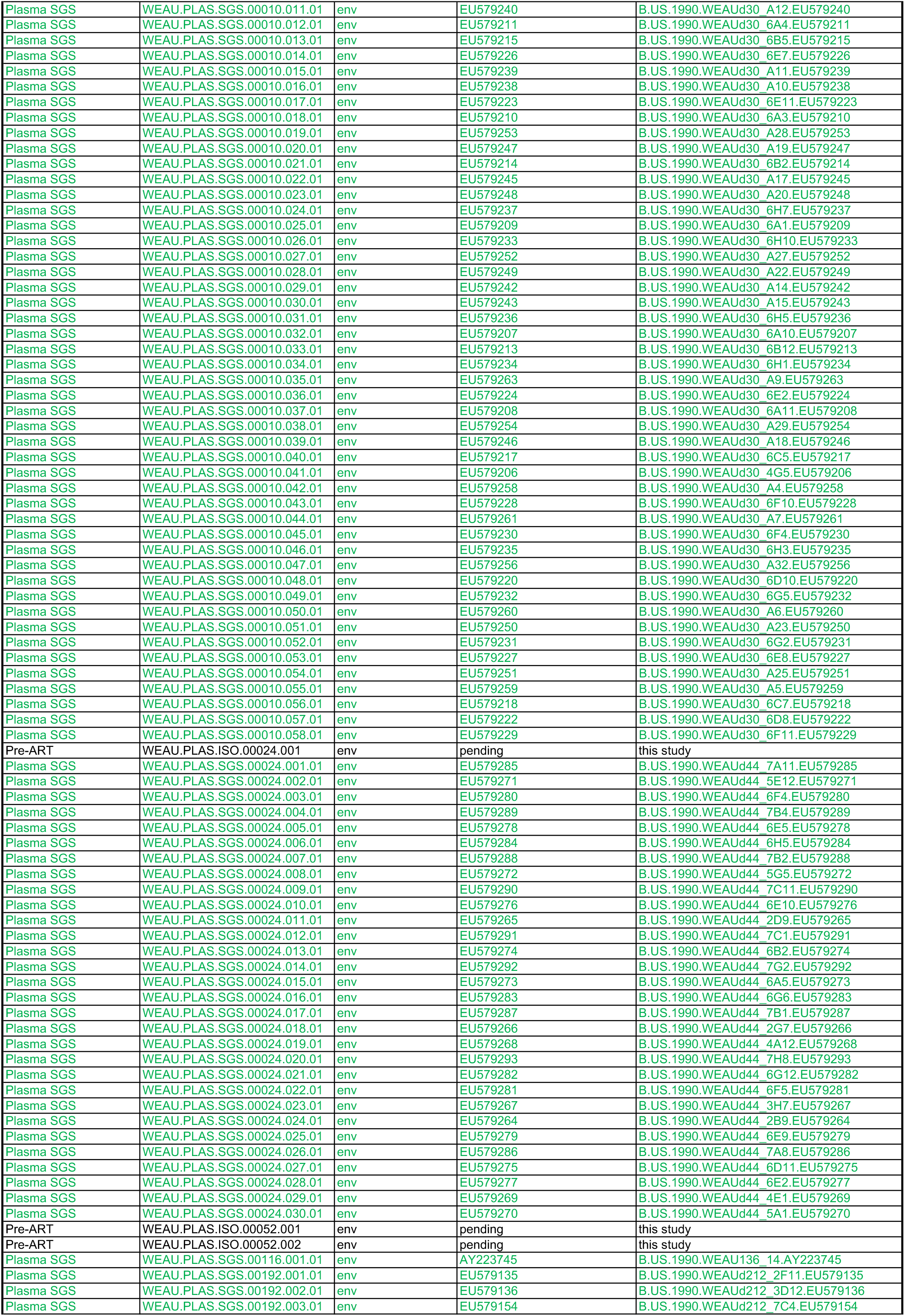
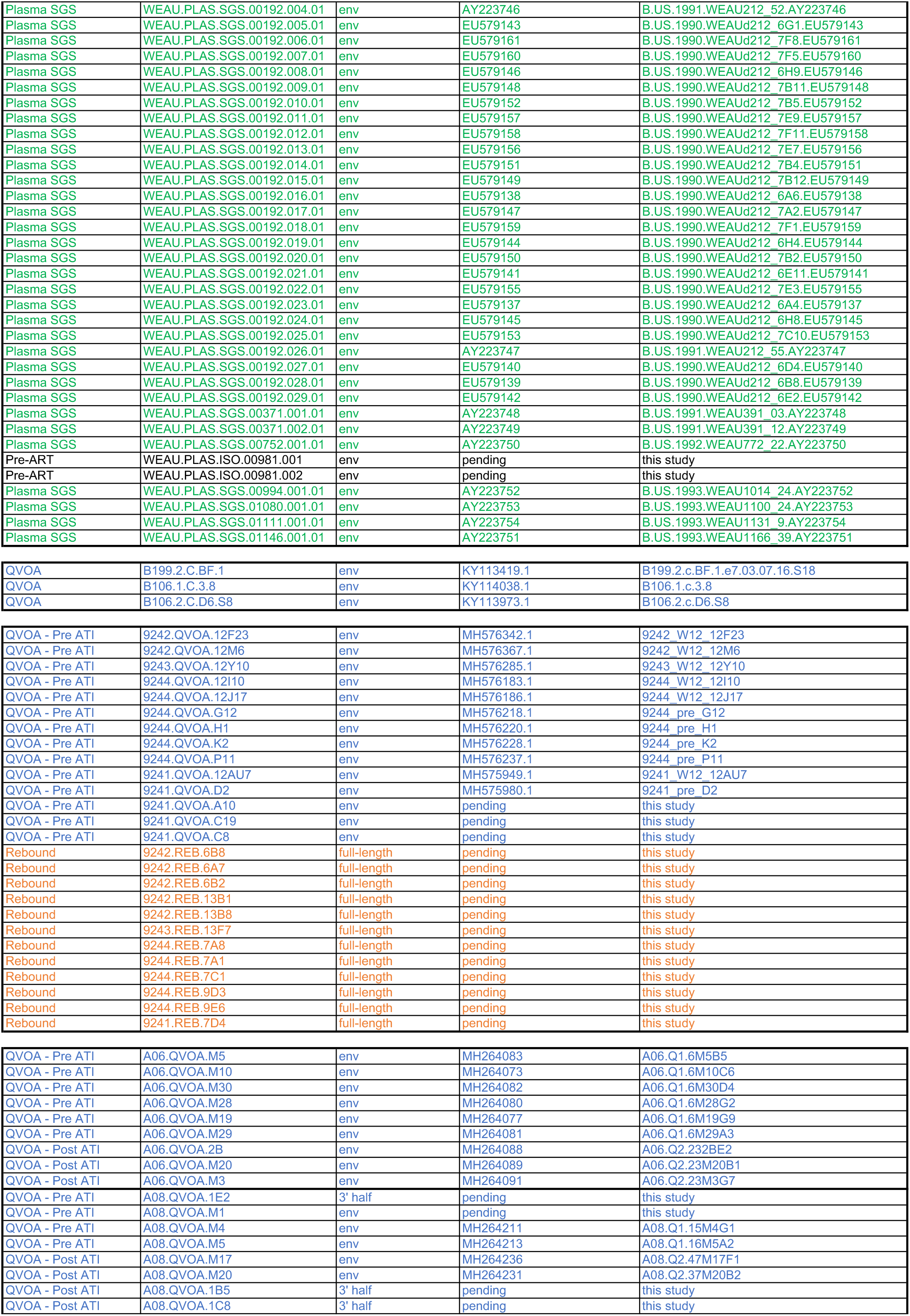
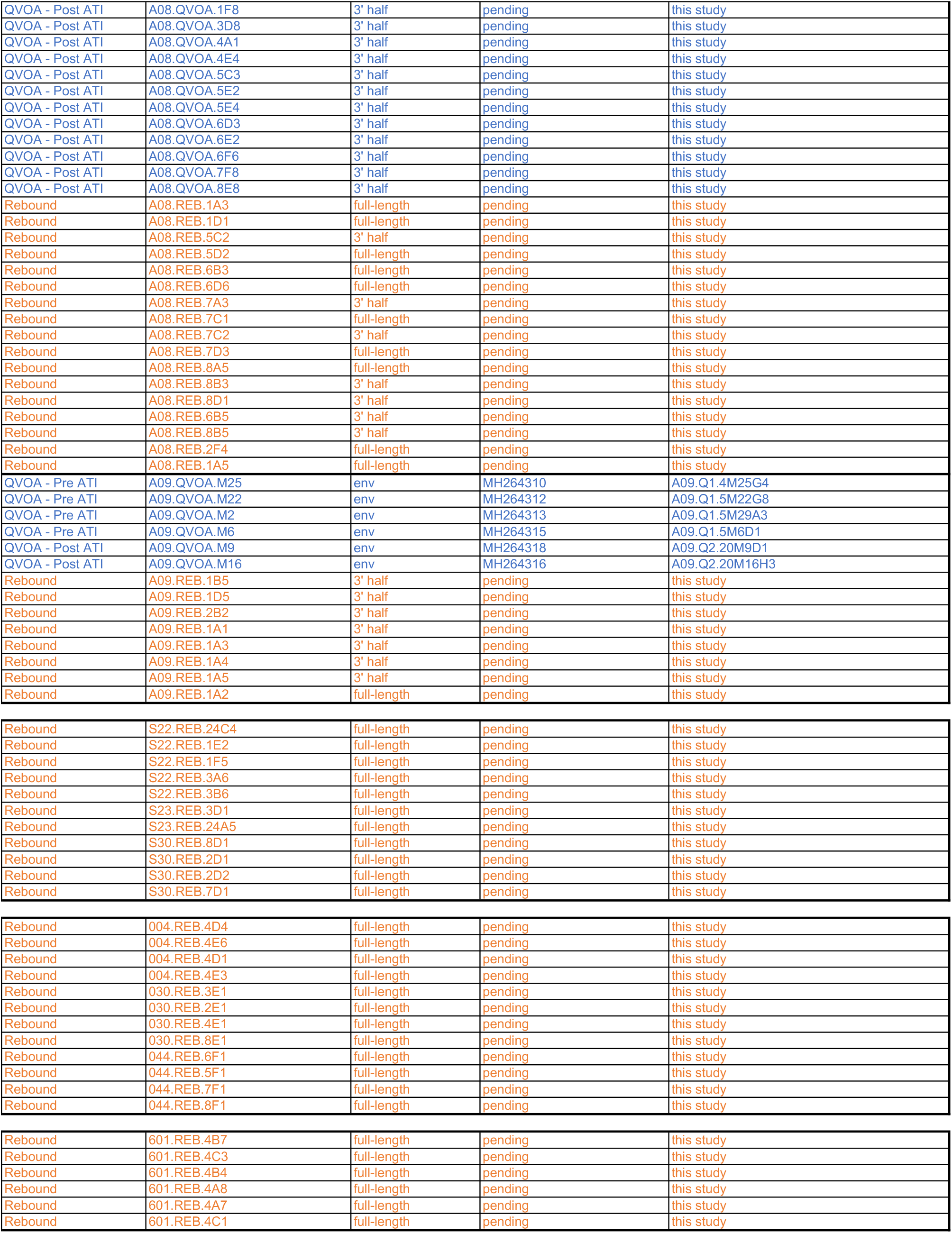
Genbank accession numbers.

## Supplementary Methods

### Bayesian models of IFN-I resistance of HIV-1 plasma isolates from longitudinally sampled participants

To create a simple model of the temporal dynamics of type I interferon (IFN-I) resistance, IFNα2 and IFNβ IC_50_ values were each modeled using a Bayesian change point hierar-chical model. The model is based on a segmented regression of the log IC_50_ making the following simplifying assumptions:

- Each participant has a level of resistance at the acute infection stage drawn from separate population-level distributions for typical, non- or fast progressors.
- Each participant has a drop (or rise) in IFN-I resistance from acute levels drawn from separate population-level distributions for typical, non- or fast progressors.
- Each participant has a time to nadir drawn from separate population-level distributions for typical, non- or fast progressors.
- Resistance changes linearly from onset of symptoms to time of nadir.
- The nadir of IFN-I resistance represents a changepoint in the data. Following this point, changes in IFN-I resistance are modeled as a linear function of CD4+ T cell count changes away from the count found at nadir.
- Where data is not present, CD4+ T cell counts are assumed to be linearly interpolated between adjacent observations.

The log IC_50_ observation from each viral isolate *i* was modeled as a normal distribution IC_50_*_i_*, ~ Normal(*μ_i_*, *σ*) with mean *μ_i_* where:

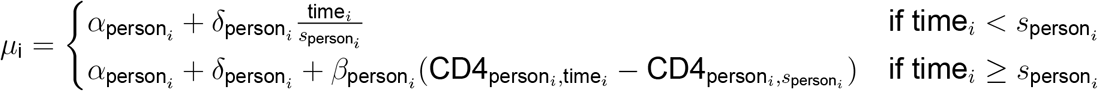

where the parameters *α_j_* represent the level of IFN-I resistance at symptom onset, *δ_j_* represents the change from symptom onset to nadir and *s_j_* represents the time of nadir in person *j*. Study participant data is represented by time» corresponding to the time since onset of symptoms and person*_i_* recording the participant from which isolate *i* was collected, CD4*_j,k_* containing the estimated CD4+ T cell count for person *j* at time *k* and progression*_j_* is the disease progression type (fast/non/typical) for participant *j*. The hierarchical probabilities for these parameters were:

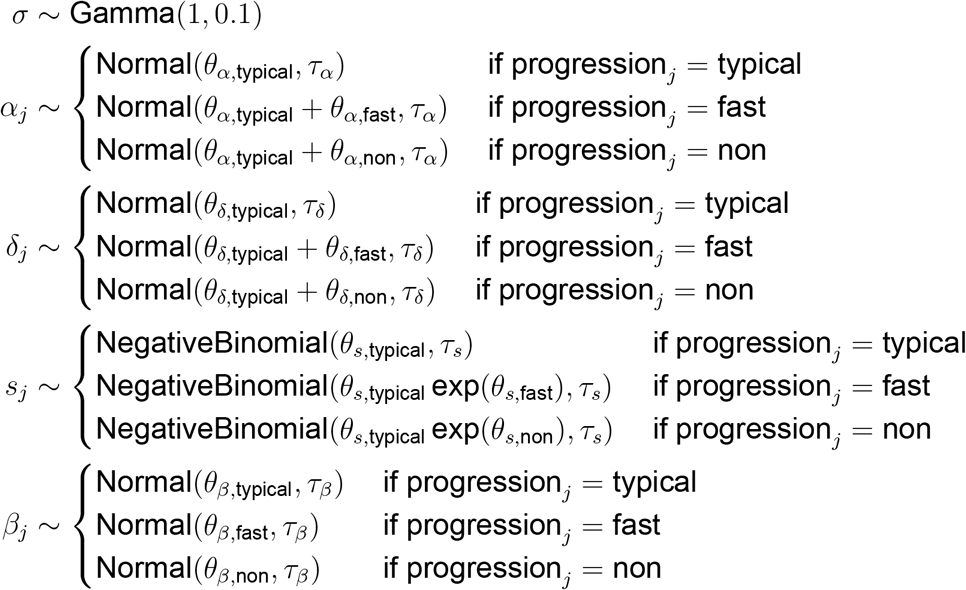

where *j* indicates each participant and NegativeBinomial(x,y) represents a negative binomial distribution parameterized such that the expected value is *x* and the variance is 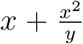. All hyperparameters were given prior probabilities of *θ_x_* ~ Normal(0,10) for parameters representing the means of a distribution and *τ_x_ -* Gamma(1,0.1) for parameters representing standard deviations other than *θ_α,typical_* and *θ_s,typical_* which were given a flat prior and *τ_s_ -* Cauchy(0,10).

For computational efficiency, the nadir time parameter *s* was discretized to weekly intervals, assumed to fall within 1 to 150 weeks after symptom onset and marginalized out of the joint probability:

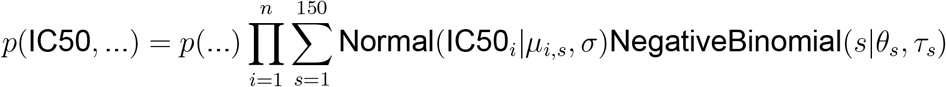

where … represents all parameters other than *s* and *μ_i,s_* is defined the same as *μ_i_*:

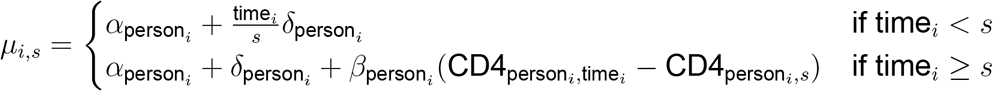

Posterior probabilities were estimated with 50 Markov chain Monte Carlo chains of 5000 iterations each using Stan (91).

### Bayesian models of IFN-I resistance of outgrowth and rebound HIV-1 isolates

To compare the IFN-I resistance of viral isolates derived from plasma samples collected during acute, chronic and rebound infections, as well as from viably frozen PBMCs collected during ART suppression (QVOA), IFNα2 and IFNβ IC_50_ values were modeled using a Bayesian hierarchical model. The model is based on the assumptions that:

- Isolates found at acute infection form a base level of IFN-I resistance for a given person. Resistances in virus isolated from chronic, ART suppressed and rebound infection for this person are modelled as changes from this initial level.
- The mean IC_50_ level within each person for acute isolates and the change from acute levels for chronic, QVOA and rebound isolates are drawn from a population-level distribution for that type.
- QVOA isolates are separated into to two populations; a “pre” group composed of QVOA viruses isolated from study participants prior to or in the absence of treatment interruption (ATI) and a “post” group of QVOA viruses isolated from participants following ATI and reinitiation of ART
- In both QVOA populations, the viruses can include some proportion of rebound-like isolates. This mixture is modeled in both pre- and post-treatment so that differences in mixture proportion between the two populations can be assessed.
- Variation in the potency of INF-I used to experimentally determine IC_50_ values may shift the inferred resistance for isolates tested in other studies. This effect is modeled as a multiplicative shift in IC_50_ for all isolates measured outside this study (acute recipient and chronic donor isolates from ref. *50*).
- Isolates from participants who received exogenous IFNα2 during treatment interruption may display altered interferon resistance. This effect is modeled as a multiplicative shift in IC_50_ for all rebound isolates from such participants (participants 004, 030, and 044 from ref. 38).

The log IC_50_ observation from each viral isolate *i* from acute, chronic and rebound isolates was modeled as a normal distribution:

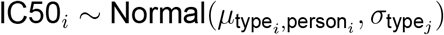

with the mean resistance for isolate type *j* from person *k*:

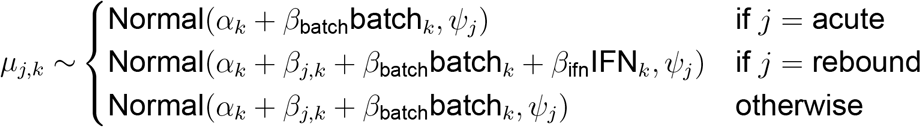

where type*_i_* indicates whether isolate *i* was isolated during acute, chronic, QVOA or rebound infection from participant person*_i_*, batch*_k_* indicates when isolates from person *k* were tested in another study and IFN_k_ indicates when person *k* was treated with exogenous IFNα2 prior to and during treatment interruption. Parameters are included for the mean resistance level during acute infection for each person *α_k_*, standard deviation of isolates of type *j* within a person *σ_j_*, standard deviation of mean resistance for type *j* isolates among people *ψ_j_*, change from acute levels in isolates of type *j* in a given participant *β_j,k_* the effects of exogenous IFN treatment *β_ifn_* and batch to batch variation in IFN in isolates assayed in previous studies *β_batch_*.

For QVOA isolates, the IC_50_ was modeled as a mixture of two populations such that:

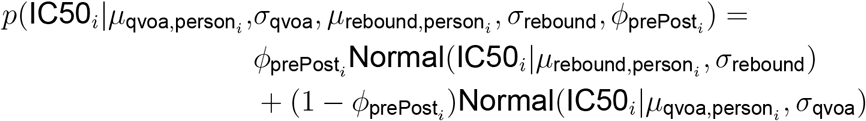

where prePost*_i_* indicates whether isolate *i* was isolated pre- or post-ATI and and *ϕ*_pre_ and *ϕ*_post_ represent the proportion of rebound-like virus present in pre- and post-ATI QVOA isolates.

The hierarchical parameter priors were modeled as:

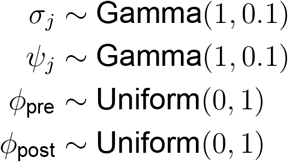

where *j* indicates the isolate type (acute, chronic, QVOA, rebound). All *α_k_*, *β_j,k_* were given flat priors and *β*_ifn_ ~ Normal(0,10) and *β*_batch_ ~ Normal(0,10).

Posterior probabilities were estimated with 50 Markov chain Monte Carlo chains of 5,000 iterations each using Stan (*91*).

## References and Notes

1. D. B. Stetson, R. Medzhitov, Type I interferons in host defense. Immunity 25, 373–381 (2006). https://doi.org/10.1016/j.immuni.2006.08.007.

2. S. Pestka, C. D. Krause, M. R. Walter, Interferons, interferon-like cytokines, and their receptors. Immunol Rev 202, 8–32 (2004). https://doi.org/10.1111/j-0105-2896.2004.00204.x.

3. H. M. Lazear, J. W. Schoggins, M. S. Diamond, Shared and distinct functions of type I and type III interferons. Immunity 50, 907–923 (2019). https://doi.org/10.1016/j.immuni.2019.03.025.

4. E. V. Mesev, R. A. LeDesma, A. Ploss, Decoding type I and III interferon signalling during viral infection. Nat Microbiol 4, 914–924 (2019). https://doi.org/10.1038/s41564-019-0421-x.

5. G. Schreiber, The molecular basis for differential type I interferon signaling. J Biol Chem 292, 7285–7294 (2017). https://doi.org/10.1074/jbc.R116.774562.

6. C. Thomas, I. Moraga, D. Levin, P. O. Krutzik, Y. Podoplelova, A. Trejo, C. Lee, G. Yarden, S. E. Vleck, J. S. Glenn, G. P. Nolan, J. Piehler, G. Schreiber, K. C. Garcia, Structural linkage between ligand discrimination and receptor activation by type I interferons. Cell 146, 621–632 (2011). https://doi.org/10.1016/j.cell.2011.06.048.

7. T. Doyle, C. Goujon, M. H. Malim, HIV-1 and interferons: who’s interfering with whom? Nat Rev Microbiol 13, 403–413 (2015). https://doi.org/10.1038/nrmicro3449.

8. J. W. Schoggins, S. J. Wilson, M. Panis, M. Y. Murphy, C. T. Jones, P. Bieniasz, C. M. Rice, A diverse range of gene products are effectors of the type I interferon antiviral response. Nature 472, 481–485 (2011). https://doi.org/10.1038/nature09907.

9. C. M. Berry, Understanding Interferon Subtype Therapy for Viral Infections: Harnessing the power of the innate immune system. Cytokine Growth Factor Rev 31, 83–90 (2016). https://doi.org/10.1016/j.cytogfr.2016.08.001.

10. T. B. Lavoie, E. Kalie, S. Crisafulli-Cabatu, R. Abramovich, G. DiGioia, K. Moolchan, S. Pestka, G. Schreiber, Binding and activity of all human alpha interferon subtypes. Cytokine 56, 282–289 (2011). https://doi.org/10.1016/j.cyto.2011.07.019.

11. H. P. Moll, T. Maier, A. Zommer, T. Lavoie, C. Brostjan, The differential activity of interferon-alpha subtypes is consistent among distinct target genes and cell types. Cytokine 53, 52–59 (2011). https://doi.org/10.1016/j.cyto.2010.09.006.

12. R. Szubin, W. L. Chang, T. Greasby, L. Beckett, N. Baumgarth, Rigid interferon-alpha subtype responses of human plasmacytoid dendritic cells. J Interferon Cytokine Res 28, 749–763 (2008). https://doi.org/10.1089/jir.2008.0037.

13. P. Borrow, Innate immunity in acute HIV-1 infection. Curr Opin HIV AIDS 6, 353–363 (2011). https://doi.org/10.1097/COH.0b013e3283495996.

14. S. E. Bosinger, N. S. Utay, Type I interferon: understanding its role in HIV pathogenesis and therapy. Curr HIV/AIDS Rep 12, 41–53 (2015). https://doi.org/10.1007/s11904-014-0244-6.

15. K. Nganou-Makamdop, D. C. Douek, Manipulating the interferon signaling pathway: Implications for HIV infection. Virol Sin 34, 192–196 (2019). https://doi.org/10.1007/s12250-019-00085-5.

16. N. S. Utay, D. C. Douek, Interferons and HIV infection: The Good, the Bad, and the Ugly. Pathog Immun 1, 107–116 (2016). https://doi.org/10.20411/pai.v1i1.125.

17. S. E. Bosinger, D. L. Sodora, G. Silvestri, Generalized immune activation and innate immune responses in simian immunodeficiency virus infection. Curr Opin HIV AIDS 6, 411–418 (2011). https://doi.org/10.1097/COH.0b013e3283499cf6.

18. A. R. Stacey, P. J. Norris, L. Qin, E. A. Haygreen, E. Taylor, J. Heitman, M. Lebedeva, A. DeCamp, D. Li, D. Grove, S. G. Self, P. Borrow, Induction of a striking systemic cytokine cascade prior to peak viremia in acute human immunodeficiency virus type 1 infection, in contrast to more modest and delayed responses in acute hepatitis B and C virus infections. J Virol 83, 3719–3733 (2009). https://doi.org/10.1128/JVI.01844-08.

19. K. Abel, D. M. Rocke, B. Chohan, L. Fritts, C. J. Miller, Temporal and anatomic relationship between virus replication and cytokine gene expression after vaginal simian immunodeficiency virus infection. J Virol 79, 12164–12172 (2005). https://doi.org/10.1128/JVI.79.19.12164-12172.2005.

20. Q. Li, J. D. Estes, P. M. Schlievert, L. Duan, A. J. Brosnahan, P. J. Southern, C. S. Reilly, M. L. Peterson, N. Schultz-Darken, K. G. Brunner, K. R. Nephew, S. Pambuccian, J. D. Lifson, J. V. Carlis, A. T. Haase, Glycerol monolaurate prevents mucosal SIV transmission. Nature 458, 1034–1038 (2009). https://doi.org/10.1038/nature07831.

21. N. G. Sandler, S. E. Bosinger, J. D. Estes, R. T. Zhu, G. K. Tharp, E. Boritz, D. Levin, S. Wijeyesinghe, K. N. Makamdop, G. Q. del Prete, B. J. Hill, J. K. Timmer, E. Reiss, G. Yarden, S. Darko, E. Contijoch, J. P. Todd, G. Silvestri, M. Nason, R. B. Norgren, Jr., B. F. Keele, S. Rao, J. A. Langer, J. D. Lifson, G. Schreiber, D. C. Douek, Type I interferon responses in rhesus macaques prevent SIV infection and slow disease progression. Nature 511, 601–605 (2014). https://doi.org/10.1038/nature13554.

22. R. S. Veazey, H. A. Pilch-Cooper, T. J. Hope, G. Alter, A. M. Carias, M. Sips, X. Wang, B. Rodriguez, S. F. Sieg, A. Reich, P. Wilkinson, M. J. Cameron, M. M. Lederman, Prevention of SHIV transmission by topical IFN-beta treatment. Mucosal Immunol 9, 1528–1536 (2016). https://doi.org/10.1038/mi.2015.146.

23. D. Carnathan, B. Lawson, J. Yu, K. Patel, J. M. Billingsley, G. K. Tharp, O. M. Delmas, R. Dawoud, P. Wilkinson, C. Nicolette, M. J. Cameron, R. P. Sekaly, S. E. Bosinger, G. Silvestri, T. H. Vanderford, Reduced chronic lymphocyte activation following interferon alpha blockade during the acute phase of simian immunodeficiency virus infection in rhesus macaques. J Virol 92, (2018). https://doi.org/10.1128/JVI.01760-17.

24. G. A. Hardy, S. Sieg, B. Rodriguez, D. Anthony, R. Asaad, W. Jiang, J. Mudd, T. Schacker, N. T. Funderburg, H. A. Pilch-Cooper, R. Debernardo, R. L. Rabin, M. M. Lederman, C. V. Harding, Interferon-alpha is the primary plasma type-I IFN in HIV-1 infection and correlates with immune activation and disease markers. PLoS One 8, e56527 (2013). https://doi.org/10.1371/journal.pone.0056527.

25. A. R. Sedaghat, J. German, T. M. Teslovich, J. Cofrancesco, Jr., C. C. Jie, C. C. Talbot, Jr., R. F. Siliciano, Chronic CD4+ T-cell activation and depletion in human immunodeficiency virus type 1 infection: type I interferon-mediated disruption of T-cell dynamics. J Virol 82, 1870–1883 (2008). https://doi.org/10.1128/JVI.02228-07.

26. H. Rempel, B. Sun, C. Calosing, S. K. Pillai, L. Pulliam, Interferon-alpha drives monocyte gene expression in chronic unsuppressed HIV-1 infection. AIDS 24, 1415–1423 (2010). https://doi.org/10.1097/QAD.0b013e32833ac623.

27. S. Fernandez, S. Tanaskovic, K. Helbig, R. Rajasuriar, M. Kramski, J. M. Murray, M. Beard, D. Purcell, S. R. Lewin, P. Price, M. A. French, CD4+ T-cell deficiency in HIV patients responding to antiretroviral therapy is associated with increased expression of interferon-stimulated genes in CD4+ T cells. J Infect Dis 204, 1927–1935 (2011). https://doi.org/10.1093/infdis/jir659.

28. G. Silvestri, D. L. Sodora, R. A. Koup, H. M. McClure, S. I. Staprans, M. B. Feinberg, Nonpathogenic SIV infection of sooty mangabeys Is characterized by limited bystander immunopathology despite chronic high-level viremia. Immunity 18, 441–452 (2003). https://doi.org/10.1016/S1074-7613(03)00060-8.

29. S. M. Dillon, K. Guo, G. L. Austin, S. Gianella, P. A. Engen, E. A. Mutlu, J. Losurdo, G. Swanson, P. Chakradeo, A. Keshavarzian, A. L. Landay, M. L. Santiago, C. C. Wilson, A compartmentalized type I interferon response in the gut during chronic HIV-1 infection is associated with immunopathogenesis. AIDS 32, 1599–1611 (2018). https://doi.org/10.1097/QAD.0000000000001863.

30. J. P. Herbeuval, A. Boasso, J. C. Grivel, A. W. Hardy, S. A. Anderson, M. J. Dolan, C. Chougnet, J. D. Lifson, G. M. Shearer, TNF-related apoptosis-inducing ligand (TRAIL) in HIV-1-infected patients and its in vitro production by antigen-presenting cells. Blood 105, 2458–2464 (2005). https://doi.org/10.1182/blood-2004-08-3058.

31. D. Sauce, C. Elbim, V. Appay, Monitoring cellular immune markers in HIV infection: from activation to exhaustion. Curr Opin HIV AIDS 8, 125–131 (2013). https://doi.org/10.1097/C0H.0b013e32835d08a9.

32. S. Lederer, D. Favre, K. A. Walters, S. Proll, B. Kanwar, Z. Kasakow, C. R. Baskin, R. Palermo, J. M. McCune, M. G. Katze, Transcriptional profiling in pathogenic and non-pathogenic SIV infections reveals significant distinctions in kinetics and tissue compartmentalization. PLoS Pathog 5, e1000296 (2009). https://doi.org/10.1371/journal.ppat.1000296.

33. S. E. Bosinger, Q. Li, S. N. Gordon, N. R. Klatt, L. Duan, L. Xu, N. Francella, A. Sidahmed, A. J. Smith, E. M. Cramer, M. Zeng, D. Masopust, J. V. Carlis, L. Ran, T. H. Vanderford, M. Paiardini, R. B. Isett, D. A. Baldwin, J. G. Else, S. I. Staprans, G. Silvestri, A. T. Haase, D. J. Kelvin, Global genomic analysis reveals rapid control of a robust innate response in SIV-infected sooty mangabeys. J Clin Invest 119, 3556–3572 (2009). https://doi.org/10.1172/JCI40115.

34. B. Jacquelin, V. Mayau, B. Targat, A. S. Liovat, D. Kunkel, G. Petitjean, M. A. Dillies, P. Roques, C. Butor, G. Silvestri, L. D. Giavedoni, P. Lebon, F. Barre-Sinoussi, A. Benecke, M. C. Muller-Trutwin, Nonpathogenic SIV infection of African green monkeys induces a strong but rapidly controlled type I IFN response. J Clin Invest 119, 3544–3555 (2009). https://doi.org/10.1172/JCI40093.

35. P. W. Hunt, HIV and inflammation: mechanisms and consequences. Curr HIV/AIDS Rep 9, 139-147 (2012). https://doi.org/10.1007/s11904-012-0118-8.

36. A. Soper, I. Kimura, S. Nagaoka, Y. Konno, K. Yamamoto, Y. Koyanagi, K. Sato, Type I interferon responses by HIV-1 infection: Association with disease progression and control. Front Immunol 8, 1823 (2017). https://doi.org/10.3389/fimmu.2017.01823.

37. D. M. Asmuth, R. L. Murphy, S. L. Rosenkranz, J. J. Lertora, S. Kottilil, Y. Cramer, E. S. Chan, R. T. Schooley, C. R. Rinaldo, N. Thielman, X. D. Li, S. M. Wahl, J. Shore, J. Janik, R. A. Lempicki, Y. Simpson, R. B. Pollard, A. C. T. G. A. Team, Safety, tolerability, and mechanisms of antiretroviral activity of pegylated interferon Alfa-2a in HIV-1-monoinfected participants: a phase II clinical trial. J Infect Dis 201, 1686–1696 (2010). https://doi.org/10.1086/652420.

38. L. Azzoni, A. S. Foulkes, E. Papasavvas, A. M. Mexas, K. M. Lynn, K. Mounzer, P. Tebas, J. M. Jacobson, I. Frank, M. P. Busch, S. G. Deeks, M. Carrington, U. O’Doherty, J. Kostman, L. J. Montaner, Pegylated Interferon alfa-2a monotherapy results in suppression of HIV type 1 replication and decreased cell-associated HIV DNA integration. J Infect Dis 207, 213–222 (2013). https://doi.org/10.1093/infdis/jis663.

39. F. Boue, J. Reynes, C. Rouzioux, D. Emilie, F. Souala, R. Tubiana, C. Goujard, R. Lancar, D. Costagliola, Alpha interferon administration during structured interruptions of combination antiretroviral therapy in patients with chronic HIV-1 infection: INTERVAC ANRS 105 trial. AIDS 25, 115–118 (2011). https://doi.org/10.1097/QAD.0b013e328340a1e7.

40. C. Goujard, D. Emilie, C. Roussillon, V. Godot, C. Rouzioux, A. Venet, C. Colin, G. Pialoux, P. M. Girard, V. Boilet, M. L. Chaix, P. Galanaud, G. Chene, A.-I. S. Group, Continuous versus intermittent treatment strategies during primary HIV-1 infection: the randomized ANRS INTERPRIM Trial. AIDS 26, 1895–1905 (2012). https://doi.org/10.1097/QAD.0b013e32835844d9.

41. A. Zhen, V. Rezek, C. Youn, B. Lam, N. Chang, J. Rick, M. Carrillo, H. Martin, S. Kasparian, P. Syed, N. Rice, D. G. Brooks, S. G. Kitchen, Targeting type I interferon-mediated activation restores immune function in chronic HIV infection. J Clin Invest 127, 260–268 (2017). https://doi.org/10.1172/JCI89488.

42. L. Cheng, J. Ma, J. Li, D. Li, G. Li, F. Li, Q. Zhang, H. Yu, F. Yasui, C. Ye, L. C. Tsao, Z. Hu, L. Su, L. Zhang, Blocking type I interferon signaling enhances T cell recovery and reduces HIV-1 reservoirs. J Clin Invest 127,269-279 (2017). https://doi.org/10.1172/JCI90745.

43. X. Wei, J. M. Decker, S. Wang, H. Hui, J. C. Kappes, X. Wu, J. F. Salazar-Gonzalez, M. G. Salazar, J. M. Kilby, M. S. Saag, N. L. Komarova, M. A. Nowak, B. H. Hahn, P. D. Kwong, G. M. Shaw, Antibody neutralization and escape by HIV-1. Nature 422, 307–312 (2003). https://doi.org/10.1038/nature01470.

44. K. J. Bar, C. Y. Tsao, S. S. Iyer, J. M. Decker, Y. Yang, M. Bonsignori, X. Chen, K. K. Hwang, D. C. Montefiori, H. X. Liao, P. Hraber, W. Fischer, H. Li, S. Wang, S. Sterrett, B. F. Keele, V. V. Ganusov, A. S. Perelson, B. T. Korber, I. Georgiev, J. S. McLellan, J. W. Pavlicek, F. Gao, B. F. Haynes, B. H. Hahn, P. D. Kwong, G. M. Shaw, Early low-titer neutralizing antibodies impede HIV-1 replication and select for virus escape. PLoS Pathog 8, e1002721 (2012). https://doi.org/10.1371/journal.ppat.1002721.

45. P. Borrow, H. Lewicki, X. Wei, M. S. Horwitz, N. Peffer, H. Meyers, J. A. Nelson, J. E. Gairin, B. H. Hahn, M. B. Oldstone, G. M. Shaw, Antiviral pressure exerted by HIV-1-specific cytotoxic T lymphocytes (CTLs) during primary infection demonstrated by rapid selection of CTL escape virus. Nat Med 3, 205–211 (1997). https://doi.org/10.1038/nm0297-205.

46. N. Goonetilleke, M. K. Liu, J. F. Salazar-Gonzalez, G. Ferrari, E. Giorgi, V. V. Ganusov, B. F. Keele, G. H. Learn, E. L. Turnbull, M. G. Salazar, K. J. Weinhold, S. Moore, C. C. C. B, N. Letvin, B. F. Haynes, M. S. Cohen, P. Hraber, T. Bhattacharya, P. Borrow, A. S. Perelson, B. H. Hahn, G. M. Shaw, B. T. Korber, A. J. McMichael, The first T cell response to transmitted/founder virus contributes to the control of acute viremia in HIV-1 infection. J Exp Med 206, 1253–1272 (2009). https://doi.org/10.1084/jem.20090365.

47. A. S. Perelson, A. U. Neumann, M. Markowitz, J. M. Leonard, D. D. Ho, HIV-1 dynamics in vivo: virion clearance rate, infected cell life-span, and viral generation time. Science 271, 1582–1586 (1996). https://doi.org/10.1126/science.271.5255.1582.

48. B. Ramratnam, S. Bonhoeffer, J. Binley, A. Hurley, L. Zhang, J. E. Mittler, M. Markowitz, J. P. Moore, A. S. Perelson, D. D. Ho, Rapid production and clearance of HIV-1 and hepatitis C virus assessed by large volume plasma apheresis. Lancet 354, 1782–1785 (1999). https://doi.org/10.1016/S0140-6736(99)02035-8.

49. N. F. Parrish, F. Gao, H. Li, E. E. Giorgi, H. J. Barbian, E. H. Parrish, L. Zajic, S. S. Iyer, J. M. Decker, A. Kumar, B. Hora, A. Berg, F. Cai, J. Hopper, T. N. Denny, H. Ding, C. Ochsenbauer, J. C. Kappes, R. P. Galimidi, A. P. West, Jr., P. J. Bjorkman, C. B. Wilen, R. W. Doms, M. O’Brien, N. Bhardwaj, P. Borrow, B. F. Haynes, M. Muldoon, J. P. Theiler, B. Korber, G. M. Shaw, B. H. Hahn, Phenotypic properties of transmitted founder HIV-1. Proc Natl Acad Sci U S A 110, 6626–6633 (2013). https://doi.org/10.1073/pnas.1304288110.

50. S. S. Iyer, F. Bibollet-Ruche, S. Sherrill-Mix, G. H. Learn, L. Plenderleith, A. G. Smith, H. J. Barbian, R. M. Russell, M. V. Gondim, C. Y. Bahari, C. M. Shaw, Y. Li, T. Decker, B. F. Haynes, G. M. Shaw, P. M. Sharp, P. Borrow, B. H. Hahn, Resistance to type 1 interferons is a major determinant of HIV-1 transmission fitness. Proc Natl Acad Sci U S A 114, E590-E599 (2017). https://doi.org/10.1073/pnas.1620144114.

51. M. S. Kunzi, H. Farzadegan, J. B. Margolick, D. Vlahov, P. M. Pitha, Identification of human immunodeficiency virus primary isolates resistant to interferon-alpha and correlation of prevalence to disease progression. J Infect Dis 171, 822–828 (1995). https://doi.org/10.1093/infdis/171A822.

52. B. R. Edlin, M. H. St Clair, P. M. Pitha, S. M. Whaling, D. M. King, J. D. Bitran, R. A. Weinstein, In-vitro resistance to zidovudine and alpha-interferon in HIV-1 isolates from patients: correlations with treatment duration and response. Ann Intern Med 117, 457–460 (1992). https://doi.org/10.7326/0003-4819-117-6-457.

53. J. F. Salazar-Gonzalez, E. Bailes, K. T. Pham, M. G. Salazar, M. B. Guffey, B. F. Keele, C. A. Derdeyn, P. Farmer, E. Hunter, S. Allen, O. Manigart, J. Mulenga, J. A. Anderson, R. Swanstrom, B. F. Haynes, G. S. Athreya, B. T. Korber, P. M. Sharp, G. M. Shaw, B. H. Hahn, Deciphering human immunodeficiency virus type 1 transmission and early envelope diversification by single-genome amplification and sequencing. J Virol 82, 3952–3970 (2008). https://doi.org/10.1128/JVI.02660-07.

54. B. F. Keele, E. E. Giorgi, J. F. Salazar-Gonzalez, J. M. Decker, K. T. Pham, M. G. Salazar, C. Sun, T. Grayson, S. Wang, H. Li, X. Wei, C. Jiang, J. L. Kirchherr, F. Gao, J. A. Anderson, L. H. Ping, R. Swanstrom, G. D. Tomaras, W. A. Blattner, P. A. Goepfert, J. M. Kilby, M. S. Saag, E. L. Delwart, M. P. Busch, M. S. Cohen, D. C. Montefiori, B. F. Haynes, B. Gaschen, G. S. Athreya, H. Y. Lee, N. Wood, C. Seoighe, A. S. Perelson, T. Bhattacharya, B. T. Korber, B. H. Hahn, G. M. Shaw, Identification and characterization of transmitted and early founder virus envelopes in primary HIV-1 infection. Proc Natl Acad Sci U S A 105, 7552–7557 (2008). https://doi.org/10.1073/pnas.0802203105.

55. F. Gao, M. Bonsignori, H. X. Liao, A. Kumar, S. M. Xia, X. Lu, F. Cai, K. K. Hwang, H. Song, T. Zhou, R. M. Lynch, S. M. Alam, M. A. Moody, G. Ferrari, M. Berrong, G. Kelsoe, G. M. Shaw, B. H. Hahn, D. C. Montefiori, G. Kamanga, M. S. Cohen, P. Hraber, P. D. Kwong, B. T. Korber, J. R. Mascola, T. B. Kepler, B. F. Haynes, Cooperation of B cell lineages in induction of HIV-1-broadly neutralizing antibodies. Cell 158, 481–491 (2014). https://doi.org/10.1016/j.cell.2014.06.022.

56. M. Bonsignori, E. F. Kreider, D. Fera, R. R. Meyerhoff, T. Bradley, K. Wiehe, S. M. Alam, B. Aussedat, W. E. Walkowicz, K. K. Hwang, K. O. Saunders, R. Zhang, M. A. Gladden, A. Monroe, A. Kumar, S. M. Xia, M. Cooper, M. K. Louder, K. McKee, R. T. Bailer, B. W. Pier, C. A. Jette, G. Kelsoe, W. B. Williams, L. Morris, J. Kappes, K. Wagh, G. Kamanga, M. S. Cohen, P. T. Hraber, D. C. Montefiori, A. Trama, H. X. Liao, T. B. Kepler, M. A. Moody, F. Gao, S. J. Danishefsky, J. R. Mascola, G. M. Shaw, B. H. Hahn, S. C. Harrison, B. T. Korber, B. F. Haynes, Staged induction of HIV-1 glycan-dependent broadly neutralizing antibodies. Sci Transl Med 9, (2017). https://doi.org/10.1126/scitranslmed.aai7514.

57. K. Wagh, E. F. Kreider, Y. Li, H. J. Barbian, G. H. Learn, E. Giorgi, P. T. Hraber, T. G. Decker, A. G. Smith, M. V. Gondim, L. Gillis, J. Wandzilak, G. Y. Chuang, R. Rawi, F. Cai, P. Pellegrino, I. Williams, J. Overbaugh, F. Gao, P. D. Kwong, B. F. Haynes, G. M. Shaw, P. Borrow, M. S. Seaman, B. H. Hahn, B. Korber, Completeness of HIV-1 envelope glycan shield at transmission determines neutralization breadth. Cell Rep 25, 893-908 e897 (2018). https://doi.org/10.1016/j.celrep.2018.09.087.

58. A. Gelman, J. B. Carlin, H. S. Stern, D. B. Rubin, Bayesian Data Analysis, Second Edition (Texts in Statistical Science). (Chapman & Hall/CRC 2003).

59. A. E. Fenton-May, O. Dibben, T. Emmerich, H. Ding, K. Pfafferott, M. M. Aasa-Chapman, P. Pellegrino, I. Williams, M. S. Cohen, F. Gao, G. M. Shaw, B. H. Hahn, C. Ochsenbauer, J. C. Kappes, P. Borrow, Relative resistance of HIV-1 founder viruses to control by interferon-alpha. Retrovirology 10, 146 (2013). https://doi.org/10.1186/1742-4690-10-146.

60. G. M. Laird, E. E. Eisele, S. A. Rabi, J. Lai, S. Chioma, J. N. Blankson, J. D. Siliciano, R. F. Siliciano, Rapid quantification of the latent reservoir for HIV-1 using a viral outgrowth assay. PLoS Pathog 9, e1003398 (2013). https://doi.org/10.1371/journal.ppat.1003398.

61. J. C. Lorenzi, Y. Z. Cohen, L. B. Cohn, E. F. Kreider, J. P. Barton, G. H. Learn, T. Oliveira, C. L. Lavine, J. A. Horwitz, A. Settler, M. Jankovic, M. S. Seaman, A. K. Chakraborty, B. H. Hahn, M. Caskey, M. C. Nussenzweig, Paired quantitative and qualitative assessment of the replication-competent HIV-1 reservoir and comparison with integrated proviral DNA. Proc Natl Acad Sci U S A 113, E7908-E7916 (2016). https://doi.org/10.1073/pnas.1617789113.

62. P. Mendoza, H. Gruell, L. Nogueira, J. A. Pai, A. L. Butler, K. Millard, C. Lehmann, I. Suarez, T. Y. Oliveira, J. C. C. Lorenzi, Y. Z. Cohen, C. Wyen, T. Kummerle, T. Karagounis, C. L. Lu, L. Handl, C. Unson-O’Brien, R. Patel, C. Ruping, M. Schlotz, M. Witmer-Pack, I. Shimeliovich, G. Kremer, E. Thomas, K. E. Seaton, J. Horowitz, A. P. West, Jr., P. J. Bjorkman, G. D. Tomaras, R. M. Gulick, N. Pfeifer, G. Fatkenheuer, M. S. Seaman, F. Klein, M. Caskey, M. C. Nussenzweig, Combination therapy with anti-HIV-1 antibodies maintains viral suppression. Nature 561, 479–484 (2018). https://doi.org/10.1038/s41586-018-0531-2.

63. K. J. Bar, M. C. Sneller, L. J. Harrison, J. S. Justement, E. T. Overton, M. E. Petrone, D. B. Salantes, C. A. Seamon, B. Scheinfeld, R. W. Kwan, G. H. Learn, M. A. Proschan, E. F. Kreider, J. Blazkova, M. Bardsley, E. W. Refsland, M. Messer, K. E. Clarridge, N. B. Tustin, P. J. Madden, K. Oden, S. J. O’Dell, B. Jarocki, A. R. Shiakolas, R. L. Tressler, N. A. Doria-Rose, R. T. Bailer, J. E. Ledgerwood, E. V. Capparelli, R. M. Lynch, B. S. Graham, S. Moir, R. A. Koup, J. R. Mascola, J. A. Hoxie, A. S. Fauci, P. Tebas, T. W. Chun, Effect of HIV antibody VRC01 on viral rebound after treatment interruption. N Engl J Med 375, 2037–2050 (2016). https://doi.org/10.1056/NEJMoa1608243.

64. D. B. Salantes, Y. Zheng, F. Mampe, T. Srivastava, S. Beg, J. Lai, J. Z. Li, R. L. Tressler, R. A. Koup, J. Hoxie, M. Abdel-Mohsen, S. Sherrill-Mix, K. McCormick, E. T. Overton, F. D. Bushman, G. H. Learn, R. F. Siliciano, J. M. Siliciano, P. Tebas, K. J. Bar, HIV-1 latent reservoir size and diversity are stable following brief treatment interruption. J Clin Invest 128, 3102–3115 (2018). https://doi.org/10.1172/JCI120194.

65. E. Papasavvas, J. R. Kostman, K. Mounzer, R. M. Grant, R. Gross, C. Gallo, L. Azzoni, A. Foulkes, B. Thiel, M. Pistilli, A. Mackiewicz, J. Shull, L. J. Montaner, Randomized, controlled trial of therapy interruption in chronic HIV-1 infection. PLoS Med 1, e64 (2004). https://doi.org/10.1371/journal.pmed.0010064.

66. Y. Z. Cohen, J. C. C. Lorenzi, L. Krassnig, J. P. Barton, L. Burke, J. Pai, C. L. Lu, P. Mendoza, T. Y. Oliveira, C. Sleckman, K. Millard, A. L. Butler, J. P. Dizon, S. A. Belblidia, M. Witmer-Pack, I. Shimeliovich, R. M. Gulick, M. S. Seaman, M. Jankovic, M. Caskey, M. C. Nussenzweig, Relationship between latent and rebound viruses in a clinical trial of anti-HIV-1 antibody 3BNC117. J Exp Med 215, 2311–2324 (2018). https://doi.org/10.1084/jem.20180936.

67. L. Azzoni, E. Papasavvas, J. Kostman, P. Tebas, K. Mounzer, I. Frank, K. M. Lynn, L. Lalley-Chareczko, R. Feng, S. Appel, B. Howell, D. Holder, S. L. Goh, G. Wu, L. M. Montaner, paper presented at the Conference on Retroviruses and Opportunistic Infections, Seattle Washington, 2019. Interferon α2b reduces inducible CD4-associated HIV in ART-suppressed individuals

68. M. Markowitz, M. Louie, A. Hurley, E. Sun, M. Di Mascio, A. S. Perelson, D. D. Ho, A novel antiviral intervention results in more accurate assessment of human immunodeficiency virus type 1 replication dynamics and T-cell decay in vivo. J Virol 77, 5037–5038 (2003). https://doi.org/10.1128/jvi.77.8.5037-5038.2003.

69. T. L. Foster, H. Wilson, S. S. Iyer, K. Coss, K. Doores, S. Smith, P. Kellam, A. Finzi, P. Borrow, B. H. Hahn, S. J. D. Neil, Resistance of transmitted founder HIV-1 to IFITM-mediated restriction. Cell Host Microbe 20, 429-442 (2016). https://doi.org/10.10167i.chom.2016.08.006.

70. D. R. Collins, G. D. Gaiha, B. D. Walker, CD8(+) T cells in HIV control, cure and prevention. Nat Rev Immunol, (2020). https://doi.org/10.1038/s41577-020-0274-9.

71. L. D. Harris, B. Tabb, D. L. Sodora, M. Paiardini, N. R. Klatt, D. C. Douek, G. Silvestri, M. Muller-Trutwin, I. Vasile-Pandrea, C. Apetrei, V. Hirsch, J. Lifson, J. M. Brenchley, J. D. Estes, Downregulation of robust acute type I interferon responses distinguishes nonpathogenic simian immunodeficiency virus (SIV) infection of natural hosts from pathogenic SIV infection of rhesus macaques. J Virol 84, 7886–7891 (2010). https://doi.org/10.1128/JVI.02612-09.

72. Q. Li, A. J. Smith, T. W. Schacker, J. V. Carlis, L. Duan, C. S. Reilly, A. T. Haase, Microarray analysis of lymphatic tissue reveals stage-specific, gene expression signatures in HIV-1 infection. J Immunol 183, 1975–1982 (2009). https://doi.org/10.4049/iimmunol.0803222.

73. M. von Sydow, A. Sonnerborg, H. Gaines, O. Strannegard, Interferon-alpha and tumor necrosis factor-alpha in serum of patients in various stages of HIV-1 infection. AIDS Res Hum Retroviruses 7, 375–380 (1991). https://doi.org/10.1089/aid.1991.7.375.

74. M. R. Betts, J. F. Krowka, T. B. Kepler, M. Davidian, C. Christopherson, S. Kwok, L. Louie, J. J. Eron, H. Sheppard, J. A. Frelinger, Human immunodeficiency virus type 1-specific cytotoxic T lymphocyte activity Is inversely correlated with HIV type 1 viral load in HIV type 1-infected long-term survivors. AIDS Res Hum Retroviruses 15, 1219–1228 (2004). https://doi.org/10.1089/088922299310313.

75. J. A. Warren, G. Clutton, N. Goonetilleke, Harnessing CD8(+) T cells under HIV antiretroviral therapy. Front Immunol 10, 291 (2019). https://doi.org/10.3389/fimmu.2019.00291.

76. M. Bonsignori, M. A. Moody, R. J. Parks, T. M. Holl, G. Kelsoe, C. B. Hicks, N. Vandergrift, G. D. Tomaras, B. F. Haynes, HIV-1 envelope induces memory B cell responses that correlate with plasma antibody levels after envelope gp120 protein vaccination or HIV-1 infection. J Immunol 183, 2708–2717 (2009). https://doi.org/10.4049/jimmunol.0901068.

77. J. L. Mitchell, H. Takata, R. Muir, D. J. Colby, E. Kroon, T. A. Crowell, C. Sacdalan, S. Pinyakorn, S. Puttamaswin, K. Benjapornpong, R. Trichavaroj, R. L. Tressler, L. Fox, V. R. Polonis, D. L. Bolton, F. Maldarelli, S. R. Lewin, E. K. Haddad, P. Phanuphak, M. L. Robb, N. L. Michael, M. de Souza, N. Phanuphak, J. Ananworanich, L. Trautmann, R. V. Rv, R. V. S. Groups, Plasmacytoid dendritic cells sense HIV replication before detectable viremia following treatment interruption. J Clin Invest 130, 2845–2858 (2020). https://doi.org/10.1172/JCI130597.

78. A. Ugarte, Y. Romero, A. Tricas, C. Casado, C. Lopez-Galindez, F. Garcia, L. Leal, Unintended HIV-1 infection during analytical therapy interruption. J Infect Dis 221, 1740–1742 (2020). https://doi.org/10.1093/infdis/jiz611.

79. J. D. Lelievre, L. Hocqueloux, Unintended HIV-1 transmission to a sex partner in a study of a therapeutic vaccine candidate. J Infect Dis 220, S5-S6 (2019). https://doi.org/10.1093/infdis/jiz012.

80. M. R. Abrahams, S. B. Joseph, N. Garrett, L. Tyers, M. Moeser, N. Archin, O. D. Council, D. Matten, S. Zhou, D. Doolabh, C. Anthony, N. Goonetilleke, S. A. Karim, D. M. Margolis, S. K. Pond, C. Williamson, R. Swanstrom, The replication-competent HIV-1 latent reservoir is primarily established near the time of therapy initiation. Sci Transl Med 11, (2019). https://doi.org/10.1126/scitranslmed.aaw5589.

81. B. Julg, L. Dee, J. Ananworanich, D. H. Barouch, K. Bar, M. Caskey, D. J. Colby, L. Dawson, K. L. Dong, K. Dube, J. Eron, J. Frater, R. T. Gandhi, R. Geleziunas, P. Goulder, G. J. Hanna, R. Jefferys, R. Johnston, D. Kuritzkes, J. Z. Li, U. Likhitwonnawut, J. van Lunzen, J. Martinez-Picado, V. Miller, L. J. Montaner, D. F. Nixon, D. Palm, G. Pantaleo, H. Peay, D. Persaud, J. Salzwedel, K. Salzwedel, T. Schacker, V. Sheikh, O. S. Sogaard, S. Spudich, K. Stephenson, J. Sugarman, J. Taylor, P. Tebas, C. T. Tiemessen, R. Tressler, C. D. Weiss, L. Zheng, M. L. Robb, N. L. Michael, J. W. Mellors, S. G. Deeks, B. D. Walker, Recommendations for analytical antiretroviral treatment interruptions in HIV research trials-report of a consensus meeting. Lancet HIV 6, e259-e268 (2019). https://doi.org/10.1016/S2352-3018(19)30052-9.

82. E. N. Borducchi, J. Liu, J. P. Nkolola, A. M. Cadena, W. H. Yu, S. Fischinger, T. Broge, P. Abbink, N. B. Mercado, A. Chandrashekar, D. Jetton, L. Peter, K. McMahan, E. T. Moseley, E. Bekerman, J. Hesselgesser, W. Li, M. G. Lewis, G. Alter, R. Geleziunas, D. H. Barouch, Antibody and TLR7 agonist delay viral rebound in SHIV-infected monkeys. Nature 563, 360–364 (2018). https://doi.org/10.1038/s41586-018-0600-6.

83. L. K. Vibholm, C. V. Konrad, M. H. Schleimann, G. Frattari, A. Winckelmann, V. Klastrup, N. M. Jensen, S. S. Jensen, M. Schmidt, B. Wittig, K. Zuwala, K. Mack, R. Olesen, S. Hua, M. Lichterfeld, L. Ostergaard, P. W. Denton, M. Tolstrup, O. S. Sogaard, Effects of 24-week Toll-like receptor 9 agonist treatment in HIV type 1+ individuals. AIDS 33, 1315–1325 (2019). https://doi.org/10.1097/QAD.0000000000002213.

84. D. M. Margolis, J. V. Garcia, D. J. Hazuda, B. F. Haynes, Latency reversal and viral clearance to cure HIV-1. Science 353, aaf6517 (2016). https://doi.org/10.1126/science.aaf6517.

85. E. Papasavvas, L. Azzoni, A. V. Kossenkov, N. Dawany, K. H. Morales, M. Fair, B. N. Ross, K. Lynn, A. Mackiewicz, K. Mounzer, P. Tebas, J. M. Jacobson, J. R. Kostman, L. Showe, L. J. Montaner, NK response correlates with HIV decrease in pegylated IFN-alpha2a-treated antiretroviral therapy-suppressed subjects. J Immunol 203, 705–717 (2019). https://doi.org/10.4049/jimmunol.1801511.

86. J. F. Salazar-Gonzalez, M. G. Salazar, B. F. Keele, G. H. Learn, E. E. Giorgi, H. Li, J. M. Decker, S. Wang, J. Baalwa, M. H. Kraus, N. F. Parrish, K. S. Shaw, M. B. Guffey, K. J. Bar, K. L. Davis, C. Ochsenbauer-Jambor, J. C. Kappes, M. S. Saag, M. S. Cohen, J. Mulenga, C. A. Derdeyn, S. Allen, E. Hunter, M. Markowitz, P. Hraber, A. S. Perelson, T. Bhattacharya, B. F. Haynes, B. T. Korber, B. H. Hahn, G. M. Shaw, Genetic identity, biological phenotype, and evolutionary pathways of transmitted/founder viruses in acute and early HIV-1 infection. J Exp Med 206, 1273–1289 (2009). https://doi.org/10.1084/jem.20090378.

87. F. Sievers, A. Wilm, D. Dineen, T. J. Gibson, K. Karplus, W. Li, R. Lopez, H. McWilliam, M. Remmert, J. Soding, J. D. Thompson, D. G. Higgins, Fast, scalable generation of high-quality protein multiple sequence alignments using Clustal Omega. Mol Syst Biol 7, 539 (2011). https://doi.org/10.1038/msb.2011.75.

88. A. Stamatakis, RAxML version 8: a tool for phylogenetic analysis and post-analysis of large phylogenies. Bioinformatics 30, 1312–1313 (2014). https://doi.org/10.1093/bioinformatics/btu033.

89. J. Maydt, T. Lengauer, Recco: recombination analysis using cost optimization. Bioinformatics 22, 1064–1071 (2006). https://doi.org/10.1093/bioinformatics/btl057.

90. The R Project for Statistical Computing, https://www.r-project.org/.

91. B. Carpenter, A. Gelman, M. D. Hoffman, D. Lee, B. Goodrich, M. Betancourt, M. Brubaker, J. Guo, P. Li, A. Riddell, Stan: A Probabilistic Programming Language. Journal of Statistical Software 76, (2017). https://doi.org/10.18637/jss.v076.i01.

